# A rigorous evaluation of optimal peptide targets for MS-based clinical diagnostics of Coronavirus Disease 2019 (COVID-19)

**DOI:** 10.1101/2021.02.09.21251427

**Authors:** Andrew T. Rajczewski, Subina Mehta, Dinh Duy An Nguyen, Björn A. Grüning, James E. Johnson, Thomas McGowan, Timothy J. Griffin, Pratik D. Jagtap

## Abstract

The Coronavirus Disease 2019 (COVID-19) global pandemic has had a profound, lasting impact on the world’s population. A key aspect to providing care for those with COVID-19 and checking its further spread is early and accurate diagnosis of infection, which has been generally done via methods for amplifying and detecting viral RNA molecules. Detection and quantitation of peptides using targeted mass spectrometry-based strategies has been proposed as an alternative diagnostic tool due to direct detection of molecular indicators from non-invasively collected samples as well as the potential for high-throughput analysis in a clinical setting; many studies have revealed the presence of viral peptides within easily accessed patient samples. However, evidence suggests that some viral peptides could serve as better indicators of COVID-19 infection status than others, due to potential misidentification of peptides derived from human host proteins, poor spectral quality, high limits of detection etc. In this study we have compiled a list of 639 peptides identified from Sudden Acute Respiratory Syndrome Coronavirus 2 (SARS-CoV-2) samples, including from in vitro and clinical sources. These datasets were rigorously analyzed using automated, Galaxy-based workflows containing tools such as PepQuery, BLAST-P, and the Multi-omic Visualization Platform as well as the open-source tools MetaTryp and Proteomics Data Viewer (PDV). Using PepQuery for confirming peptide spectrum matches, we were able to narrow down the 639 peptide possibilities to 87 peptides which were most robustly detected and specific to the SARS-CoV-2 virus. The specificity of these sequences to coronavirus taxa was confirmed using Unipept and BLAST-P. Applying stringent statistical scoring thresholds, combined with manual verification of peptide spectrum match quality, 4 peptides derived from the nucleocapsid phosphoprotein and membrane protein were found to be most robustly detected across all cell culture and clinical samples, including those collected non-invasively. We propose that these peptides would be of the most value for clinical proteomics applications seeking to detect COVID-19 from a variety of sample types. We also contend that samples taken from the upper respiratory tract and oral cavity have the highest potential for diagnosis of SARS-CoV-2 infection from easily collected patient samples using mass spectrometry-based proteomics assays.

## Introduction

In the latter half of 2019, a pneumonia-like disease arose in the Wuhan Province of China^1^. Subsequent analysis showed the cause to be a betacoronavirus initially called 2019-novel coronavirus (2019-nCoV). This disease soon spread throughout the world and came to be known as coronavirus disease 2019 (COVID-19) with the clinical classification Sudden Acute Respiratory Syndrome Coronavirus 2 (SARS-CoV-2). As of the writing of this manuscript, there are over 106 million patients infected world-wide with COVID-19, with a current global death toll sitting at 2.3 million people^2^. Patients report a litany of symptoms, ranging from fever, cough, and muscle aches in mild cases to acute respiratory distress syndrome (ARDS), multiple-organ failure, and death in the most severe cases^3, 4^.

While the development of therapeutic treatments for infected patients^5, 6^ and the eventual development of vaccines against SARS-CoV-2^7-9^ are of great importance for the management of this disease, rapid and effective diagnosis of COVID-19 infection has been and continues to be of primary importance. Most testing strategies used in the diagnosis of active COVID-19 infections utilize quantitative Reverse Transcription Polymerase Chain Reaction (RT-qPCR) of viral RNA in samples collected from patients^10, 11^. Rapid COVID-19 testing is generally performed on readily accessible patient-derived samples with high viral loads, such as nasopharyngeal swabs and saliva. To improve turnover time and increase the volume of tests that can be performed, innovations in RNA-based testing have been introduced to cut down on the time required. Testing protocols have been developed that eschew the isolation of RNA from patient samples, allowing for much faster RT-qPCR analyses^12^. In addition, techniques such as Reverse Transcription Loop-mediated isothermal AMPlification (RT-LAMP)^13^ and Specific High Sensitivity Enzymatic Reporter UnLOCKing (SHERLOCK)^14^ diagnostics allow for rapid point-of-care detection of SARS-CoV-2 RNA without the need for sophisticated training in PCR.

While these techniques are generally fast and highly specific for viral RNA, improper sample collection, storage, or processing could result in the degradation of RNA yielding potential false negative tests. In addition, their reliance on sequence amplification using reverse transcriptases and DNA polymerases introduces the potential for false negatives through the inhibition of these enzymes by components of the sample^15, 16^. Due to the better chemical stability of proteins compared to RNA, as well as the lack of a need for intermediary enzymes and signal amplification via PCR, clinical proteomics has emerged as a potential supplemental test for the diagnosis of COVID-19 through direct detection of viral peptides via LC-MS^17^. Specifically, targeted methods such as selected reaction monitoring (SRM) and parallel reaction monitoring (PRM) to detect peptides specific to the virus could be most useful in a clinical setting^18, 19^. However, not all the potential viral peptides derived from SARS-CoV-2 infection are equally suitable as targets, based on well-known limitations of targeted LC-MS methods for proteomics; some tryptic peptides of SARS-CoV-2 could have intrinsic physicochemical properties limiting their reproducible detection in a mass spectrometer, as well as co-elution from the LC with more abundant peptides that mask their presence in the sample. In addition, proteomics software can sometimes make putative peptide spectrum matches (PSMs) with spectra that are of poor quality, making for uncertain identification of peptides of interest^20, 21^. Additionally, a key requirement for targeting peptides for virus detection is that these are specific to the SARS-CoV-2 virus, with no potential overlap with other coronaviruses or other organisms.

In order to evaluate the most robustly detectable SARS-CoV-2 peptides, and make the detection of these viral peptides in human samples in a clinical setting all the more feasible, we set out to examine proteomic datasets from three cell culture-based studies^22-24^ and five clinical studies^25-29^. We utilized automated workflows implemented in the Galaxy platform and made accessible via the European Galaxy public instance to first identify as many SARS-CoV-2 peptides possible in all samples, creating a master list of SARS-CoV-2 peptides identified across the samples. We then interrogated these peptides using the PepQuery search engine^30^ to confirm the quality of these PSMs and determine whether the matched sequences were unique to SARS-CoV-2 or could be better ascribed to the human proteome or that of another closely related coronavirus. Peptides and their associated PSMs which survived this rigorous filtering were then manually validated using the Multi-omics Visualization Platform^31^ and further analyzed for specificity to the SARS-CoV-2 virus via BLAST-P^32^ and MetaTryp^33^. Taken together, our analyses enable the construction of a high-confidence target peptide list that would form the basis of a targeted clinical proteomics assay for SARS-CoV-2 infection.

## Methods

### Case Study

For establishing workflows to evaluate virus-specific peptides, three published cell culture datasets^22-24^ which used SARS-CoV-2 infected Vero cell lines were chosen, along with five clinical datasets^25-29^.

### Cell Culture Datasets

Gouveia *et al*. published a dataset (PXD018804) with SARS-CoV-2 infected Vero cells from *Chlorocebus* primates to generate a high-resolution mass spectrometry dataset. The second dataset was published by Grenga *et al*. (PXD018594) wherein a seven-day time course shotgun proteomics study was performed on Vero E6 cells infected by Italy-INMI1 SARS-CoV-2 virus at two multiplicities of infection. The third cell culture dataset chosen was published by Davidson *et al*. (PXD018241), which also utilized Vero E6 cells to investigate the viral transcriptome and proteome.

### Clinical Datasets

The first clinical dataset chosen was from the study by Cardozo et al. (PXD021328), wherein they collected bottom-up mass spectrometry (MS) data on combined oropharyngeal and nasopharyngeal samples from ten COVID-19 positive patient samples. The second clinical dataset was from the Ihling group (PXD019423) to detect SARS-CoV-2 virus proteins from saline gargle samples of COVID-19 infected patients. The last dataset was obtained from the Rivera group (PXD020394), where they performed comparative quantitative proteomic analysis from oro- and naso-pharyngeal swabs used for COVID-19 diagnosis. Datasets derived from COVID-19 patient lung biopsies (PXD018094) and bronchoalveolar lavage fluid (BALF) (PXD022085) were analyzed to determine the utility of our workflow to identify SARS-CoV-2 in clinically relevant sample types.

### Sequence Database Searching

The Galaxy workflow for peptide identification (**Figure 1, Figure 2a**) includes conversion of RAW MS/MS datafiles derived from Thermo Fisher instruments to MGF and mzML format. In the case of the cell culture study, the MGF files are searched against the combined database of *Chlorocebus* sequences, contaminant proteins (cRAP) and SARS-Cov-2 proteins. For the clinical database, the resultant MGF files were searched against the combined database of Human Uniprot proteome, contaminants, and SARS-Cov-2 proteins database.

**Figure 1:**
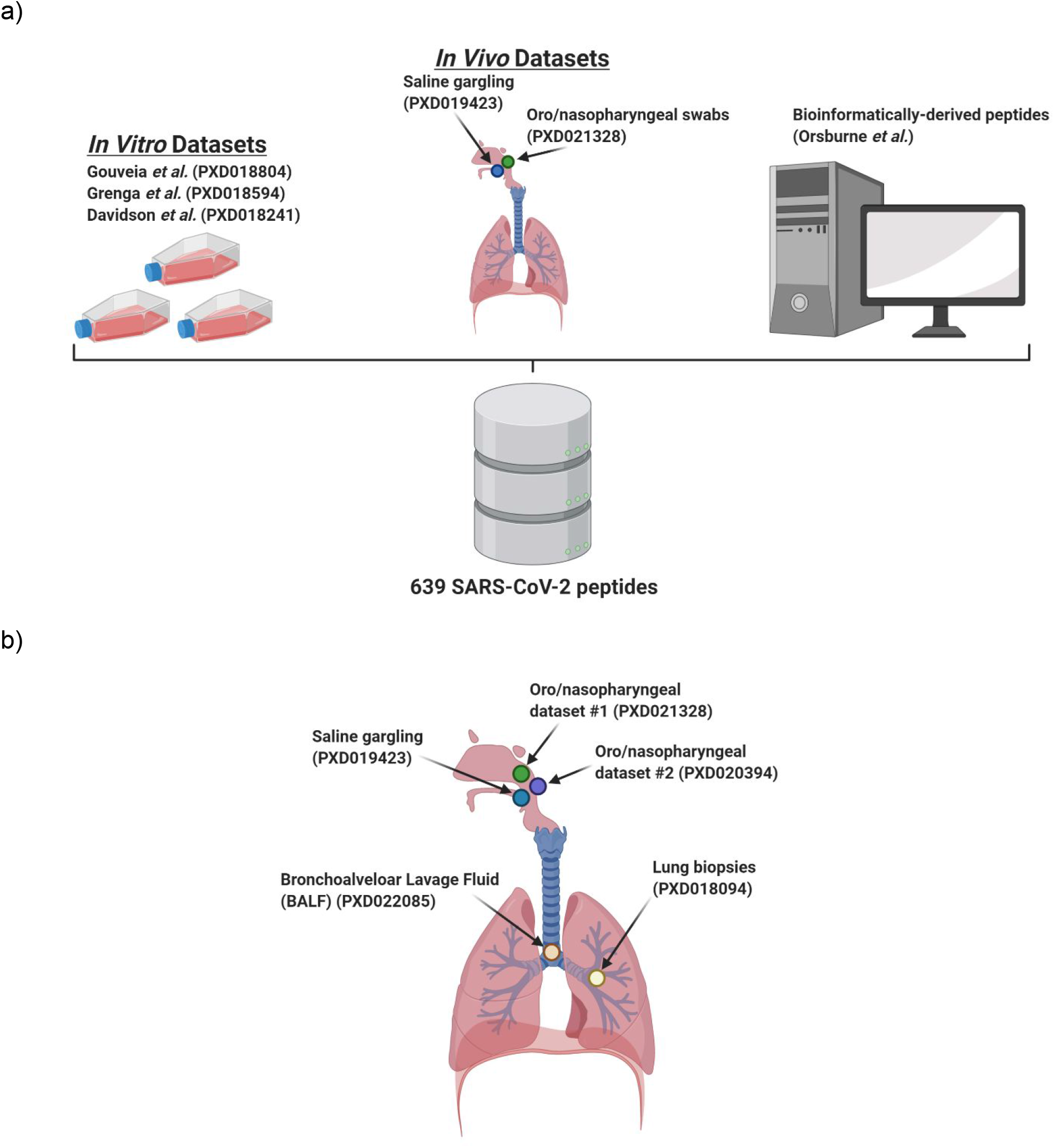
MS/MS datasets used in the determination of optimal SARS-CoV-2 peptides for COVID-19 diagnosis. a) Cell culture, clinical, and bioinformatic datasets used to generate the SARS-CoV-2 peptide panel. b) Clinical datasets queried using the initially characterized peptide panel from a) to determine the feasibility of COVID-19 diagnosis via targeted proteomics as well as determine the optimal peptide targets for those assays. Figures were made using BioRender.

**Figure 2:**
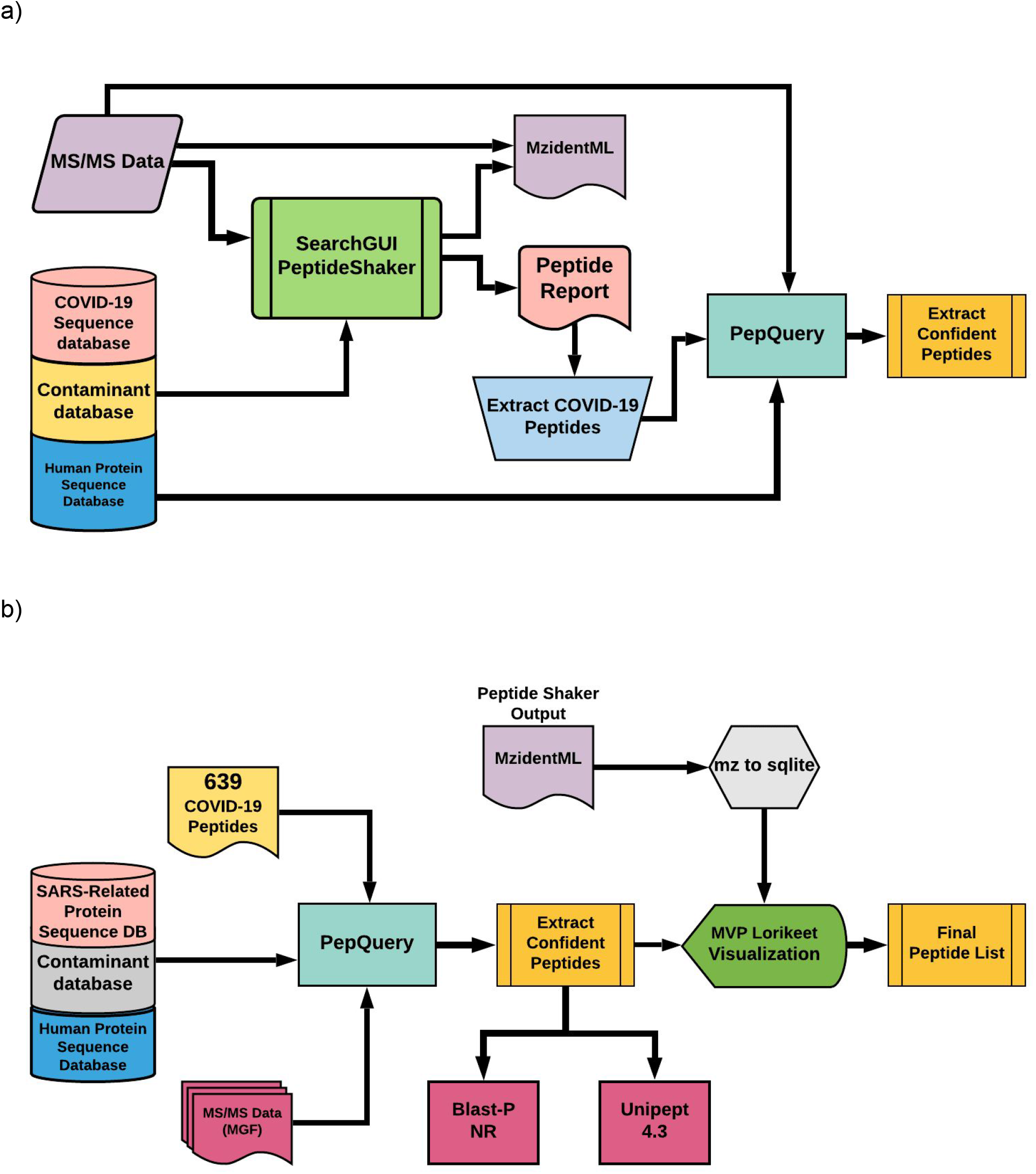
Workflows used in the interrogation of MS-data to identify and validate SARS-CoV-2 peptides. a) Galaxy-based sequence database searching workflow to detect and confirm SARS-CoV-2 peptides. MS/MS spectra from cell culture or clinical datasets were searched against appropriate protein sequence databases (protein sequences from COVID-19, contaminants, and Human Protein sequences) using SearchGUI/ Peptide Shaker. The peptide output was filtered to extract COVID-19 peptides and the output was confirmed using PepQuery to extract confident peptides. mzidentML generated through this workflow was subsequently used for analysis in the Lorikeet viewer; b) Workflow to validate detected SARS-CoV-2 peptides. A list of 639 Peptides (theoretical and validated peptides obtained from the cell-culture and clinical datasets) was subjected to PepQuery analysis of COVID-19 datasets to identify the presence of SARS-CoV-2 peptides. The quality of the peptide spectral matches (PSMs) was reviewed using Lorikeet visualization within the Multi-omics Visualization Platform for further validation. Peptides were also searched against the NCBI-non redundant database and Unipept 4.3 for taxonomic annotation.

For sequence database searching in the workflow, the search algorithms - X! tandem, MSGF+, OMSSA were used within SearchGUI^34^ to generate PSMs, followed by False Discovery Rate (FDR) and protein grouping analysis using PeptideShaker^35^. The search parameters for digestion, modifications, tolerance, and FDR were chosen accordingly from the published methods for each of these datasets (**Supplementary Data 1**). The peptide report generated using PeptideShaker was used to extract confident COVID-19 peptides. The peptides were validated using PepQuery analysis with MS tolerance of 10 ppm and MS/MS tolerance of 0.05 Da. The SARS-CoV-2 peptides detected from the three cell culture datasets and two clinical datasets were merged with the peptide list from *in silico* analysis of genomic sequences by Orsburn *et al*.^36^ and target peptides from Gouveia et al.^*22*^ to generate a peptide panel for interrogation of clinical data sets. The re-analysis of the dataset using the workflow is available online on the COVID-Galaxy website (https://COVID-19.galaxyproject.org/proteomics) and the workflows and outputs can be found <u>online</u> (see Data and Workflow Availability).

### Peptide Validation

The SARS-CoV-2 peptide panel identified by sequence database searching and comprising 639 peptides was subjected to the Peptide Validation workflow (**Figure 2b**), interrogating the clinical MS/MS datasets described above. The peptide validation workflow includes re-analysis by PepQuery as well as manual visualization and inspection in the Lorikeet application of Multi-omics Visualization Platform (MVP) to ascertain the quality of peptide sequences matched to MS/MS spectra. The workflow also included optional in-line characterization of these peptides by Galaxy-based searching against NCBI-non redundant (nr) BLAST-P and Unipept^37^ analysis. Further offline analysis (non-Galaxy based) was performed using NCBI BLAST-P analysis as well as the MetaTryp^33^ coronavirus database.

## Results

### Sequence Database Searching Results

Sequence database searching to generate peptide spectrum matches (PSMs) and identify peptides from the three cell culture datasets (**Figure 1a**) using the workflow shown in Figure 1a led to detection of 139 peptides, 99 peptides and 579 peptides, respectively. For the two clinical datasets analyzed using the workflow, we detected 76 and 8 peptides, respectively (**Table 1**). These peptides together represented 630 unique peptides corresponding to several proteins coded in the SARS-CoV-2 genome; to these we then added a further 6 unique peptides generated from *in silico* translated data by Orsburn *et al*^36^ and 3 peptides exclusively detected by Gouveia *et al*.^*22*^ from the deep proteomic analysis of CoviD-19 virus infected Vero cells, to generate a list of 639 unique SARS CoV-2 peptides (**Supplemental Table 1**). This 639-peptide panel was further used to interrogate the clinical datasets and determine the reliability of their detection using shotgun MS-based proteomics. BLAST-P analysis of the 639-peptide panel showed that these peptides mapped to 27 proteins and open reading frames within the SARS-CoV-2 genome (**Figure 3**), with individual protein sequence coverage ranging from 4.7% coverage (Proofreading exoribonuclease Guanine-N7 methyltransferase protein) to 93.7% coverage (Nucleocapsid protein) (**Supplementary Figure 1**).

**Table 1:**
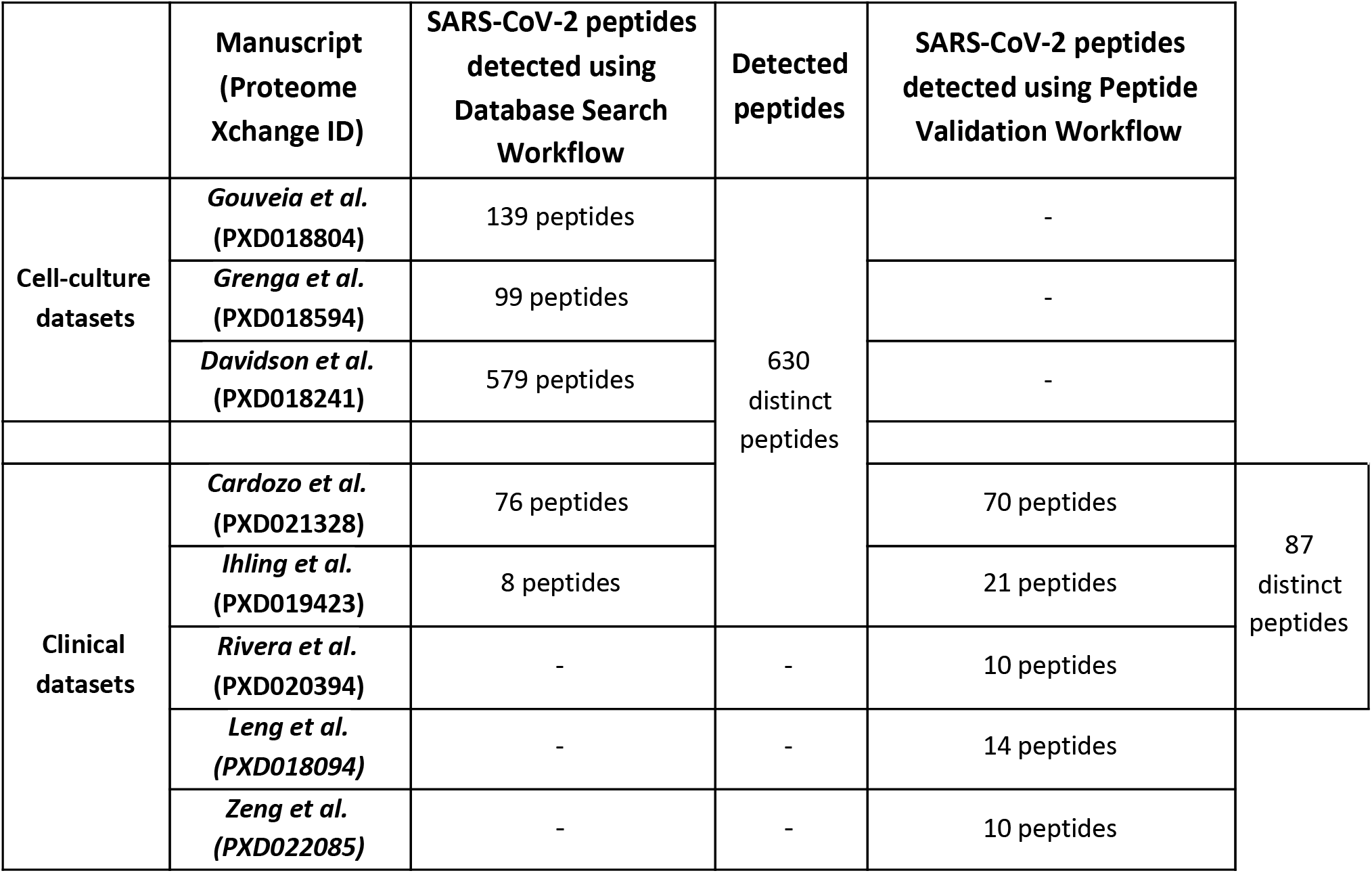
Peptides generated from MS datasets.

**Figure 3:**
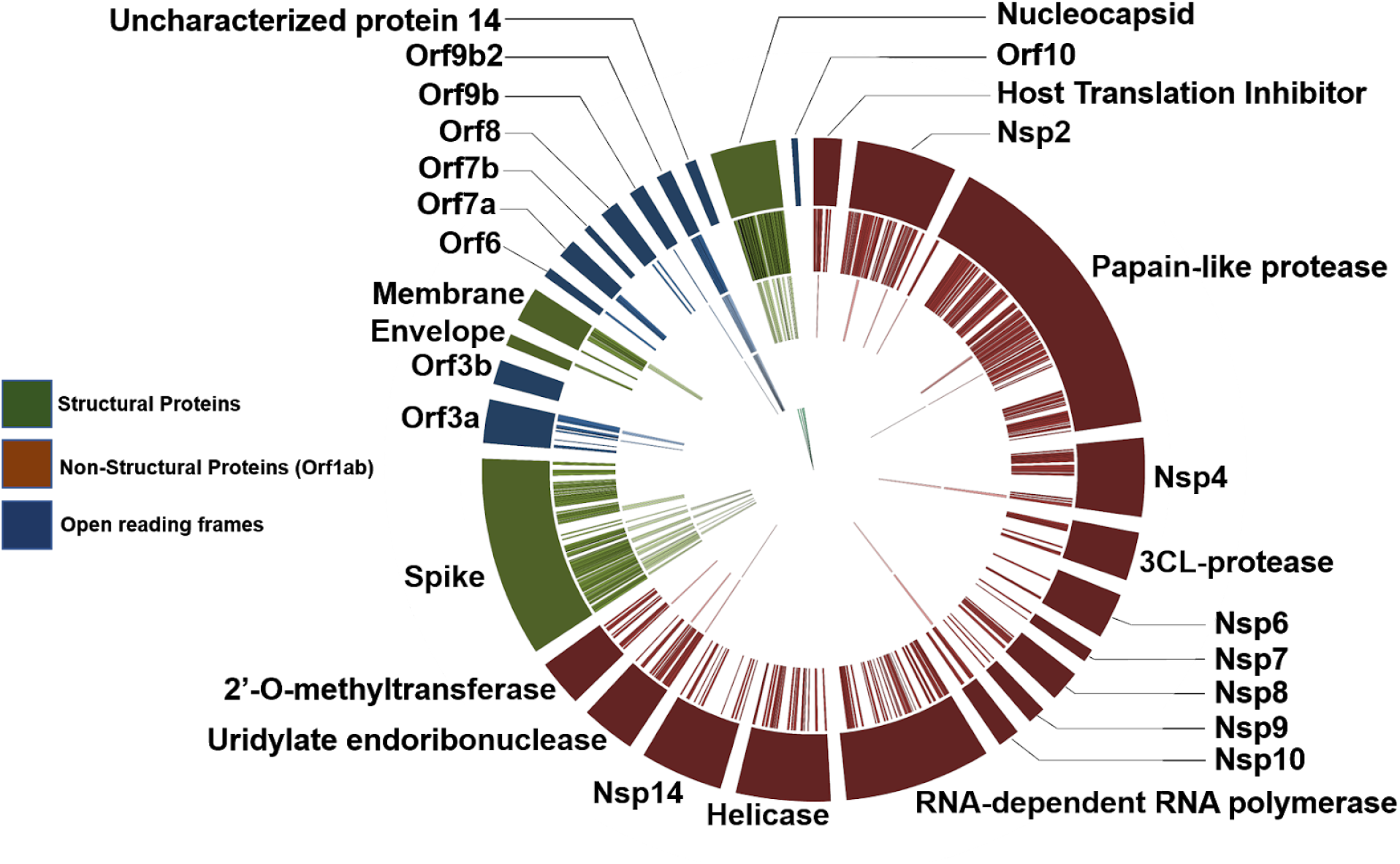
Protein assignment of detected and validated SARS-CoV-2 peptides: Circos plot of peptides against SARS-CoV-2 proteins (outermost ring). Of the 639-peptide panel (2^nd^ outermost ring), many peptides could be identified using our validation workflow in clinical and cell culture datasets (3^rd^ outermost ring). Peptides derived from ORF9b, papain-like protease, Nsp4, Nsp10, uridylate endoribonuclease (Nsp15) and certain spike protein peptides were only found in cell culture datasets (2^nd^ innermost ring). Final peptides chosen for targeted analysis are annotated in the innermost ring. Circos plot was generated in Galaxy^38^.

### Peptide Validation Results

Having derived a comprehensive panel of 639 peptides detected across multiple COVID-19 datasets, we then utilized a validation workflow based around the PepQuery tool to interrogate the dataset PXD020394, derived from oro- and naso-pharyngeal swabs collected in the clinic from patients positive and negative for COVID-19. This resulted in validation of 10 SARS-CoV-2 peptides from our panel matching to MS/MS spectra generated in these clinically relevant samples (**Supplementary Figure 2**).

We detected eight of these 10 peptides in COVID-19 positive sample replicates - with the peptide RGPEQTQGNFGDQELIR being detected in all positive sample replicates, followed by TATKAYNVTQAFGR and AYNVTQAFGR detected in 6 out of 10 replicate samples (**Supplementary Figure 2**). We also detected two peptides-GVEAVMYMGTLSYEQFK and CDLQNYGDSATLPK-from COVID-19 negative samples.

We also re-analyzed the two clinical datasets used in the generation of the 639 panel (the second oro/nasopharyngeal dataset from Cardozo *et al*. as well as the saline gargling dataset), using our validation workflow. The validation workflow provides a complementary method to the initial sequence database searching method for confirming peptide spectrum matches, based primarily on the PepQuery tool. For the oro/nasopharyngeal dataset, we confirmed confident identification of 70 peptides using the peptide validation workflow (as compared to 76 detected using the initial sequence database searching workflow). For the saline gargling dataset, we confirmed the presence of 21 peptides using the peptide validation workflow (as compared to 8 peptides detected using the peptide search workflow). Considering all peptides detected in clinical samples using the peptide validation workflow, we detected 87 peptides with confidence (**Table 1**). These validated peptides were assigned to known proteins from the COVID-19 proteome. Most of the peptides detected in the upper respiratory tract were aligned to structural proteins making up the viral capsid such as nucleocapsid protein N, the viral matrix protein M, and the spike protein S; fewer peptides were aligned to proteins involved in viral replication such as papain-like protease, RNA-directed RNA polymerase, non-structural protein, 2’-O-methyltransferase and host translation inhibitor (**Figure 3**). The highest number of peptides were identified in the oro/nasopharyngeal dataset which consisted of combined oropharyngeal and nasopharyngeal swabs analyzed by Cardozo *et al*.; fewer peptides were identified from PXD019423 and PXD020493, which were derived from gargled saline samples and a second study of combined oropharyngeal and nasopharyngeal samples, respectively.

Based on the sample-type from which they were detected (clinical samples versus *in vitro* cell culture experiments) and their source (empirically derived from MS/MS data versus theoretically determined based on genomic sequence data), we categorized them as being present or absent in the various datasets based on their confident detection using our validation workflow. We found that the validated peptides clustered into distinct groups based on their source sample and dataset of origin, and how they were originally identified (**Supplemental Table 1**). Eleven peptides were found to be highly consistent across the upper respiratory clinical datasets as well as the in vitro cell culture datasets. In considering theoretical peptides proposed by Orsburn *et al*., eleven of those predicted peptides were in clinical samples and eight were detected in the *in vitro* cell culture samples. Twenty-two SARS-CoV-2 peptides that were not initially identified using the database search workflow were identified by matching to MS/MS spectra using the PepQuery-based validation workflow across multiple datasets.

Clinical datasets from lung biopsies (PXD018094) and BALF (PXD022085) were interrogated using our PepQuery validation workflow and the 639-peptide panel to determine the applicability of our approach in detecting SARS-CoV-2 within the deeper respiratory tract. Our validation workflow was able to confidently match MS/MS to 15 peptides in the lung biopsy dataset and 37 peptides in the BALF dataset. In comparing the peptides found within the upper respiratory samples to those detected within the lung biopsy samples and the BALF samples, it is clear that the majority of the peptides detected in the deep lung datasets are unique to the sample being analyzed, with no peptides common to all three of the upper respiratory tract samples (**Supplementary Table 1**). Despite this apparent disparity, BLAST-P analysis reveals the alignment of SARS-CoV-2 peptides identified in deep lung tissue corresponding to a similar complement of SARS-CoV-2 proteins as the upper respiratory tract datasets, including additional structural proteins such as the Spike protein and Membrane glycoprotein as well as other nonstructural and replication proteins such as RNA-directed RNA polymerase, Protease 3CL-PRO, etc. In addition, the lung biopsy and BALF datasets also included MS-data from patients negative for COVID-19. In contrast to the 2 SARS-CoV-2 PSMs identified in the oro/nasopharyngeal samples from COVID-19-negative patients, samples analyzed from lung biopsies of COVID-19-negative patients identified 21 SARS-CoV-2 peptides. Similarly, 37 peptides were detected in BALF samples isolated from patients that tested negative for COVID-19.

The last category of peptides that we evaluated were detected from COVID-19 cell culture studies (**Supplementary Table 1 and Supplementary Figure 3**). These peptides were derived from protein sequences that were not available in the initial Uniprot sequence databases but were subsequently added as more COVD19 strains were sequenced^39, 40^. We added these sequences to the sequence database to enable the detection of these COVID-19 proteoforms. Using this updated sequence database, we detected and validated twelve peptides from Accessory protein ORF9b from SARS-CoV-2 and two peptides from ORF1ab polyprotein from SARS-CoV-2. These peptides were observed only in the cell culture datasets, and not in the clinical datasets (**Figure 3**).

### Identifying detected peptides with highest spectral quality

As a quality check on our bioinformatic workflows, we utilized the Multi-omics Visualization platform^31^ (MVP) to manually assess the spectral quality of the peptides that passed PepQuery validation, as well as elucidate the distribution of these peptides throughout the six datasets we analyzed. In order to be useful for targeted MS-based assays for detecting SARS-CoV-2, it is critical that the peptides used as targets have excellent spectral quality to ensure adequate reliability in detecting and quantifying these peptides across a variety of clinical samples. Here, we focused on four peptides (AYNVTQAFGR, MAGNGGDAALALLLLDR, RGPEQTQGNFGDQELIR, DGIIWVATEGALNTPK) found in the positive patients from the second oro/nasopharyngeal dataset (PXD020934) that were also seen in the other clinical datasets as well as the two peptides found in the negative patients (CDLQNYGDSATLPK, GVEAVMYMGTLSYEQFK) from the same oro/nasopharyngeal dataset as benchmark examples for manually validating our spectra. For these selected peptides, in the four virus-positive samples we found a largely complete b- and/or y-ion series with at least three consecutive ions detected in either series (**Supplementary Figure 3**). In addition, we found that these reporter ions showed intensities considerably higher than the background noise of the spectra. By contrast, the two peptides found in the negative samples had a very few fragment ions detected which scarcely rose above the level of the background noise (**Supplementary Figure 3**). Together, the MS/MS spectra of these six peptides were used to generate guidelines which were then used to manually interrogate the rest of the SARS-CoV-2 spectra as being high or low confidence by the bioinformatics software and found that 16 of the peptides validated in PepQuery had MS/MS spectra suitable for confident identification.

As a part of our investigation we detected and validated eight peptides that were predicted by Orsburn *et al*. (**Supplementary Table 1, Supplementary Figure 3**). However, Lorikeet visualization of the PSM quality detected only two peptides (with sequences ADETQALPQR and FDNPVLPFNDGVYFASTEK) in the clinical sample PXD021328 dataset; of these the ADETQALPQR was also detected in all three cell cultures sample datasets while the FDNPVLPFNDGVYFASTEK sequence peptide was detected in two of the three cell culture samples (**Supplementary Table 1, Supplementary Figure 3**). All the eight peptides were found to have good quality of PSMs in the cell culture datasets by using manual validation. Out of these eight peptides, a peptide with sequence HTPINLVR was detected in all cell culture experimental datasets (**Supplementary Table 1**).

We were able to validate 22 peptides using PepQuery which were not detected in the database search workflow (**Supplementary Table 1**). Subsequent manual validation of these peptides determined only two peptides had high quality spectral annotations. The peptide of sequence DGIIWVATEGALNTPKDHIGTR was validated by using PepQuery and manual visualization in the PXD019423 dataset along with another peptide with sequence FTALTQHGKEDLK from the PXD02132 dataset (**Supplementary Figure 3**).

In order to determine the optimal candidates for the detection of SARS-CoV-2 using clinical MS-based assays, we resolved to focus on those peptides that passed PepQuery with the highest confidence, and subject these to manual inspection of spectral quality. We therefore sorted the results of our PepQuery analyses to include only those which had the highest confidence possible (p-value < 0.0001) to maximize the likelihood of passing our spectral annotation thresholds. In filtering the clinical datasets, we see a notable difference between the datasets derived from the upper respiratory tract (oro/nasopharyngeal datasets 1 and 2 as well as the saline gargling dataset) and those derived from deep lung tissue (the lung biopsy and BALF datasets) (**Figure 4**). In filtering the PepQuery results from the upper respiratory tract datasets, we noted that the structural proteins that had the most identified peptides-the nucleocapsid, membrane protein, and spike proteins-show relatively little elimination of PSMs based on our statistical thresholds, while the proteins involved in viral replication are generally lost, indicating relatively high confidence in the PepQuery validation of the peptides of the viral structural proteins. By contrast, peptides found in all proteins in the lung biopsy and BALF datasets were filtered out at this step, yielding only 3 and 4 high confidence peptides in each dataset, respectively, leaving single peptides of nucleocapsid, membrane protein, and spike protein in the lung biopsy samples and single peptides of the spike protein, papain-like protease, non-structural protein 2, and RNA-dependent RNA polymerase.

**Figure 4:**
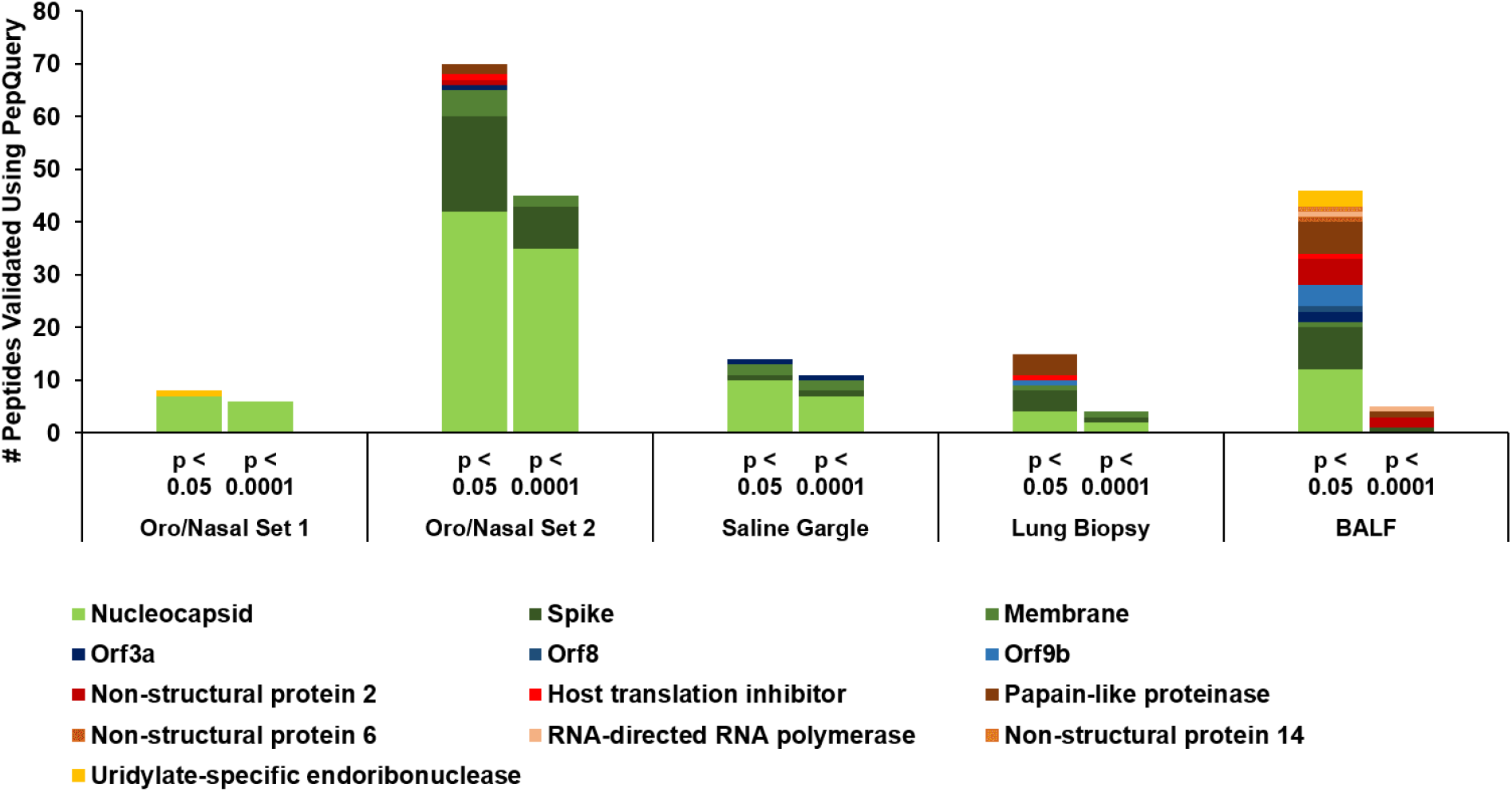
Peptide spectral matches (PSMs) of SARS-CoV-2 peptides in the upper respiratory clinical datasets are of higher confidence than deep lung datasets. PSMs validated in oro/nasopharyngeal datasets, saline gargling samples, lung biopsy samples, and bronchoalveolar lavage fluids (BALF) using PepQuery as grouped into the proteins they aligned to; columns correspond to those peptides that passed PepQuery validation with minimal required confidence (left) as well as those associated with higher confidence (right).

The spectra of those peptides found to have high-confidence in the clinical datasets (based on PepQuery scoring) were then analyzed using MVP, which leverages the Lorikeet viewer for visualization of annotated peptide MS/MS spectra. Manual analysis of the high confidence peptides detected in the lung biopsy and BALF datasets using our previously established guidelines showed only the single peptide FLALCADSIIIGGAK, a component of Non-structural protein 2 in the BALF dataset as having a good quality spectrum, suggesting that the use of clinical samples collected using more invasive methods from deep within the lung may be unsuitable for detection of SARS-CoV-2 using a clinical proteomics strategy. In contrast, 11 peptides in the upper respiratory tracks had high-confidence and high-quality MS/MS-spectra. Of these, we then chose four peptides-MAGNGGDAALALLLLDR, DGIIWVATEGALNTPK, RGPEQTQGNFGDQELIR, and IGMEVTPSGTWLTYTGAIK-which were identified in at least two of the three upper respiratory clinical datasets, determining these to be the most reliable peptides for proteomics-based detection of SARS-CoV-2 (**Figure 5**).

**Figure 5:**
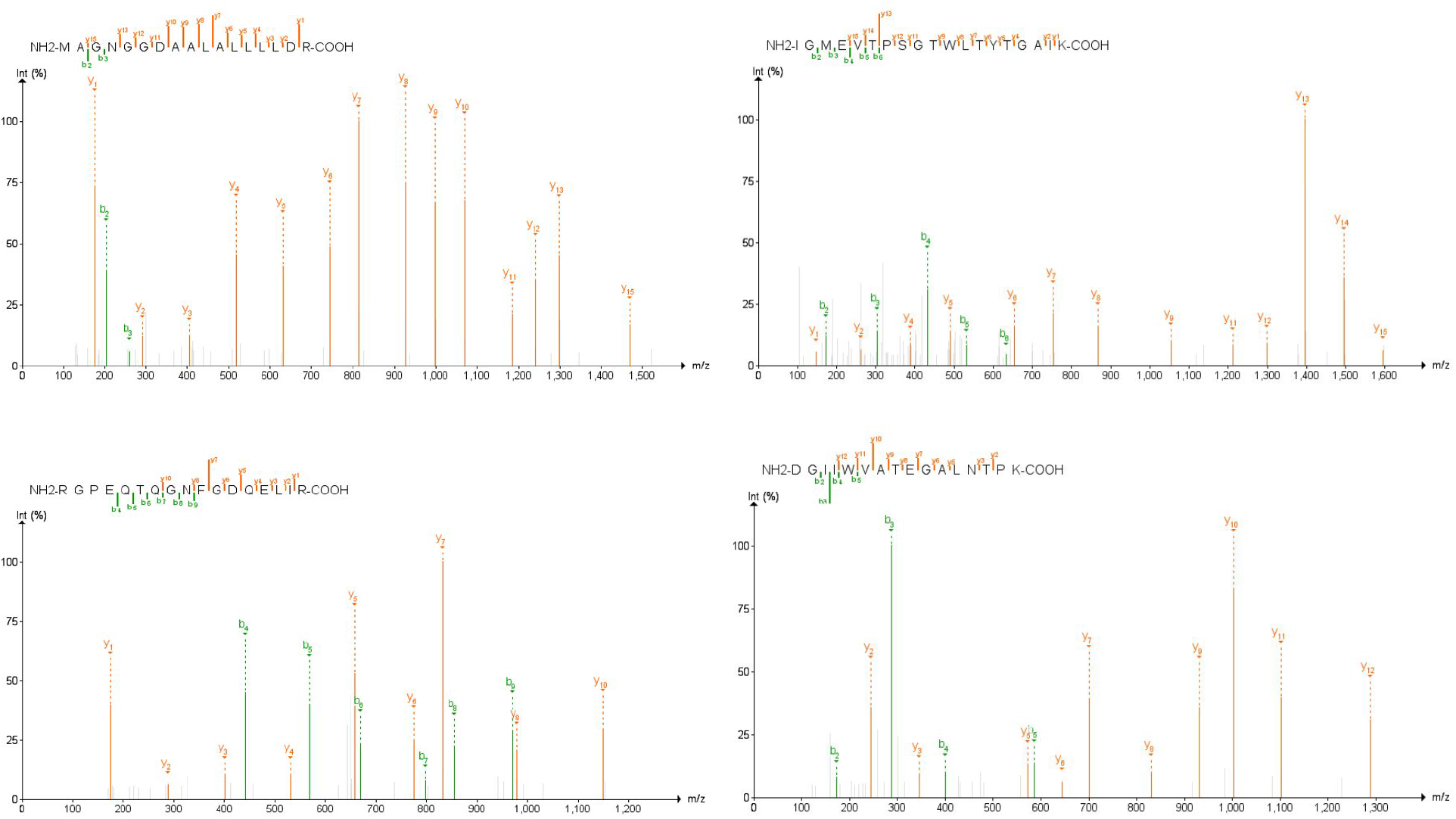
MS/MS spectra of SARS-CoV-2 peptides most confidently identified in PepQuery (p-value < 0.001) and across the most clinical samples. Spectral annotation quality was interrogated using the Lorikeet viewer implemented within the Multi-Omics Viewing Platform (MVP); images for annotated PSMs for these peptides were created using the PDV platform from the Zhang lab^41^.

### Viral specificity of high-quality peptides detected in SARS-CoV-2

We performed taxonomic analysis using MetaTryp 2.0 to validate the specificity of the four highest-quality peptides detected in clinical samples to coronaviruses (**Figure 6a**). Using this we found that these peptides mapped to proteomes of several coronaviruses, with each showing alignment to SARS-CoV-2. To gauge the degree of specificity of these peptides for SARS-CoV-2 over other coronaviruses and their potential human host, we performed BLAST-P analysis of these peptides against proteomes for SARS-CoV-2, humans, and eight known pathogenic human coronaviruses. To interrogate all possible matches to the target organisms, a relatively relaxed E-value cutoff of 1 was used. In considering the sequence alignment of these peptides, the peptides examined found a high degree of alignment to the nucleocapsid protein (N-protein) of SARS-CoV-2 (**Figure 6b**). Each of the four distinct peptides that showed alignment to the N-protein also showed 100% sequence homology uniquely to SARS-CoV-2, with decreased sequence alignment in other closely related coronaviruses. One peptide sequence, MAGNGGDAALALLLLDR, showed perfect alignment to the SARS-CoV-2 nucleocapsid protein with no alignment to the same protein in any other viruses. In all cases, no alignment to any human proteins was noted.

**Figure 6:**
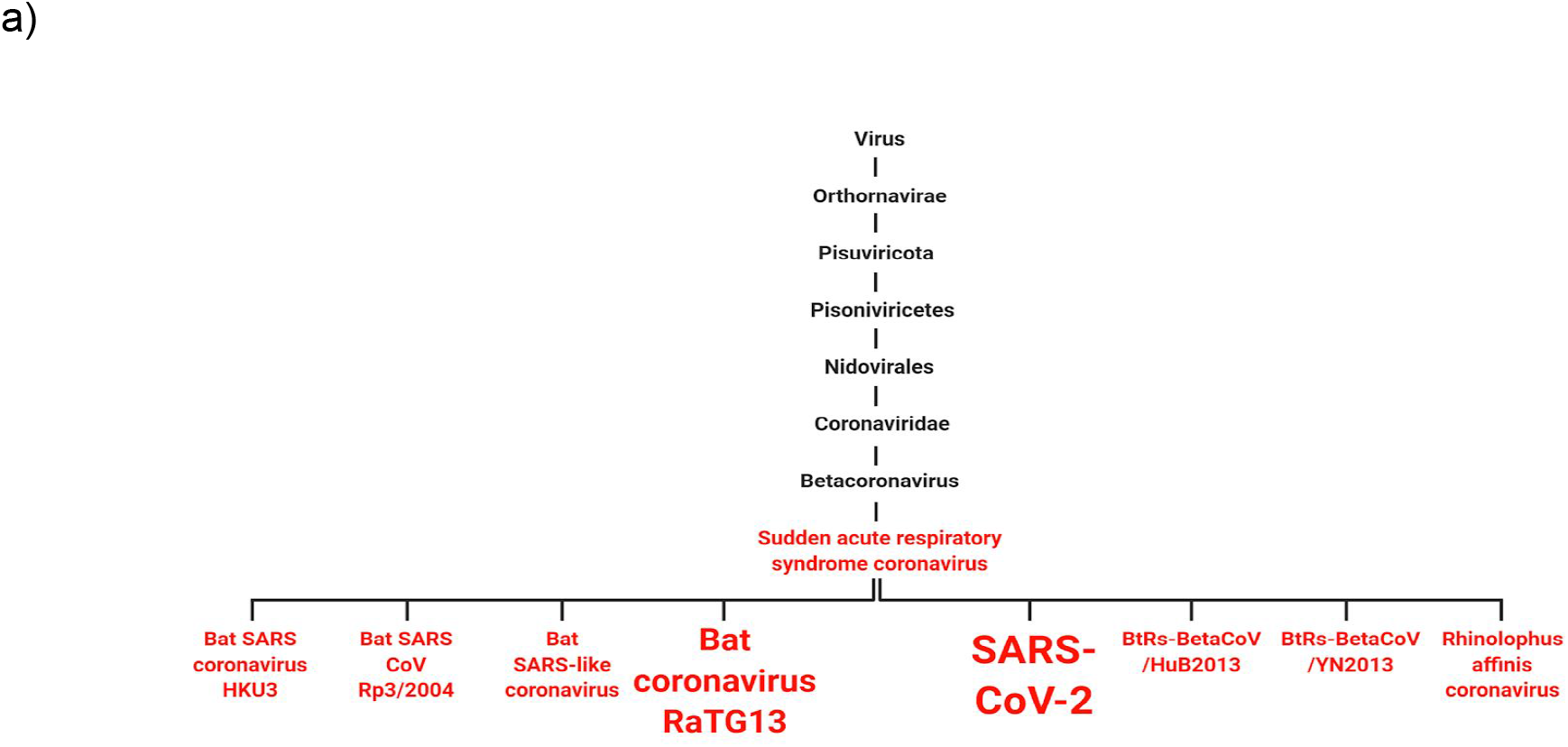
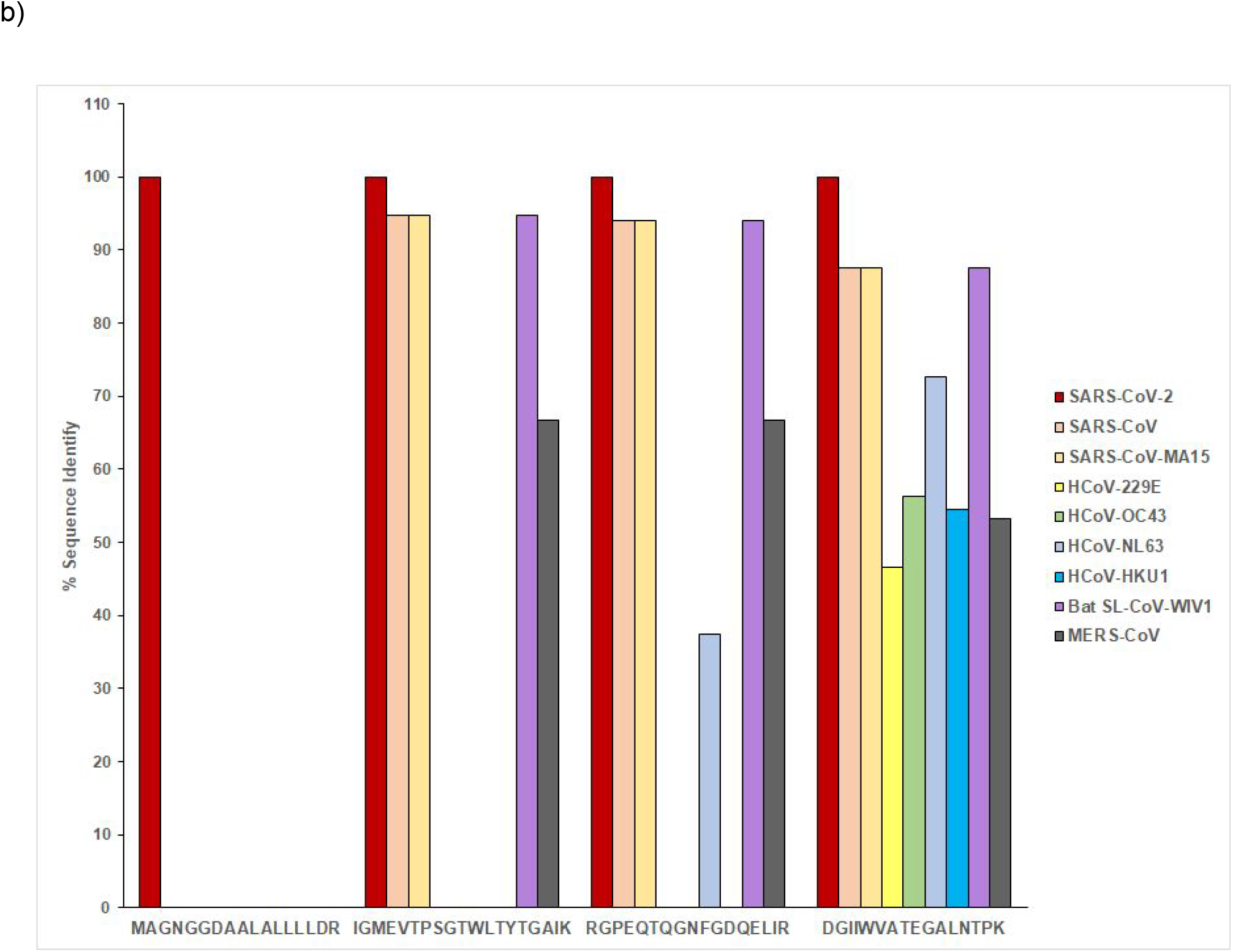
Specificity of target peptides as for coronaviruses and for SARS-CoV-2 a) MetaTryp taxonomic analysis of the 4 most consistently found peptides. Coronaviruses with matches to peptides are highlighted in red and font size is correlated with the number of peptides that show a match in that coronavirus. b) Sequence identity of peptides that show BLAST-P alignment with viral nucleocapsid protein

## Discussion and Conclusions

Bottom-up proteomics has been used to characterize tumors in biopsied breast cancer tissues^42, 43^, to explore the phenotypic changes that occur with opportunistic fungal infections in HIV/AIDS patients^44^, and even differentiate between COVID-19 patients at differing WHO severity grades^45^. While these experiments effectively measure the phenotype of patients to infer a disease state, direct detection of proteins using targeted MS-based methods (SRM) from disease organisms can be used as a diagnostic assay for diseases. For these, it is critical that the most reliable peptides, specific to the protein of interest, are determined.

The pressing nature of the COVID-19 pandemic presents an opportunity for the use of targeted MS-based proteomics to supplement conventional RT-qPCR diagnostic procedures^11^ to mitigate the false negatives inherent in the detection of viral RNA^46^, along with other advantages of direct detection of peptides, such as chemical stability of the target molecules. Ideally, direct detection of diagnostic peptides would be achieved in samples easily collected in the clinic using non-invasive methods. While many labs have begun proteomic analysis of samples to identify SARS-CoV-2 infection in both in-vitro models and clinical samples, the development of targeted assays based on this work requires preliminary work to determine those peptides which are most reliably detected and most specific for unambiguous diagnosis of infection. To mitigate this and establish the best targets possible for a SARS-CoV-2 clinical proteomics assay, we identified detectable SARS-CoV-2 peptides using Galaxy-based workflows. To narrow this list down to the most confident and reliably detected peptides, we then utilized a bioinformatics workflow built around the PepQuery search engine. Developed by Wen et al.^30^, this search engine interrogates raw mass spectrometry data for spectral matches to pre-chosen peptide sequences of interest and compares these matched spectra to reference proteomes to see whether the peptide of interest is a better match to the data than any reference peptide, scoring the peptide match much faster and with much less processing power needed than a conventional sequence database search. By using PepQuery on peptides that have already been designated as potential matches, we can utilize the increased statistical power of using multiple peptide search engines^47^ common to many proteomics software suites on a much faster time scale. Using this as well as other tools available in the Galaxy platform (e.g. the MVP and Lorikeet tools for MS/MS visualization) we were able to interrogate publicly available data to ascertain the most reliable peptides for detecting SARS-CoV-2.

In the two oro/nasopharyngeal datasets and gargled saline dataset we examined, we found 75 peptides within the original list of 639 detected peptides that showed a high-confidence match to SARS-CoV-2 proteins over human proteins or other coronavirus proteins, suggesting that the unambiguous detection of SARS-CoV-2 in patients using proteomics technology is theoretically possible. These peptides were found in proteins throughout the viral particle (**Figure 3**), with more structural protein peptides detected than replication proteins. It was observed that the datasets stemming from the clinical samples had noticeably fewer peptides validated in them compared to those from *in vitro* experiments; this is potentially due to larger amounts of material, the differential abundance of host proteins in clinical samples compared with cultured samples^48^, and the lack of viral clearance from cultured cells^49^. Of these, manual annotation found that 16 peptides could be truly said to have good quality MS/MS spectra, based on our thresholds for PSM quality and annotation.

From the 16 validated peptides with high-quality spectra, 11 peptides also were known to be high-confidence matches in PepQuery. From these we chose 4 peptides that had high-confidence matches in PepQuery, were consistently seen in clinical samples, and were unique to SARS-CoV-2, making them the best candidates for diagnosis of COVID-19 using targeted MS-based methods. It is notable that these are all found within the nucleocapsid phosphoprotein, or N-protein. The nucleocapsid phosphoprotein is common to coronaviruses and serves to complex with and stabilize the viral RNA genome and package it into the viral particle^50, 51^. The viral ribonucleoprotein complex of N-protein and gRNA is localized beneath the matrix proteins (M-proteins) and spike proteins (S-proteins) that make up the capsid surface^52, 53^. As many copies of N-protein are needed to stabilize the viral RNA, the N-protein is thought to be one of the most abundant proteins in the assembled SARS-CoV-2 viral particle^54^; analysis of SARS-CoV transcript levels in infected cells show the N-protein to be the most abundant RNA-based sub-genome within the cell^55^. Taken together, these phenomena explain the prominence of N-protein peptides across the proteomic datasets we examined. As the N-protein is a frequent amplification target for RT-qPCR assays as per FDA guidelines for diagnosis^56^, we believe that our results are complementary to current protocols in screening for and diagnosis of COVID-19.

In addition to upper respiratory tract clinical samples, we profiled datasets derived from deep within the respiratory tract, comprising a dataset derived from COVID-19 patient lung biopsies as well as a separate dataset of bronchoalveolar lavage fluid (BALF) samples from COVID-19 patients; we analyzed these MS-data against our 639 peptide panel to determine whether our methodology was suitable for SARS-CoV-2 detection in these samples. We found a lack of high-confidence peptides with high-quality spectral annotations in these samples, with only a single MS run from the PXD022085 sample yielding the peptide FLALCADSIIIGGAK which was not found in the datasets derived from higher up in the respiratory tract. Our results would suggest that samples collected using invasive methods (biopsy, lung fluid extraction), in addition to being taxing on the patients to collect, demonstrate insufficient concentrations of viral particles to be robustly detected using MS-based methods and the workflows presented here; Conversely, our results also suggest that samples collected using minimally invasive methods from the upper respiratory tract (oropharyngeal/nasopharyngeal swabs and gargling samples) could be suitable for reliable detection of the SARS-CoV-2 virus targeting the high-confidence peptides we identify here – offering an optimal method for high-throughput diagnosis of infection.

While we believe the peptides presented here constitute promising targets for COVID-19 diagnosis, there are further experiments required to establish targeted proteomics as a viable methodology for detection of SARS-CoV-2 infection in the clinical setting. The limits of detection of these peptides need to be reliably established in larger numbers of human samples collected in the clinic to determine the minimal number of viral particles that can be detected. This could help determine the optimal sample type and procedure for collection to ensure reliable results. Two of the four most reliably detected peptides also contain methionines, and one carries a mis-cleaved tryptic site – all potential factors affecting accurate quantification by SRM that require further testing for developing robust targeted detection methods. In addition, proteomic analysis of samples collected at different stages of SARS-CoV-2 infection should be performed to determine viability of targeted proteomics for detection during the full life cycle of infection. Finally, the sample processing that accompanies bottom-up proteomics^57^ should be optimized to be performed on a rapid time scale. Most conventional bottom-up proteomics experiments utilize trypsin digestions which occur overnight with incubation at 37°C, meaning a single sample would have to be processed and analyzed over the course of two days; this would have to be significantly reduced as the conventional 24-48 hour complete turnaround of RT-qPCR assays is being decreased through the use of strategies such as direct RT-qPCR^12^, RT-LAMP^13^, and CRISPR-based amplification strategies^14, 58, 59^. The turnaround time of clinical proteomics can potentially be decreased for individual samples using modified or alternative protein digestion enzymes with higher rates of reactivity^60^; in addition, automation of clinical proteomics technology can provide reproducible, robust analyses of patient samples^61, 62^.

In addition to peptides derived empirically from clinical and *in vitro* datasets, we also included theoretical SARS-CoV-2 peptides predicted bioinformatically by Orsburn *et al*. in our panel for validation; in doing so we were able to validate eight peptides in both clinical and *in vitro* datasets. It is worth noting, however, that of these eight peptides only two peptides were observed to have good quality PSMs in the clinical data, supporting the need for caution in accepting peptide identifications. The validation workflow presented here was also able to identify peptides in MS data which conventional unbiased algorithms, such as our database search workflow presented in Figure 2b, are unable to identify; this may be of use in the analysis of complex patient and environmental mass spectrometry data collected for alternate purposes in the detection of SARS-CoV-2 under various conditions. With the increased surveillance for genomic variants of the SARS-CoV-2 virus^63^, we also anticipate the utility of the peptide validation workflow in detecting variants when the variant peptide sequences are added to the 639-peptide panel.

In conclusion, we interrogated multiple proteomic datasets from COVID-19 patients and *in vitro* experiments using bioinformatics workflows in order to determine which peptides from SARS-CoV-2 would make suitable targets for a clinical proteomics assay and which would make poor targets, potentially resulting in false negatives. Through our analyses we found that of the 639 peptides that are readily detected across all samples, 87 of these were found to have a significant match within the SARS-CoV-2 proteome than that within the human proteome or other coronavirus proteomes. These peptides were narrowed down to 4 high-confidence peptides with excellent quality spectral annotations matching these sequences, found across most of the upper-respiratory tract clinical datasets analyzed in this study which we believe would be ideal candidates for diagnosis of COVID-19 via targeted proteomics.

Finally, the workflows employed here for peptide identification and validation are well-documented, open-source, and hosted on the Galaxy Europe platform where they can be edited, modified, or interfaced with other relevant bioinformatics tools to aid in analysis of proteomics data. We believe that these workflows will be extremely useful for developing and evaluating diagnostic proteomic assays for pathogenic infections^64^, protein sequence mutations arising from DNA damage, oncogenesis, etc., beyond the current application to COVID-19.

## Data Availability

The data used herein is available at the Proteomics Identification Database (PRIDE). Our workflows are documented in the shared drive, access to which is available upon request..

https://drive.google.com/drive/folders/1vYglcdCzZle2HjLCIxcxR7lU-5fM_Vbf?usp=sharing

## Declarations

## Acknowledgements

We would like to thank the European Galaxy team and ELIXIR-Europe for the help in the support during Galaxy implementation and hosting the COVID-19 project webpage. We greatly appreciate inputs and help in data organization by Ms. Emma Leith.

## Funding

We acknowledge funding for this work from the grant National Cancer Institute - Informatics Technology for Cancer Research (NCI-ITCR) grant 1U24CA199347 to T.J.G. The European Galaxy server that was used for data analysis is in part funded by Collaborative Research Centre 992 Medical Epigenetics (DFG grant SFB 992/1 2012) and German Federal Ministry of Education and Research (BMBF grants 031 A538A/A538C RBC, 031L0101B/031L0101C de.NBI-epi, 031L0106 de.STAIR (de.NBI)). Andrew T. Rajczewski was supported by Biotechnology Training Grant: NIH T32GM008347.

## Author contributions

ATR and SM drafted the original manuscript, performed data analysis and revised the final manuscript. DDAN contributed to data analysis. BAG, JEJ and TMG helped in software implementation and reviewing the manuscript. TJG critically reviewed the manuscript and provided supervision and funding for the project. PDJ conceptualized and supervised the project along with data analysis and manuscript preparation and review. All authors read and approved the final manuscript.

## Consent for publication

Not applicable

## Ethics approval and consent to participate

As this study utilized open-source mass spectrometry data of anonymous human subjects from previous studies, no ethics approval or consent to participate was needed.

## Competing interests

The authors declare that they have no competing interests

## Data and Workflow Availability

All the data and workflows used in this study are available here:

http://doi.org/10.5281/zenodo.4566595

| Data | Lab | Proteome exchange ID | Data availability |
| --- | --- | --- | --- |
| Cell culture | Gouveia *et al.* | PXD018804 | https://covid19.galaxyproject.org/proteomics/PXD018804/ |
|  | Grenga *et al.* | PXD018594 | https://covid19.galaxyproject.org/proteomics/PXD018594/ |
|  | Matthews *et al.* | PXD018241 | https://covid19.galaxyproject.org/proteomics/PXD018241/ |
| Clinical samples | Cardozo *et al.* | PXD021328 | https://covid19.galaxyproject.org/proteomics/PXD021328/ |
|  | Ihling *et al.* | PXD019423 | https://covid19.galaxyproject.org/proteomics/PXD019423/ |
|  | Rivera *et al.* | PXD020394 | https://covid19.galaxyproject.org/proteomics/PXD020394/ |

## Notes

### Competing Interest Statement

The authors have declared no competing interest.

### Author Declarations

This work was performed on open-source mass spectrometry data, and as such no IRB approval was required.

### Summary of Updates

An updated version of this manuscript with significant rewording was created, in addition to supplemental files added.

