## Supplemental Data for "A rigorous evaluation of optimal peptide targets for MS-based clinical diagnostics of Coronavirus Disease 2019 (COVID-19)"

**Supplemental Methods:** Search parameters for SearchGUI for the analysis of cell culture and clinical datasets in Galaxy. Parameters were based upon data analysis protocols detailed in the original publications.

| | Cell culture datasets | | | Clinical datasets | | |
|---|---|---|---|---|---|---|
| *Search Parameters* | **PXD018804** | **PXD018594** | **PXD018241** | **PXD021328** | **PXD019423** | **PXD020394** |
| **Algorithms** | X!Tandem, MS-GF+, OMSSA, Comet | X!Tandem, MS-GF+, OMSSA | X!Tandem, MS-GF+, OMSSA | X!Tandem, MS-GF+, OMSSA | X!Tandem, MS-GF+, OMSSA | X!Tandem, MS-GF+, OMSSA |
| **Digestion Enzymes** | Trypsin | Trypsin | Trypsin | Trypsin | Trypsin | Trypsin |
| **Missed cleavages** | 2 | 2 | 2 | 2 | 2 | 2 |
| **Precursor Ion Tolerance** | 5 ppm | 5 ppm | 10 ppm | 10 ppm | 10 ppm | 10 ppm |
| **Fragment Tolerance** | 0.02 Da | 0.02 Da | 0.6 Da | 10 ppm | 0.05 Da | 0.05 Da |
| **Minimum Charge** | 2 | 2 | 2 | 2 | 2 | 2 |
| **Maximum Charge** | 4 | 6 | 6 | 6 | 6 | 6 |
| **Fixed Modifications** | Carbamidomethylation of C | Carbamidomethylation of C | Carbamidomethylation of C | - | Carbamidomethylation of C | Carbamidomethylation of C |
| **Variable Modifications** | Deamidation of N, Deamidation of Q, Oxidation of M | Deamidation of N, Deamidation of Q, Oxidation of M | Acetylation of protein N-term Oxidation of M | Acetylation of protein N-term, Oxidation of M | Deamidation of N, Oxidation of M | Oxidation of M |
| **Minimum Peptide Length** | 6 | 8 | 8 | 8 | 8 | 8 |
| **Maximum Peptide Length** | 30 | 60 | 60 | 60 | 60 | 60 |
| **Maximum Precursor Error** | 10 ppm | 5 ppm | 10 ppm | 10 ppm | 10 ppm | 10 ppm |

Parameters for the PepQuery search engine for the validation of clinical datasets in Galaxy.

| | Clinical datasets | | | | |
|---|---|---|---|---|---|
| *PepQuery Parameters* | **PXD021328** | **PXD019423** | **PXD020394** | **PXD022085** | **PXD018094** |
| Fixed modification(s) | | Carbamidomethylation of C | Carbamidomethylation of C | Carbamidomethylation of C | Carbamidomethylation of C |
| Variable modification(s) | Oxidation of M | Oxidation of M | Oxidation of M | Oxidation of M, Acetylation of peptide N-terminus | Oxidation of M, Acetylation of peptide N-terminus |
| Max Modifications | 3 | 3 | 3 | 3 | 3 |
| Unrestricted modification? | True | True | True | True | True |
| Amino Acid substitutions? | True | True | True | True | True |
| Precursor tolerance | 10 | 10 | 10 | 10 | 10 |
| Precursor unit | ppm | ppm | ppm | ppm | ppm |
| Product tolerance (Da) | 0.05 | 0.05 | 0.05 | 0.05 | 0.05 |
| Digestion enzyme | Trypsin | Trypsin | Trypsin | Trypsin | Trypsin |
| Max missed cleavages | 2 | 2 | 2 | 2 | 2 |
| Fragmentation | CID/HCD | CID/HCD | CID/HCD | CID/HCD | CID/HCD |
| Scoring method | HyperScore | HyperScore | HyperScore | HyperScore | HyperScore |
| Max charge | 6 | 6 | 6 | 6 | 6 |
| Minimum charge | 2 | 2 | 2 | 2 | 2 |
| Minimum peaks | 10 | 10 | 10 | 10 | 10 |
| Minimum score | 12 | 12 | 12 | 12 | 12 |
| Max peptide length | 45 | 45 | 45 | 45 | 45 |
| Number of random peptides | 1000 | 1000 | 1000 | 1000 | 1000 |
| "Spectrum_file" column? | True | True | True | True | True |

**Supplementary Figure 1. Alignment of 639 peptide panel to viral proteins from SARS-CoV-2.** Peptides detected from patient and cell culture datasets were aligned to the SARS-CoV-2 proteome. Proteins were colored in terms of their classification as structural proteins (green), non-structural proteins (maroon), or open reading frames (blue).

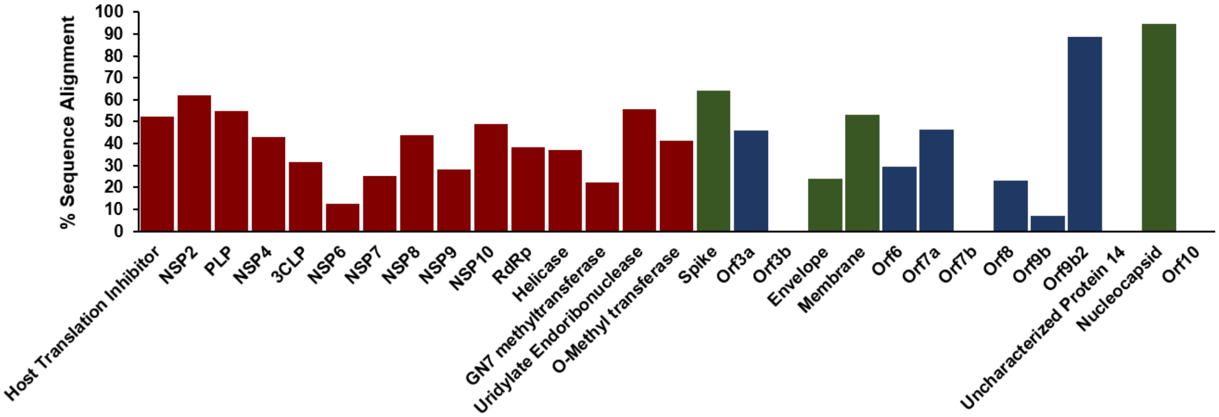

**Supplementary Figure 2. List of validated peptide spectral matches in the oro-pharyngeal and naso-pahryngeal mass spectrometry dataset (PXD020394).** Bar diagram above enlists the peptide-spectral matches after running the validation workflow for 639 SARS-CoV-2 peptide-panel against the five COVID-19 positive patient samples (with replicates) and five COVID-19 negative patient samples (with replicates). Several SARS-CoV-2 peptides were detected in COVID-19 positive samples (See samples labeled 'POS") and only two peptides were detected in two of the negative samples (NEG5 Rep1 and NEG2 Rep2). The SARS-CoV-2 peptides detected in COVID-19 negative samples did not meet the threshold of acceptable spectral quality in subsequent spectral validation using Lorikeet (See supplementary figures for Lorikeet analysis).

**ORO-PHARYNGEAL AND NASO-PHARYNGEAL SAMPLE MS DATASETS (PXD020394)**

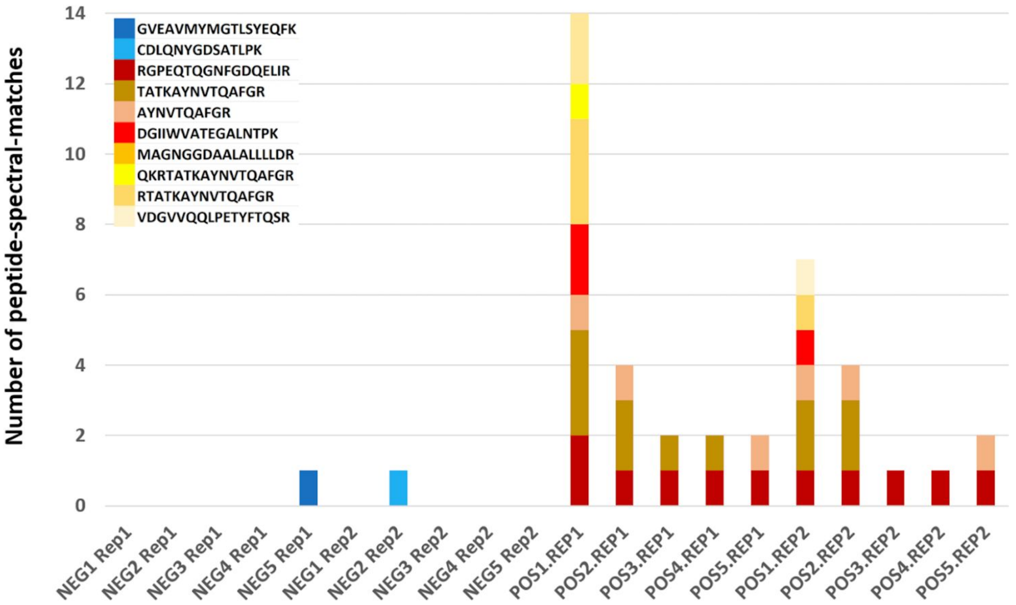

**Supplementary Figure 3. Spectra of validated SARS-CoV-2 peptides.** MS/MS spectra of 75 peptides which passed bioinformatic validation and were assessed manually. Spectra were visualized using the Multi-omics Visualization Platform and Proteomics Data Viewer platforms. Peptides were considered to have high-quality spectra if the spectra contained b- and/or y-ion series with at least three continuous product ions and if the product ions were at least three-fold more intense than noise.

**CDLQNYGDSATLPK**

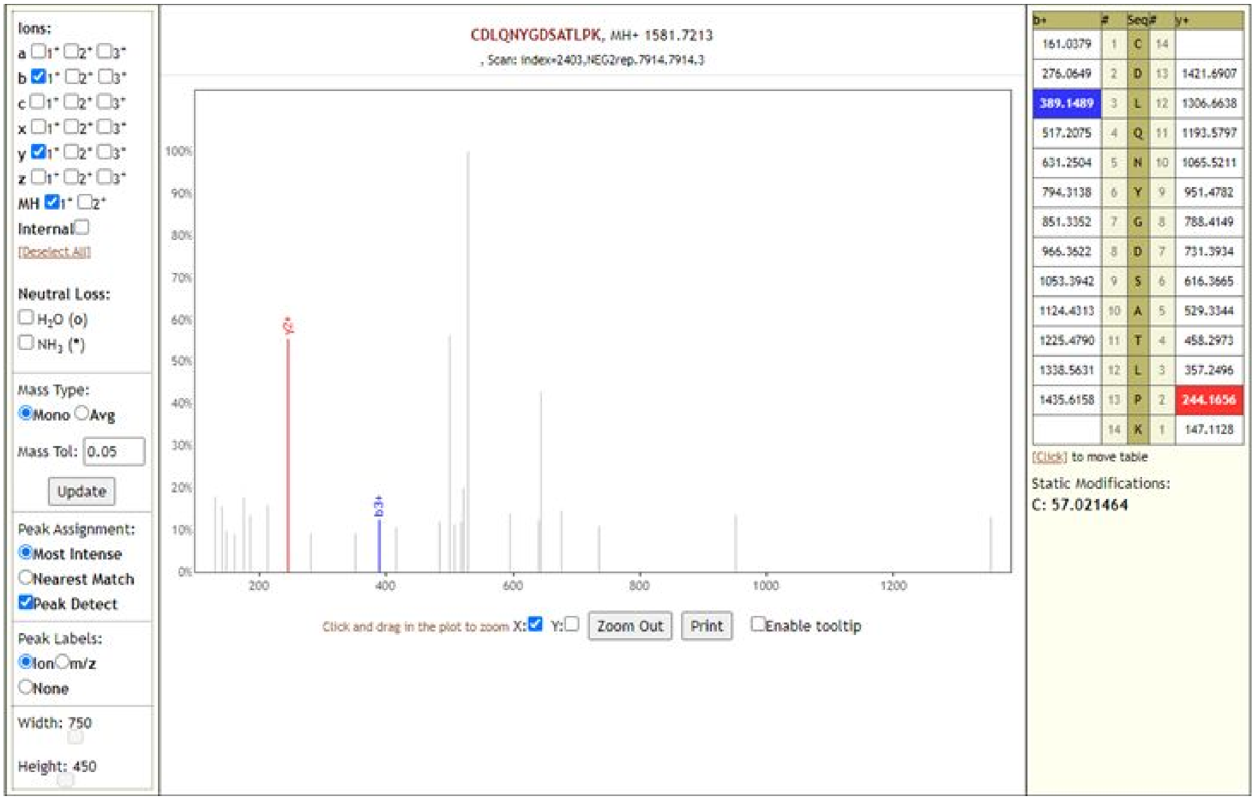

**GVEAVMYMGTLSYEQFK**

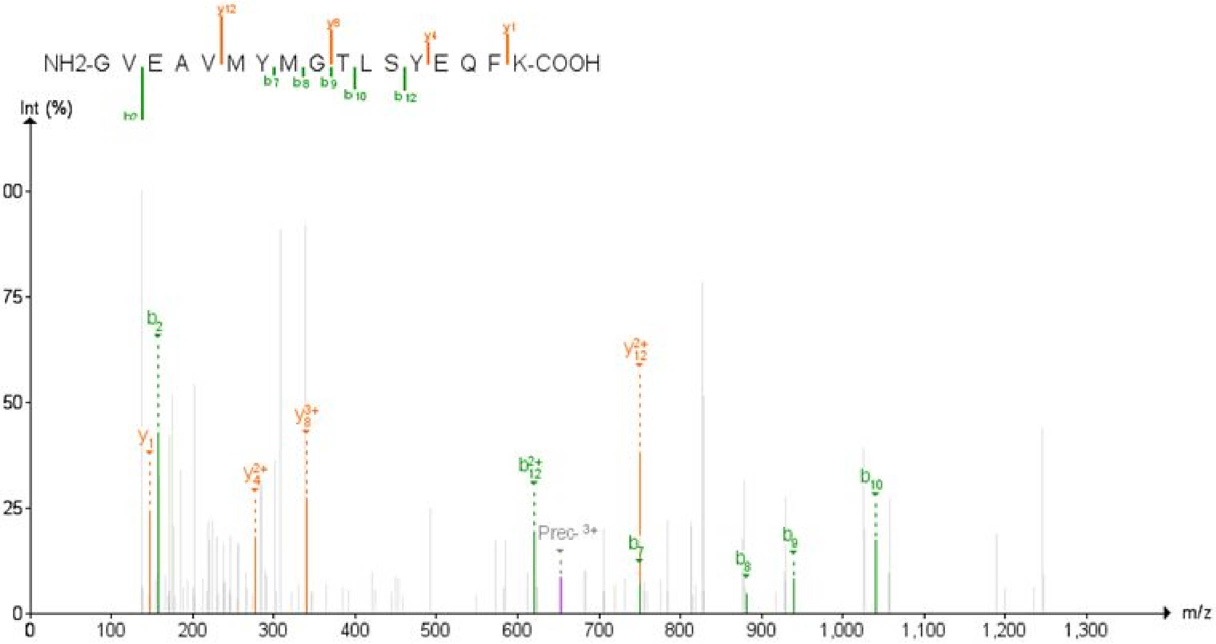

**AYNVTQAFGR**

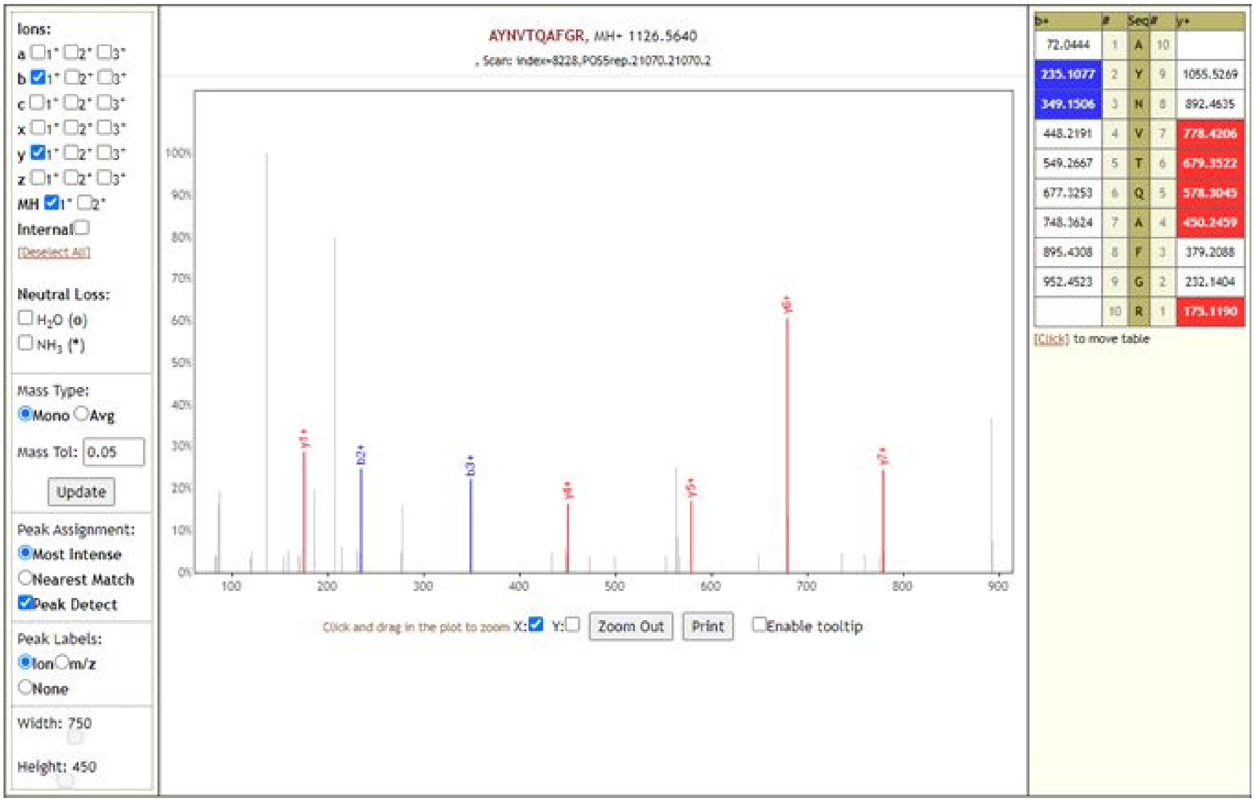

### MAGNGGDAALALLLLDR

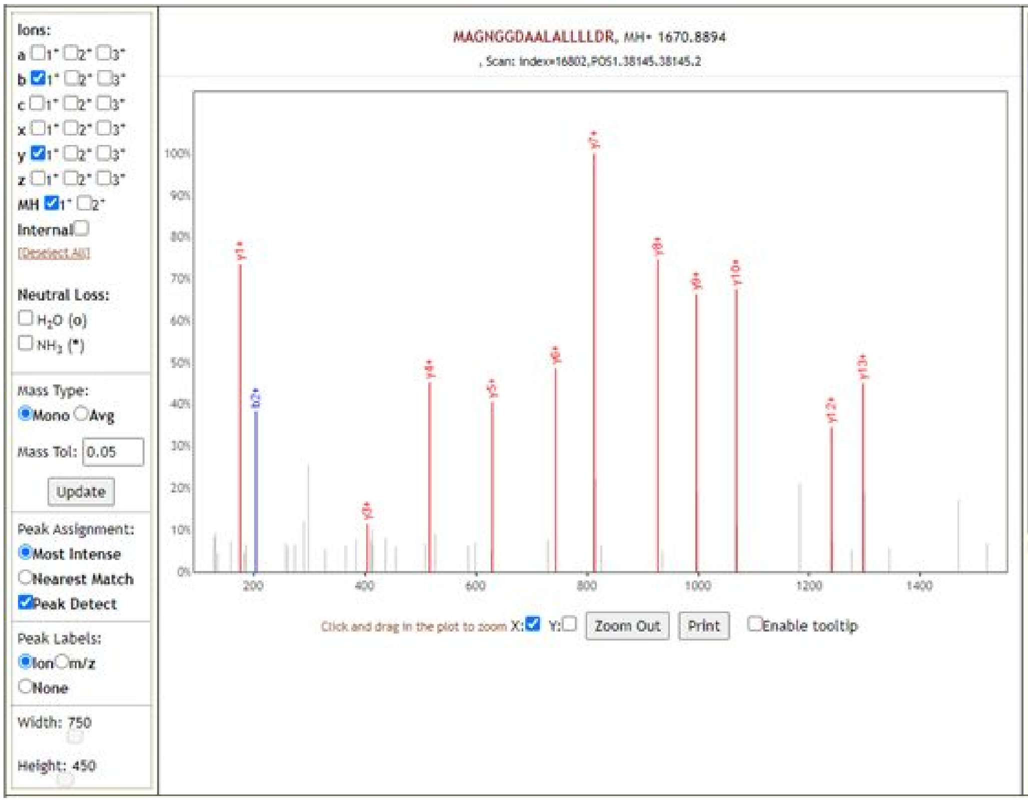

| b+ | # | Seq | # | y+ |
|---|---|---|---|---|
| 132.0478 | 1 | M | 17 | |
| 203.0849 | 2 | A | 16 | 1539.8489 |
| 260.1063 | 3 | G | 15 | 1468.8118 |
| 374.1493 | 4 | N | 14 | 1411.7904 |
| 431.1707 | 5 | G | 13 | 1297.7474 |
| 488.1922 | 6 | G | 12 | 1240.7260 |
| 603.2191 | 7 | D | 11 | 1183.7045 |
| 674.2562 | 8 | A | 10 | 1068.6776 |
| 745.2934 | 9 | A | 9 | 997.6404 |
| 858.3774 | 10 | L | 8 | 926.6033 |
| 929.4145 | 11 | A | 7 | 813.5193 |
| 1042.4986 | 12 | L | 6 | 742.4822 |
| 1155.5827 | 13 | L | 5 | 629.3981 |
| 1268.6667 | 14 | L | 4 | 516.3140 |
| 1381.7508 | 15 | L | 3 | 403.2300 |
| 1496.7777 | 16 | D | 2 | 290.1459 |
| | 17 | R | 1 | 175.1190 |

[Click] to move table

### RGPEQTQGNFGDQELIR

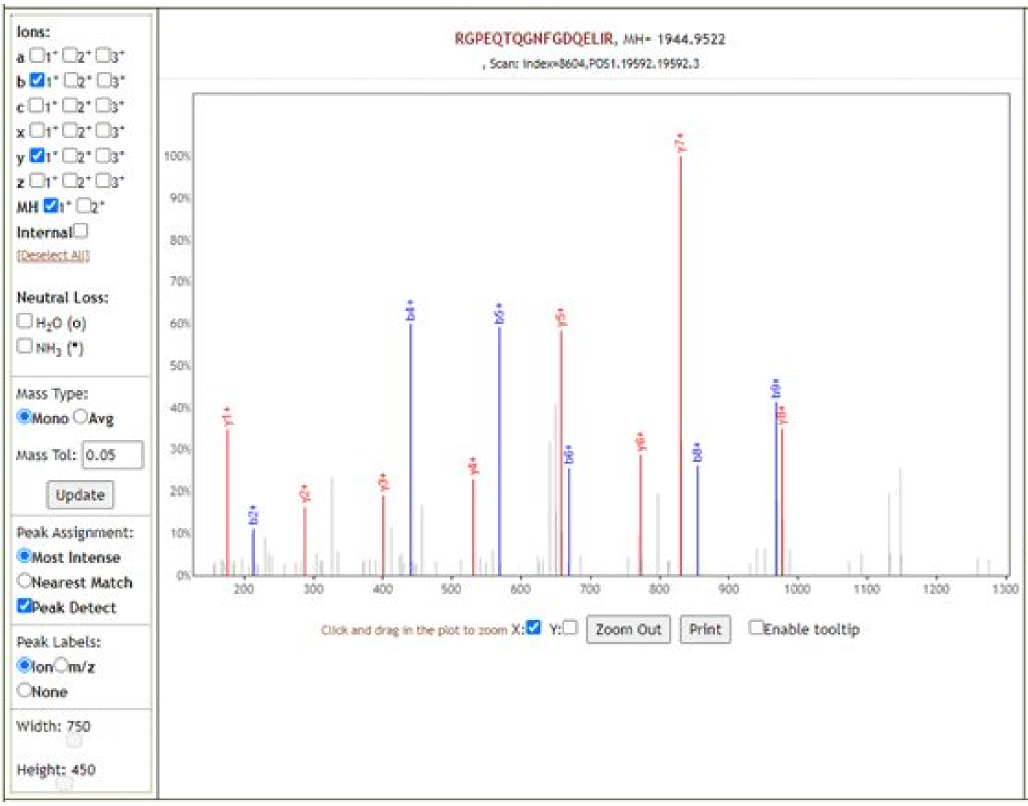

| b+ | # | Seq | # | y+ |
|---|---|---|---|---|
| 157.1084 | 1 | R | 17 | |
| 214.1299 | 2 | G | 16 | 1788.8511 |
| 311.1826 | 3 | P | 15 | 1731.8297 |
| 440.2252 | 4 | E | 14 | 1634.7769 |
| 568.2838 | 5 | Q | 13 | 1505.7343 |
| 669.3315 | 6 | T | 12 | 1377.6757 |
| 797.3900 | 7 | Q | 11 | 1276.6280 |
| 854.4115 | 8 | G | 10 | 1148.5695 |
| 968.4544 | 9 | N | 9 | 1091.5480 |
| 1115.5228 | 10 | F | 8 | 977.5051 |
| 1172.5443 | 11 | G | 7 | 830.4367 |
| 1287.5713 | 12 | D | 6 | 773.4152 |
| 1415.6298 | 13 | Q | 5 | 658.3883 |
| 1544.6724 | 14 | E | 4 | 530.3297 |
| 1657.7565 | 15 | L | 3 | 401.2871 |
| 1770.8406 | 16 | I | 2 | 288.2030 |
| | 17 | R | 1 | 175.1190 |

[Click] to move table

**DGIIWVATEGALNTPK**

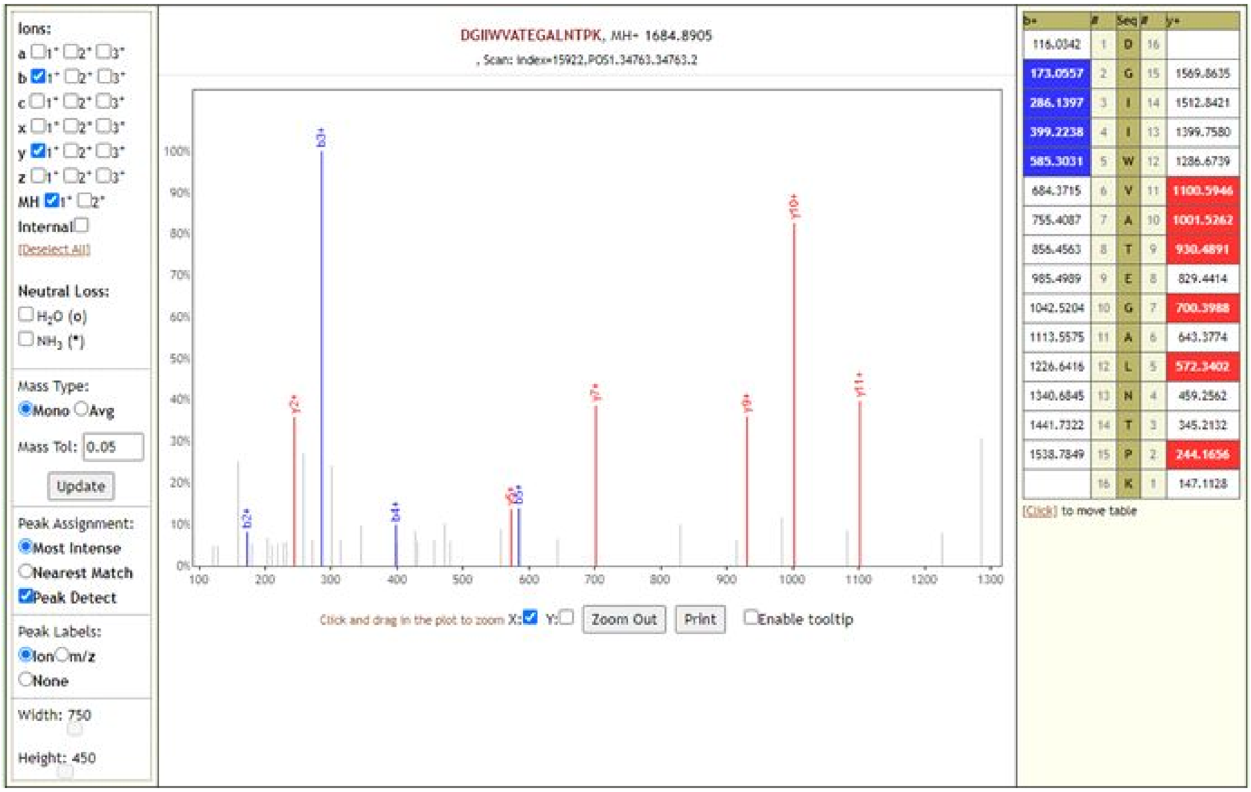

**NPANNAAIVLQLPQGTTLPK**

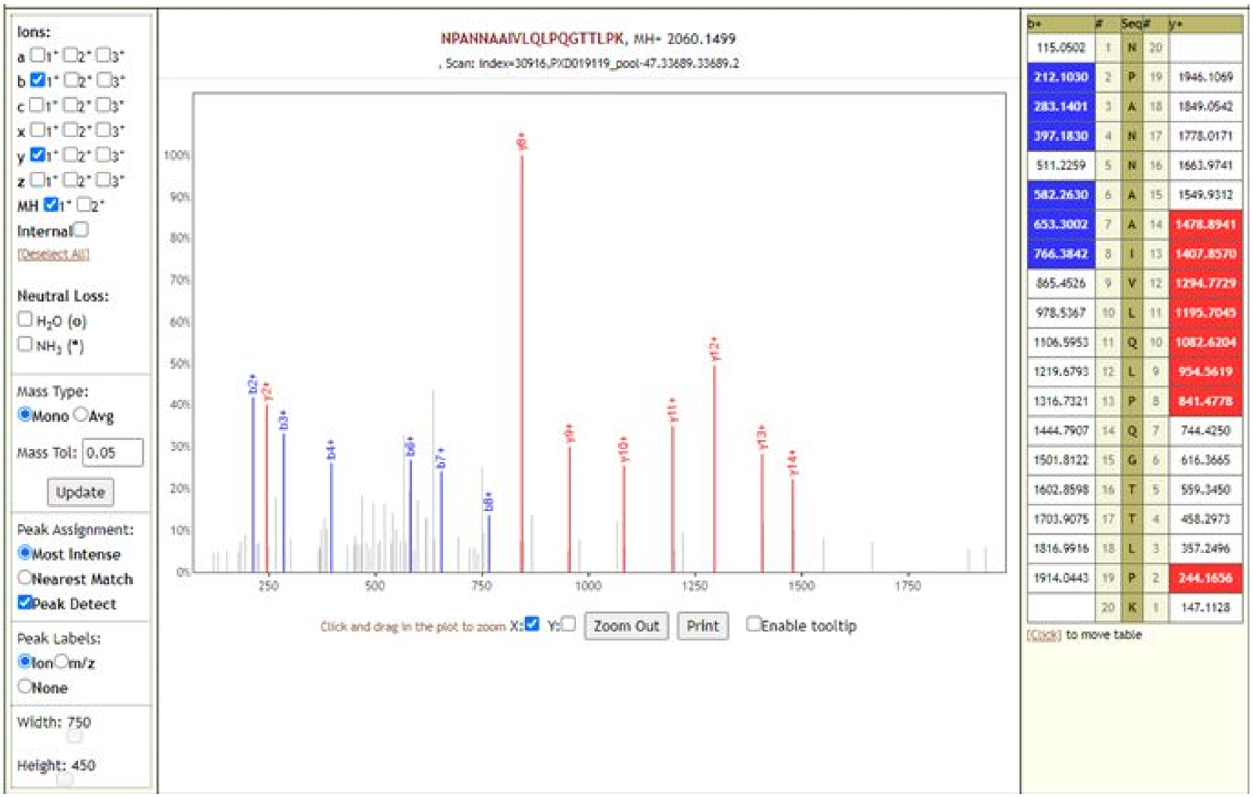

**RPQGLPNNTASWFTALTQHGKEDLKFPR**

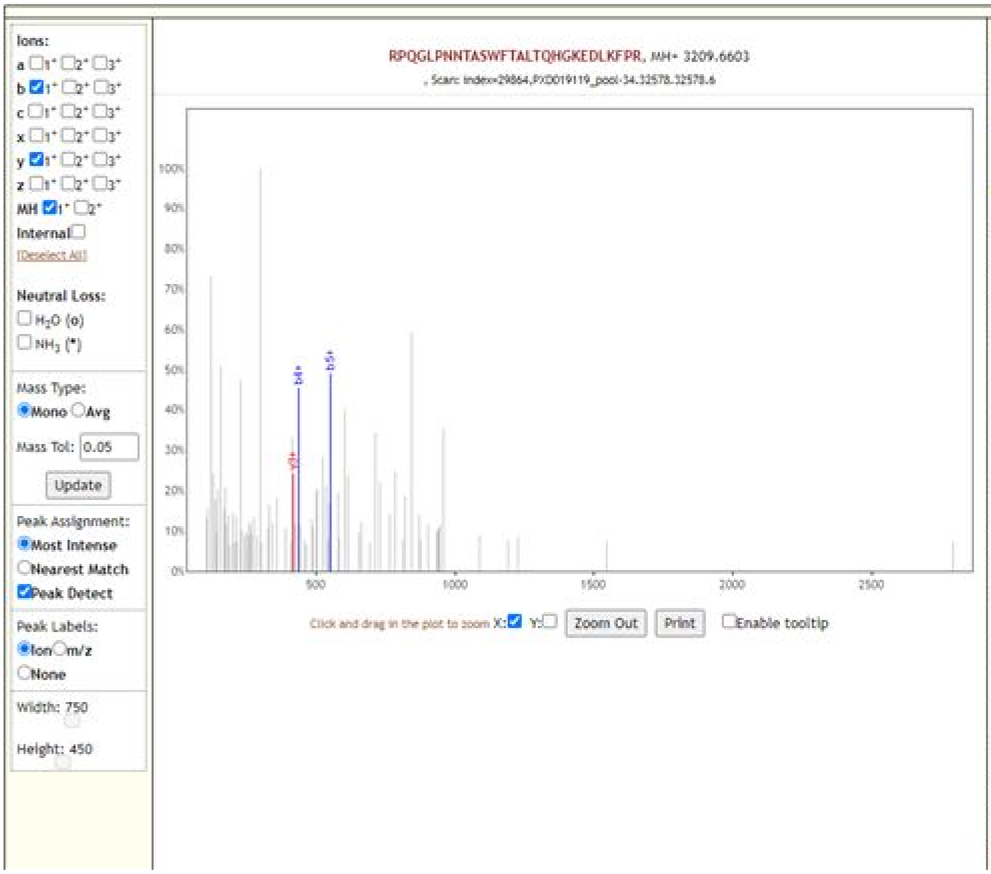

| b+ | # | Seq | # | y+ |
|---|---|---|---|---|
| 157.1084 | 1 | R | 28 | |
| 254.1612 | 2 | P | 27 | 3053.5592 |
| 382.2197 | 3 | Q | 26 | 2956.5064 |
| 439.2412 | 4 | G | 25 | 2828.4478 |
| 552.3253 | 5 | L | 24 | 2771.4264 |
| 649.3780 | 6 | P | 23 | 2658.3423 |
| 763.4209 | 7 | N | 22 | 2561.2895 |
| 877.4639 | 8 | N | 21 | 2447.2466 |
| 978.5116 | 9 | T | 20 | 2333.2037 |
| 1049.5487 | 10 | A | 19 | 2232.1560 |
| 1136.5807 | 11 | S | 18 | 2161.1189 |
| 1322.6600 | 12 | W | 17 | 2074.0869 |
| 1469.7284 | 13 | F | 16 | 1888.0076 |
| 1570.7761 | 14 | T | 15 | 1740.9391 |
| 1641.8132 | 15 | A | 14 | 1639.8915 |
| 1754.8973 | 16 | L | 13 | 1568.8544 |
| 1855.9450 | 17 | T | 12 | 1455.7703 |
| 1984.0035 | 18 | Q | 11 | 1354.7226 |
| 2121.0624 | 19 | H | 10 | 1226.6640 |
| 2178.0839 | 20 | G | 9 | 1089.6051 |
| 2306.1789 | 21 | K | 8 | 1032.5837 |
| 2435.2215 | 22 | E | 7 | 904.4887 |
| 2550.2484 | 23 | D | 6 | 775.4461 |
| 2663.3325 | 24 | L | 5 | 660.4192 |
| 2791.4274 | 25 | K | 4 | 547.3351 |
| 2938.4959 | 26 | F | 3 | 419.2401 |
| 3035.5486 | 27 | P | 2 | 272.1717 |
| | 28 | R | 1 | 175.1190 |

[Click] to move table

**LNTDHSSSSDNIALLVQ**

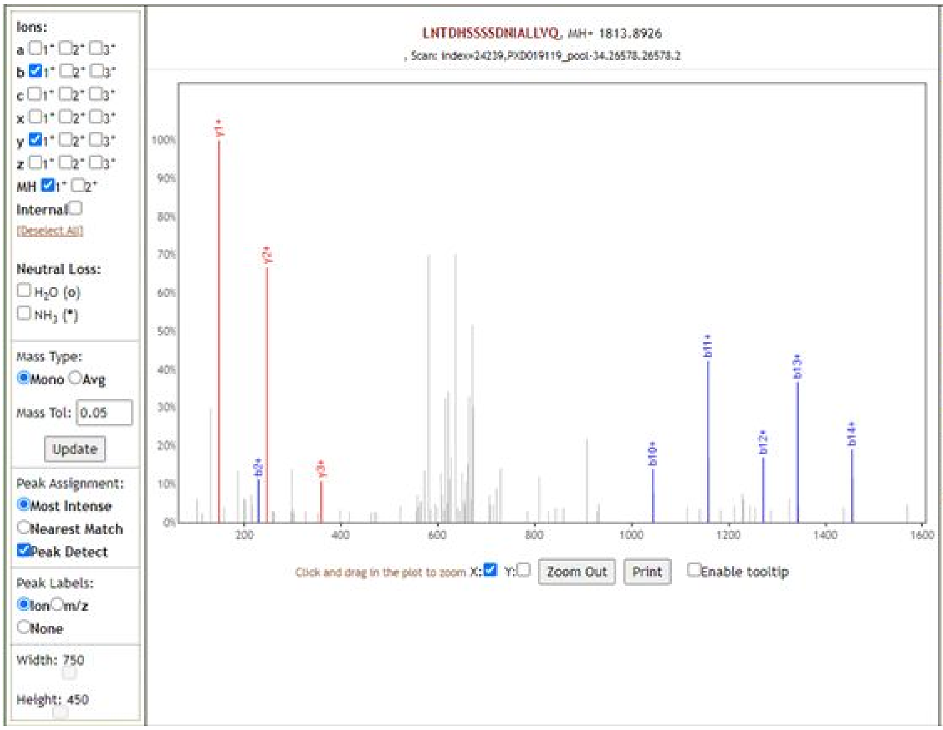

| b+ | # | Seq | # | y+ |
|---|---|---|---|---|
| 114.0913 | 1 | L | 17 | |
| 228.1343 | 2 | N | 16 | 1700.8086 |
| 329.1819 | 3 | T | 15 | 1586.7657 |
| 444.2089 | 4 | D | 14 | 1485.7180 |
| 581.2678 | 5 | H | 13 | 1370.6910 |
| 668.2998 | 6 | S | 12 | 1233.6321 |
| 755.3319 | 7 | S | 11 | 1146.6001 |
| 842.3639 | 8 | S | 10 | 1059.5681 |
| 929.3959 | 9 | S | 9 | 972.5360 |
| 1044.4229 | 10 | D | 8 | 885.5040 |
| 1158.4658 | 11 | N | 7 | 770.4771 |
| 1271.5498 | 12 | I | 6 | 656.4341 |
| 1342.5870 | 13 | A | 5 | 543.3501 |
| 1455.6710 | 14 | L | 4 | 472.3130 |
| 1568.7551 | 15 | L | 3 | 359.2289 |
| 1667.8235 | 16 | V | 2 | 246.1448 |
| | 17 | Q | 1 | 147.0764 |

[Click] to move table

### YLGTGPEAGLPYGANK

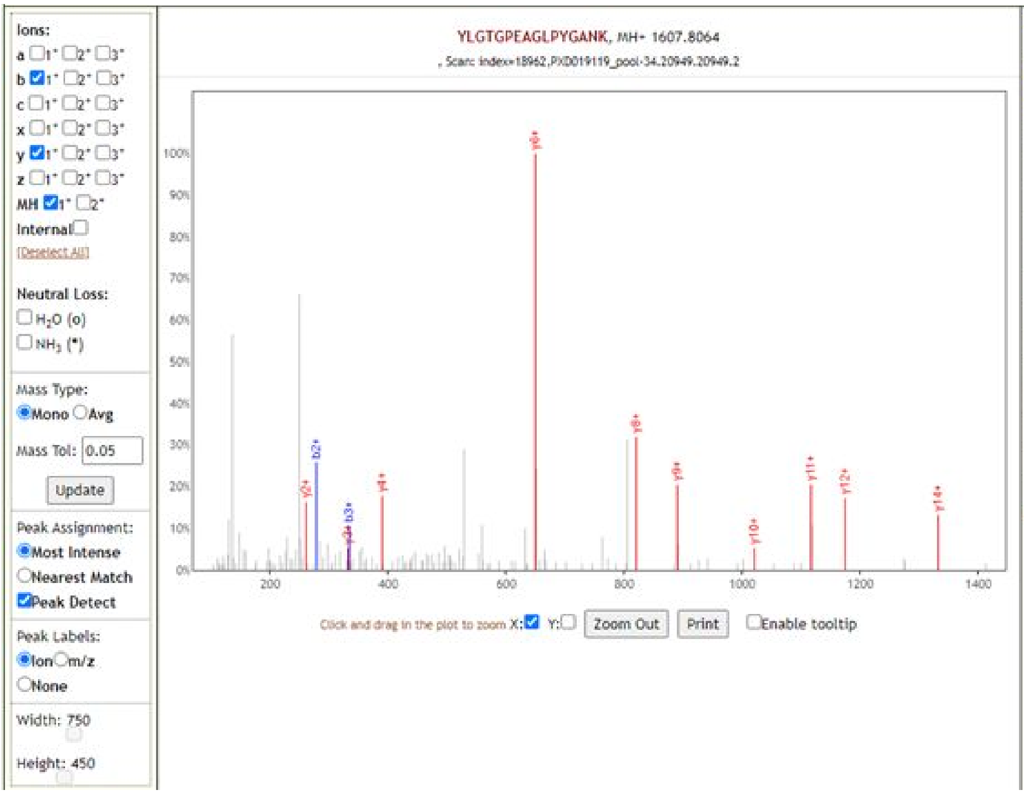

| b+ | # | Seq | # | y+ |
|---|---|---|---|---|
| 164.0706 | 1 | Y | 16 | |
| 277.1547 | 2 | L | 15 | 1444.7431 |
| 334.1761 | 3 | G | 14 | 1331.6590 |
| 435.2238 | 4 | T | 13 | 1274.6375 |
| 492.2453 | 5 | G | 12 | 1173.5899 |
| 589.2980 | 6 | P | 11 | 1116.5684 |
| 718.3406 | 7 | E | 10 | 1019.5156 |
| 789.3777 | 8 | A | 9 | 890.4730 |
| 846.3992 | 9 | G | 8 | 819.4359 |
| 959.4833 | 10 | L | 7 | 762.4145 |
| 1056.5360 | 11 | P | 6 | 649.3304 |
| 1219.5994 | 12 | Y | 5 | 552.2776 |
| 1276.6208 | 13 | G | 4 | 389.2143 |
| 1347.6579 | 14 | A | 3 | 332.1928 |
| 1461.7009 | 15 | N | 2 | 261.1557 |
| | 16 | K | 1 | 147.1128 |

[Click] to move table

### IGMEVTPSGTWLTYTGAIK

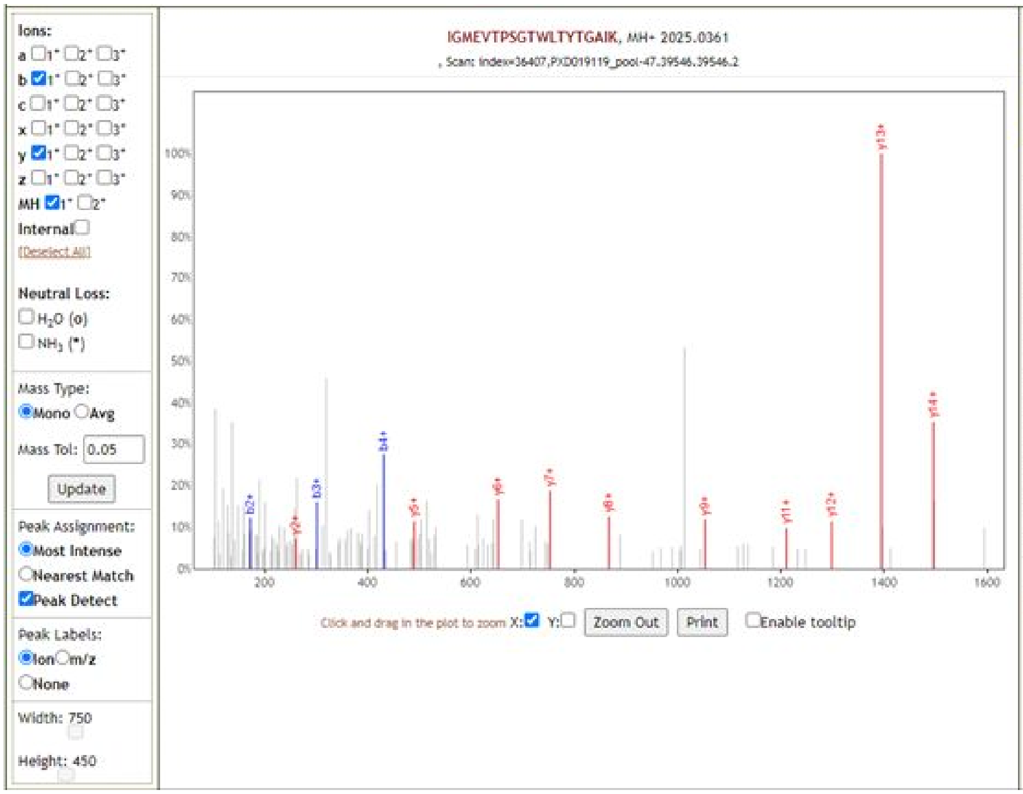

| b+ | # | Seq | # | y+ |
|---|---|---|---|---|
| 114.0913 | 1 | I | 19 | |
| 171.1128 | 2 | G | 18 | 1911.9521 |
| 302.1533 | 3 | M | 17 | 1854.9306 |
| 431.1959 | 4 | E | 16 | 1723.8901 |
| 530.2643 | 5 | V | 15 | 1594.8475 |
| 631.3120 | 6 | T | 14 | 1495.7791 |
| 728.3647 | 7 | P | 13 | 1394.7314 |
| 815.3968 | 8 | S | 12 | 1297.6787 |
| 872.4182 | 9 | G | 11 | 1210.6467 |
| 973.4659 | 10 | T | 10 | 1153.6252 |
| 1159.5452 | 11 | W | 9 | 1052.5775 |
| 1272.6293 | 12 | L | 8 | 866.4982 |
| 1373.6770 | 13 | T | 7 | 753.4141 |
| 1536.7403 | 14 | Y | 6 | 652.3665 |
| 1637.7880 | 15 | T | 5 | 489.3031 |
| 1694.8094 | 16 | G | 4 | 388.2554 |
| 1765.8465 | 17 | A | 3 | 331.2340 |
| 1878.9306 | 18 | I | 2 | 260.1969 |
| | 19 | K | 1 | 147.1128 |

[Click] to move table

**MAGNGGDAALALLLLDRLNQLESK**

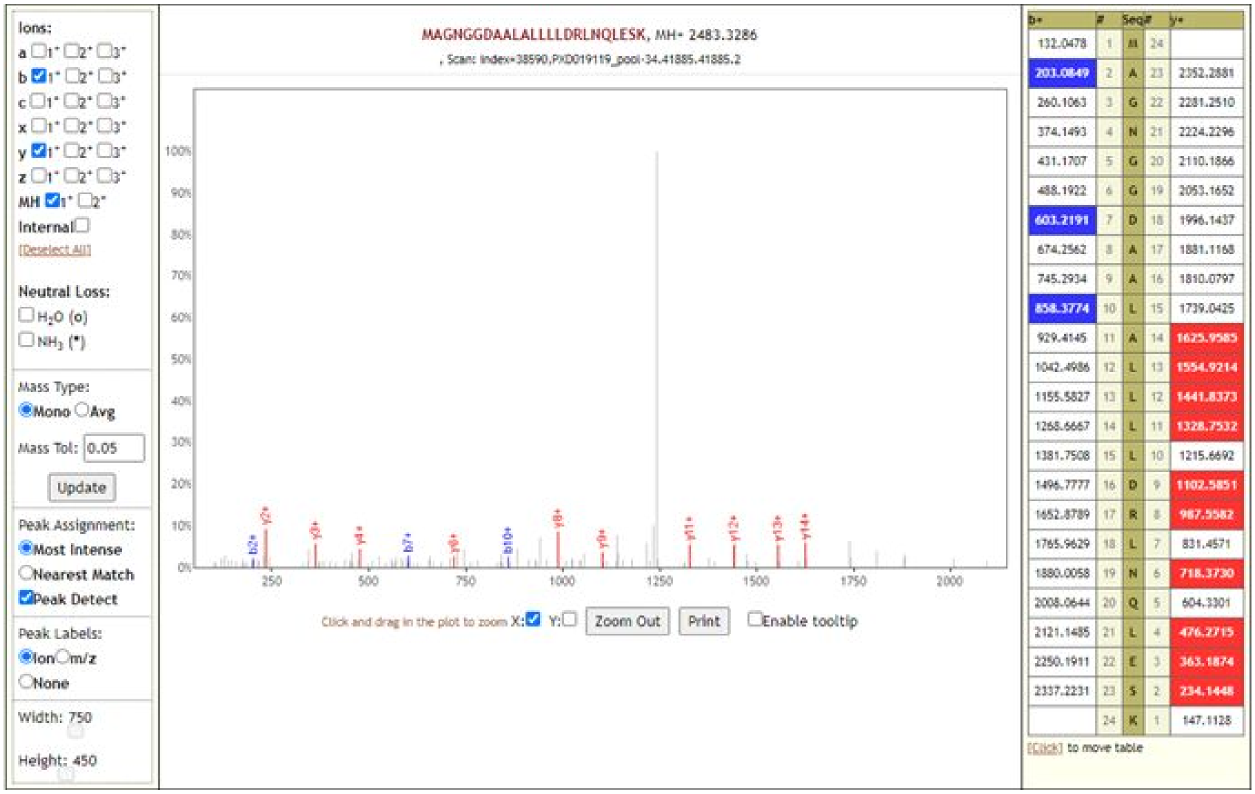

**TLLPAADLDDFSK**

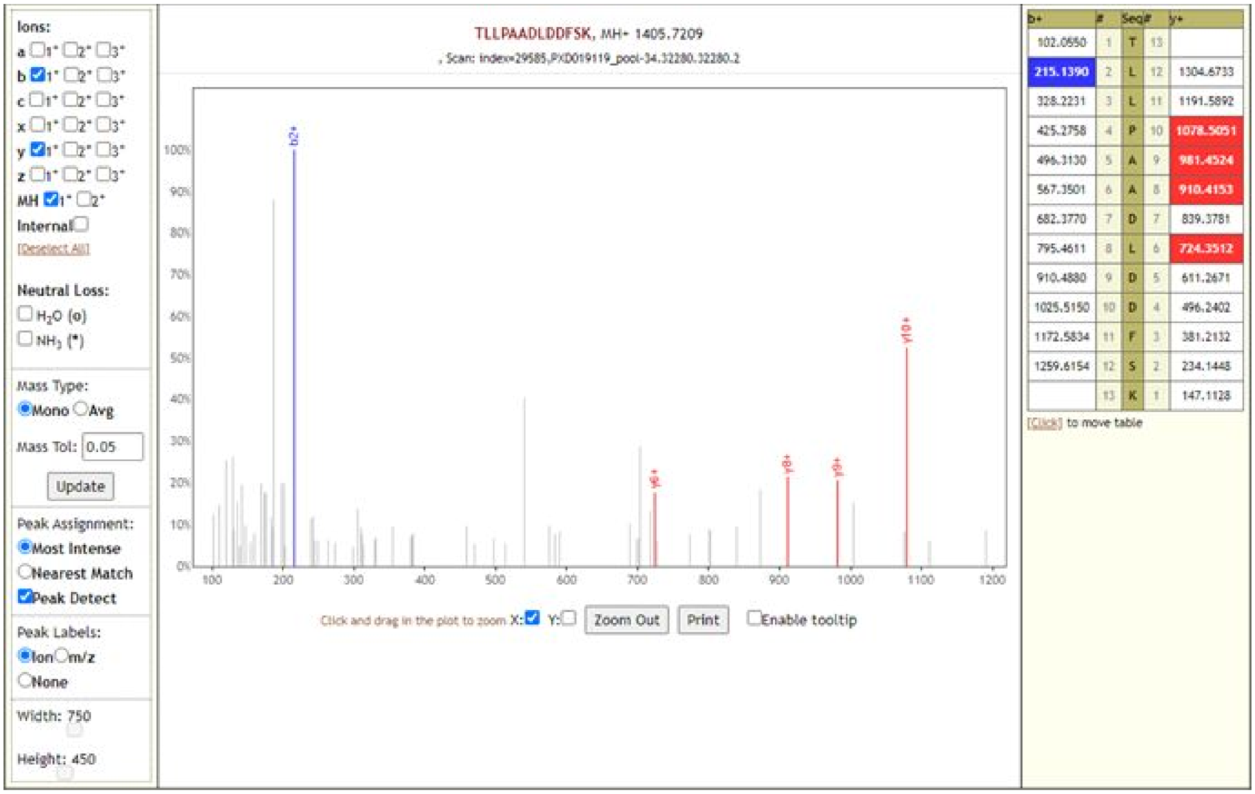

### QKRTATKAYNVTQAFGR

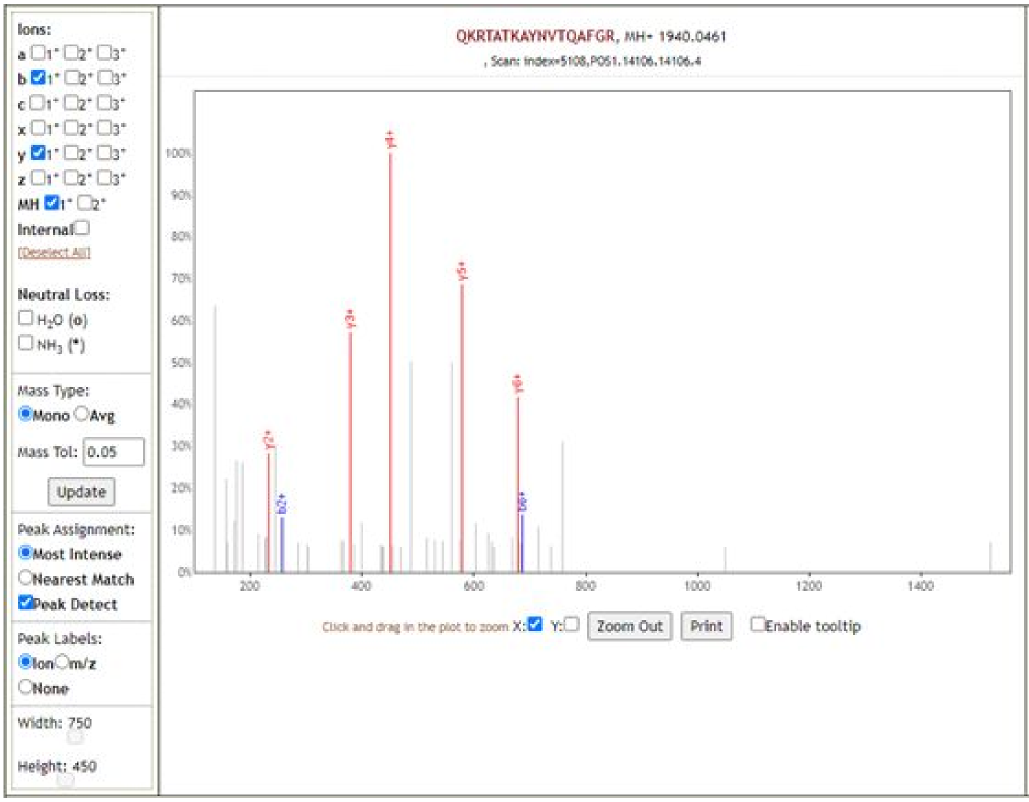

| b+ | # | Seq | # | y+ |
|---|---|---|---|---|
| 129.0659 | 1 | Q | 17 | |
| 257.1608 | 2 | K | 16 | 1811.9875 |
| 413.2619 | 3 | R | 15 | 1683.8925 |
| 514.3096 | 4 | T | 14 | 1527.7914 |
| 585.3467 | 5 | A | 13 | 1426.7437 |
| 686.3944 | 6 | T | 12 | 1355.7066 |
| 814.4894 | 7 | K | 11 | 1254.6589 |
| 885.5265 | 8 | A | 10 | 1126.5640 |
| 1048.5898 | 9 | Y | 9 | 1055.5269 |
| 1162.6327 | 10 | N | 8 | 892.4635 |
| 1261.7011 | 11 | V | 7 | 778.4206 |
| 1362.7488 | 12 | T | 6 | 679.3522 |
| 1490.8074 | 13 | Q | 5 | 578.3045 |
| 1561.8445 | 14 | A | 4 | 450.2459 |
| 1708.9129 | 15 | F | 3 | 379.2088 |
| 1765.9344 | 16 | G | 2 | 232.1404 |
| | 17 | R | 1 | 175.1190 |

[Click] to move table

### RTATKAYNVTQAFGR

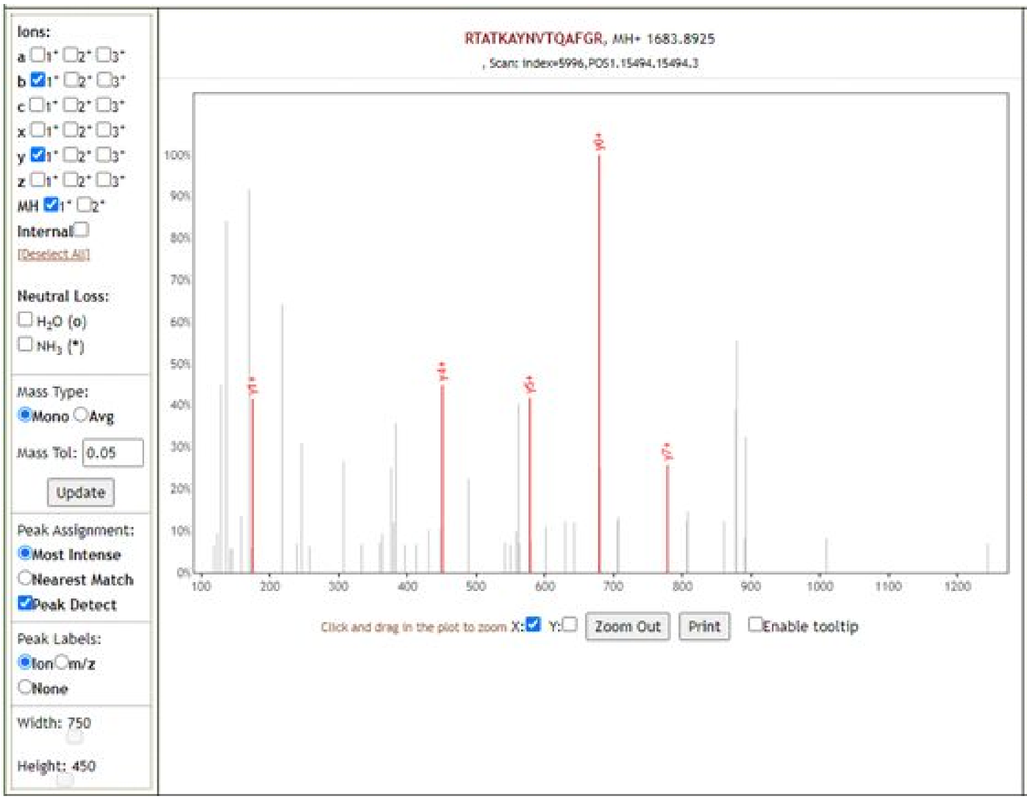

| b+ | # | Seq | # | y+ |
|---|---|---|---|---|
| 157.1084 | 1 | R | 15 | |
| 258.1561 | 2 | T | 14 | 1527.7914 |
| 329.1932 | 3 | A | 13 | 1426.7437 |
| 430.2409 | 4 | T | 12 | 1355.7066 |
| 558.3358 | 5 | K | 11 | 1254.6589 |
| 629.3729 | 6 | A | 10 | 1126.5640 |
| 792.4363 | 7 | Y | 9 | 1055.5269 |
| 906.4792 | 8 | N | 8 | 892.4635 |
| 1005.5476 | 9 | V | 7 | 778.4206 |
| 1106.5953 | 10 | T | 6 | 679.3522 |
| 1234.6539 | 11 | Q | 5 | 578.3045 |
| 1305.6910 | 12 | A | 4 | 450.2459 |
| 1452.7594 | 13 | F | 3 | 379.2088 |
| 1509.7809 | 14 | G | 2 | 232.1404 |
| | 15 | R | 1 | 175.1190 |

[Click] to move table

### TATKAYNVTQAFGR

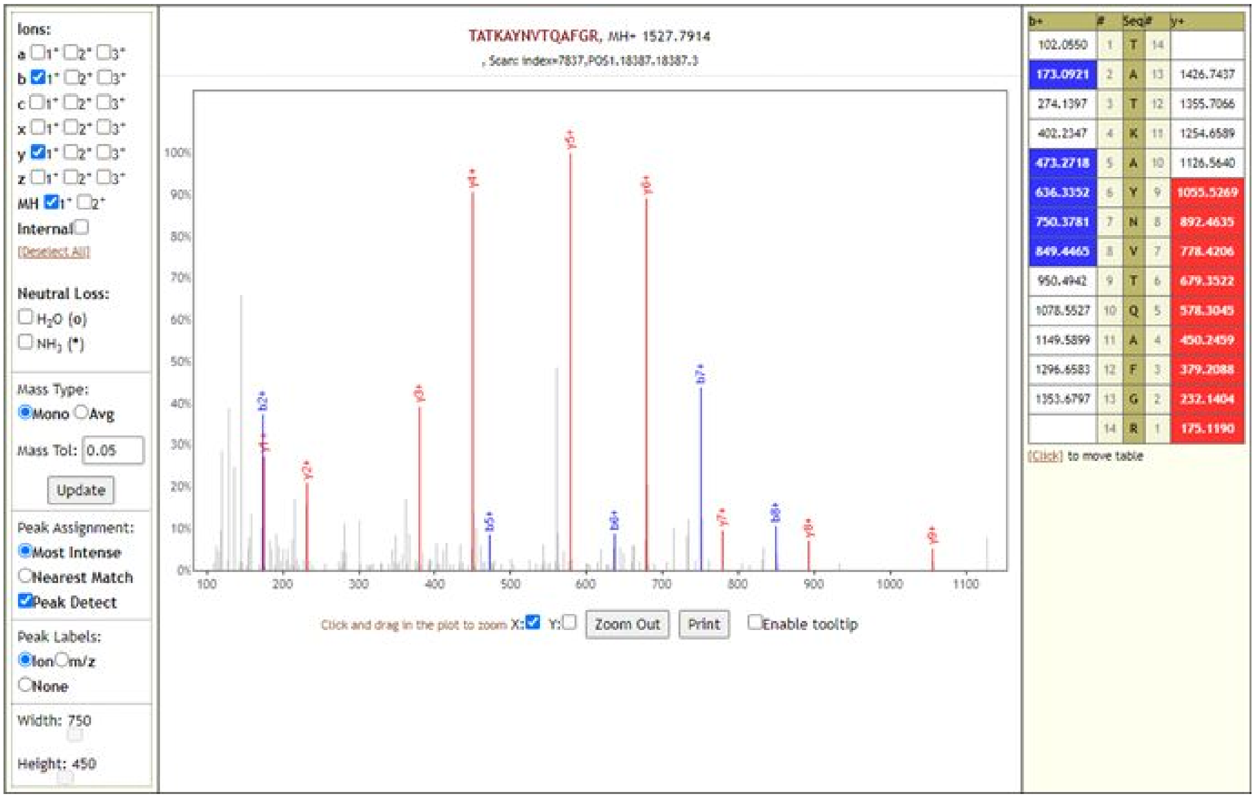

| b+ | # | Seq | # | y+ |
|---|---|---|---|---|
| 102.0550 | 1 | T | 14 | |
| 173.0921 | 2 | A | 13 | 1426.7437 |
| 274.1397 | 3 | T | 12 | 1355.7066 |
| 402.2347 | 4 | K | 11 | 1254.6589 |
| 473.2718 | 5 | A | 10 | 1126.5640 |
| 636.3352 | 6 | Y | 9 | 1055.5269 |
| 750.3781 | 7 | N | 8 | 892.4635 |
| 849.4465 | 8 | V | 7 | 778.4206 |
| 950.4942 | 9 | T | 6 | 679.3522 |
| 1078.5527 | 10 | Q | 5 | 578.3045 |
| 1149.5899 | 11 | A | 4 | 450.2459 |
| 1296.6583 | 12 | F | 3 | 379.2088 |
| 1353.6797 | 13 | G | 2 | 232.1404 |
| | 14 | R | 1 | 175.1190 |

### VDGVVQQLPETYFTQSR

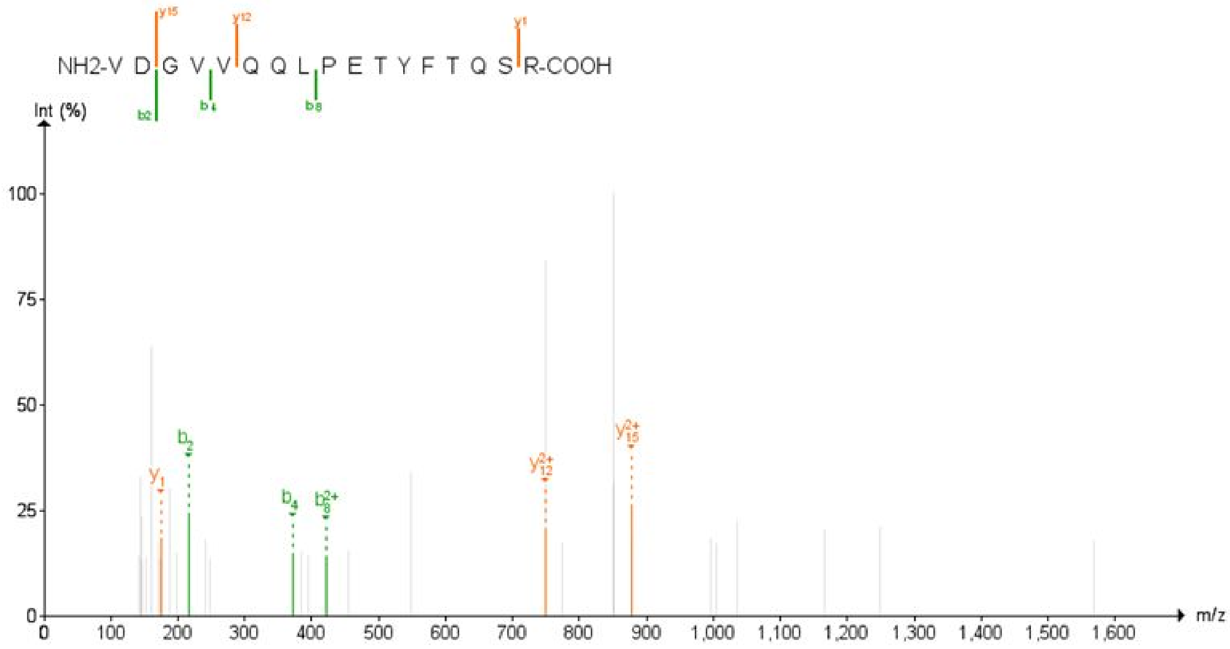

**VYSSANNCTFEYVSQPFLMDLEGK**

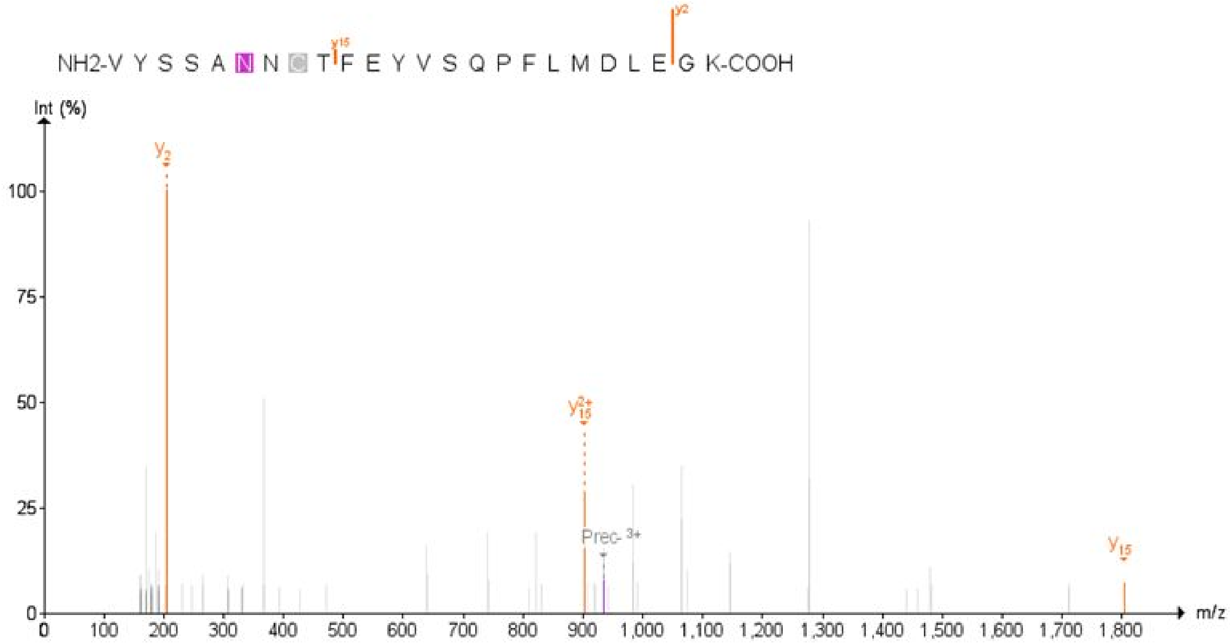

**NIDGYFK**

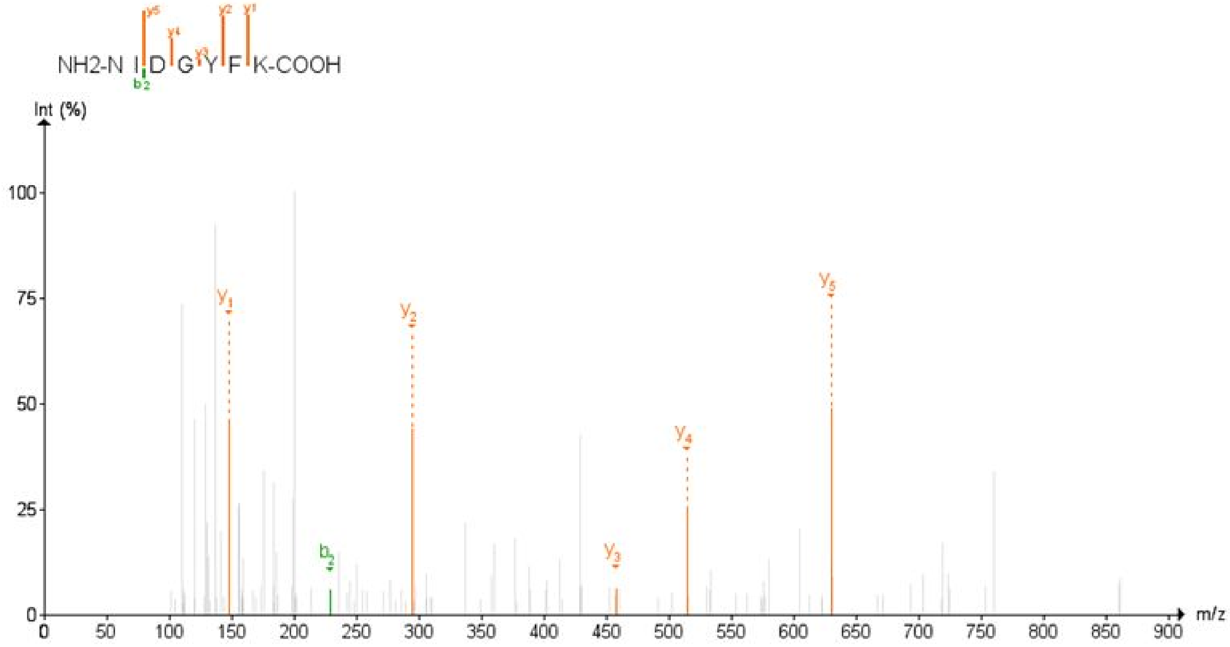

### VQPTESIVR

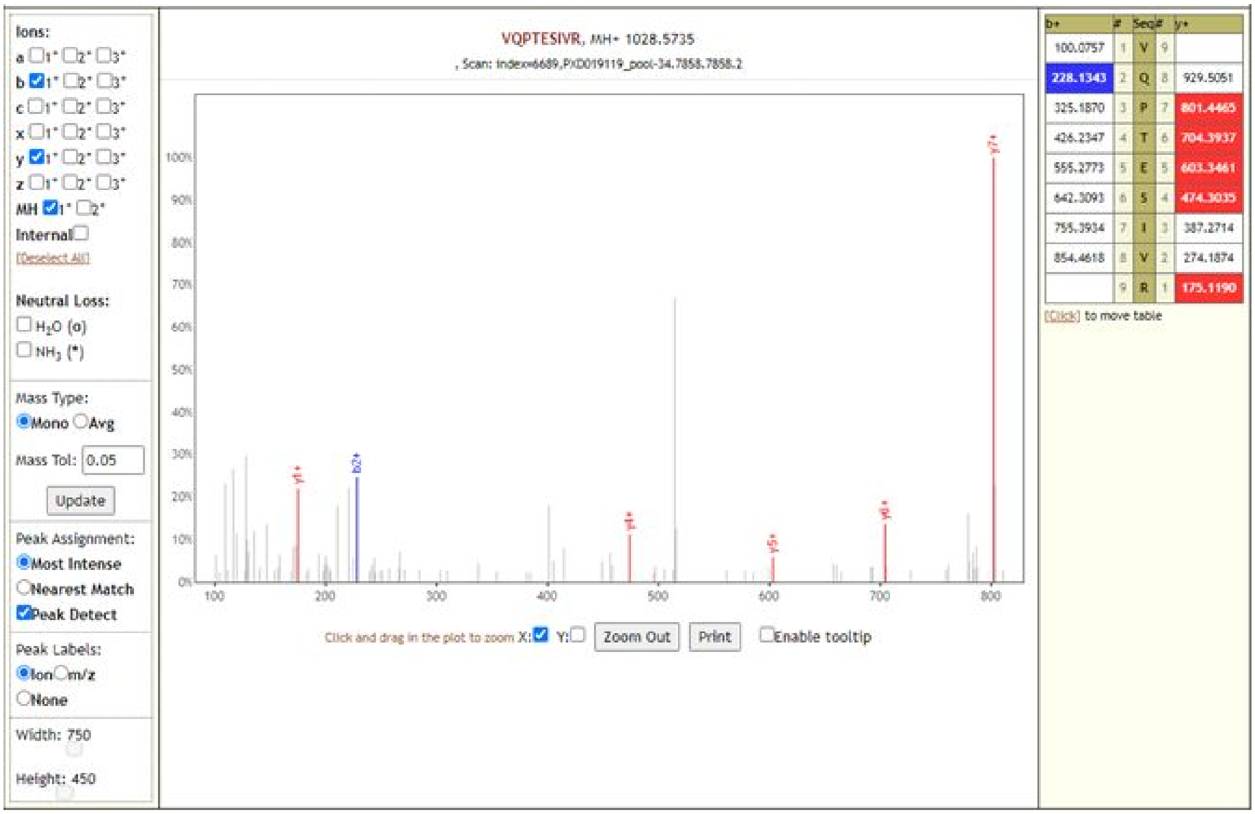

### ADETQALPQR

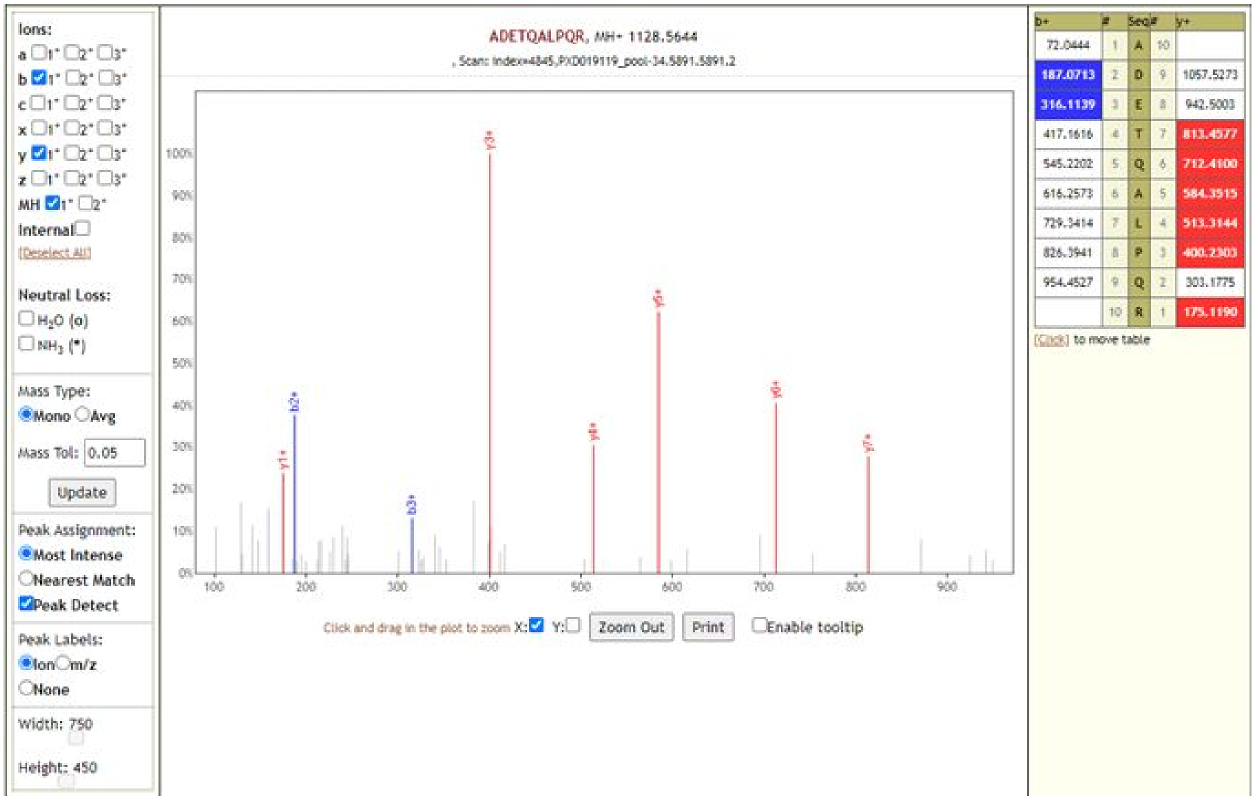

### FDNPVLPFNDGVYFASTEK

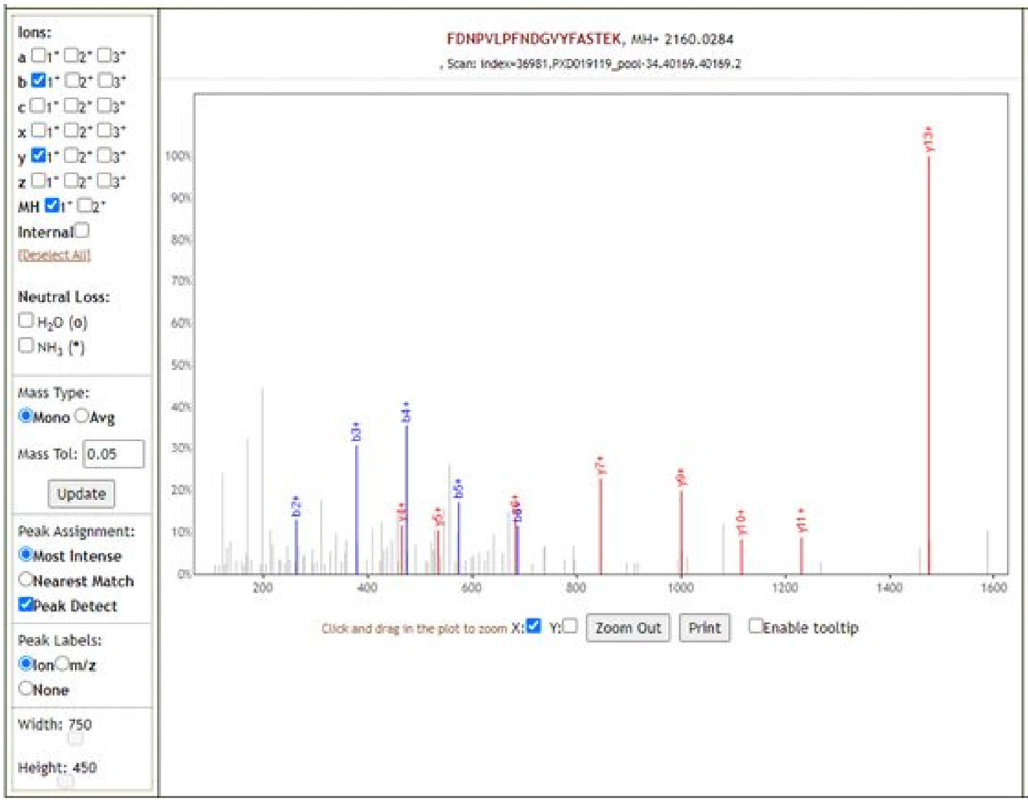

| b+ | # | Seq | # | y+ |
|---|---|---|---|---|
| 148.0757 | 1 | F | 19 | |
| 263.1026 | 2 | D | 18 | 2012.9600 |
| 377.1456 | 3 | N | 17 | 1897.9331 |
| 474.1983 | 4 | P | 16 | 1783.8901 |
| 573.2667 | 5 | V | 15 | 1686.8374 |
| 686.3508 | 6 | L | 14 | 1587.7690 |
| 783.4036 | 7 | P | 13 | 1474.6849 |
| 930.4720 | 8 | F | 12 | 1377.6321 |
| 1044.5149 | 9 | N | 11 | 1230.5637 |
| 1159.5419 | 10 | D | 10 | 1116.5208 |
| 1216.5633 | 11 | G | 9 | 1001.4938 |
| 1315.6317 | 12 | V | 8 | 944.4724 |
| 1478.6951 | 13 | Y | 7 | 845.4040 |
| 1625.7635 | 14 | F | 6 | 682.3406 |
| 1696.8006 | 15 | A | 5 | 535.2722 |
| 1783.8326 | 16 | S | 4 | 464.2351 |
| 1884.8803 | 17 | T | 3 | 377.2031 |
| 2013.9229 | 18 | E | 2 | 276.1554 |
| | 19 | K | 1 | 147.1128 |

[Click] to move table

### GWIFGTTLDSK

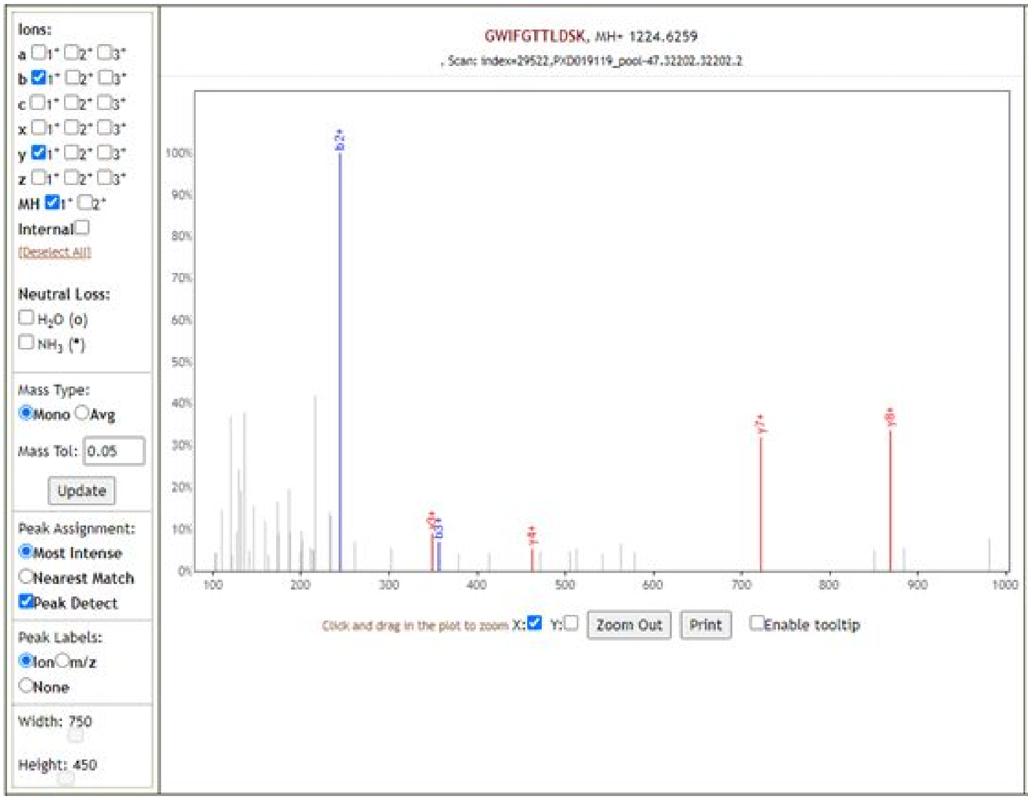

| b+ | # | Seq | # | y+ |
|---|---|---|---|---|
| 58.0287 | 1 | G | 11 | |
| 244.1081 | 2 | W | 10 | 1167.6045 |
| 357.1921 | 3 | I | 9 | 981.5251 |
| 504.2605 | 4 | F | 8 | 868.4411 |
| 561.2820 | 5 | G | 7 | 721.3727 |
| 662.3297 | 6 | T | 6 | 664.3512 |
| 763.3774 | 7 | T | 5 | 563.3035 |
| 876.4614 | 8 | L | 4 | 462.2558 |
| 991.4884 | 9 | D | 3 | 349.1718 |
| 1078.5204 | 10 | S | 2 | 234.1448 |
| | 11 | K | 1 | 147.1128 |

[Click] to move table

**QLQQSMSSADSTQA**

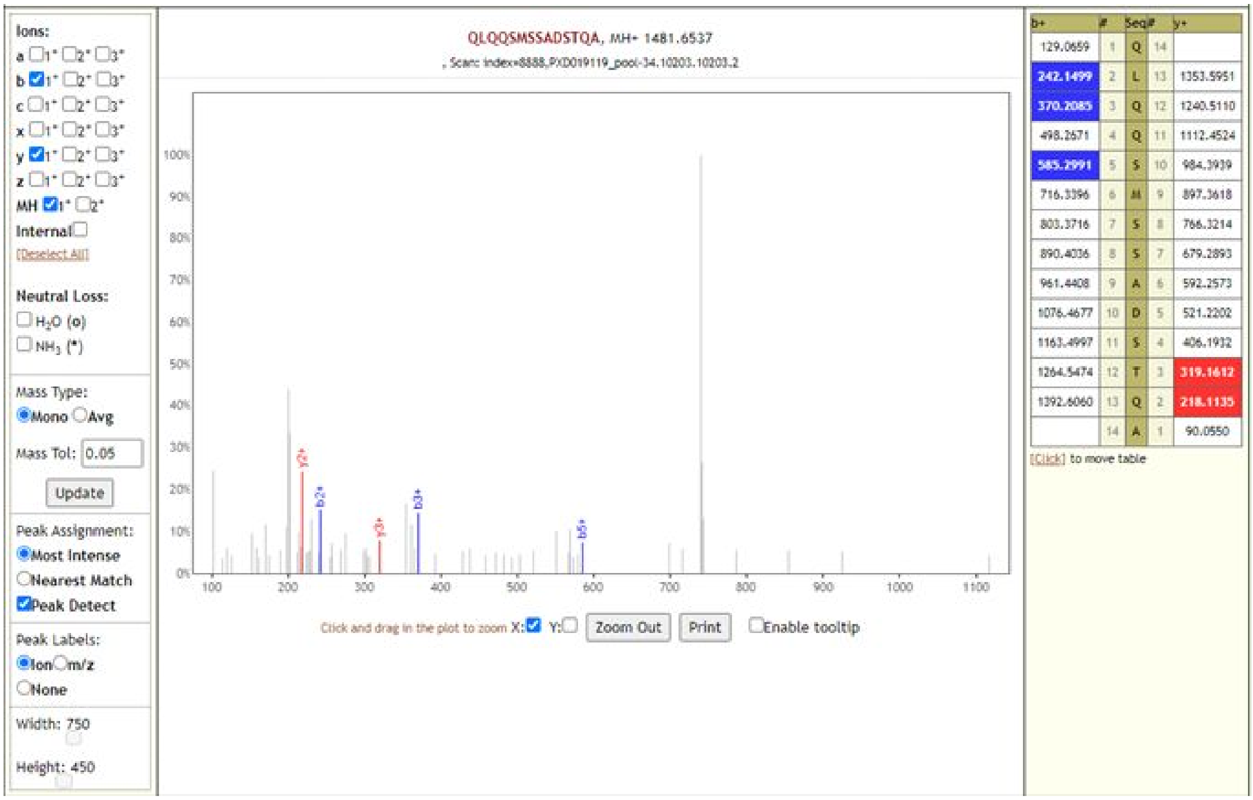

| b+ | # | Seq | # | y+ |
|---|---|---|---|---|
| 129.0659 | 1 | Q | 14 | |
| 242.1499 | 2 | L | 13 | 1353.5951 |
| 370.2085 | 3 | Q | 12 | 1240.5110 |
| 498.2671 | 4 | Q | 11 | 1112.4524 |
| 585.2991 | 5 | S | 10 | 984.3939 |
| 716.3396 | 6 | M | 9 | 897.3618 |
| 803.3716 | 7 | S | 8 | 766.3214 |
| 890.4036 | 8 | S | 7 | 679.2893 |
| 961.4408 | 9 | A | 6 | 592.2573 |
| 1076.4677 | 10 | D | 5 | 521.2202 |
| 1163.4997 | 11 | S | 4 | 406.1932 |
| 1264.5474 | 12 | T | 3 | 319.1612 |
| 1392.6060 | 13 | Q | 2 | 218.1135 |
| | 14 | A | 1 | 90.0550 |

**SWMESEFR**

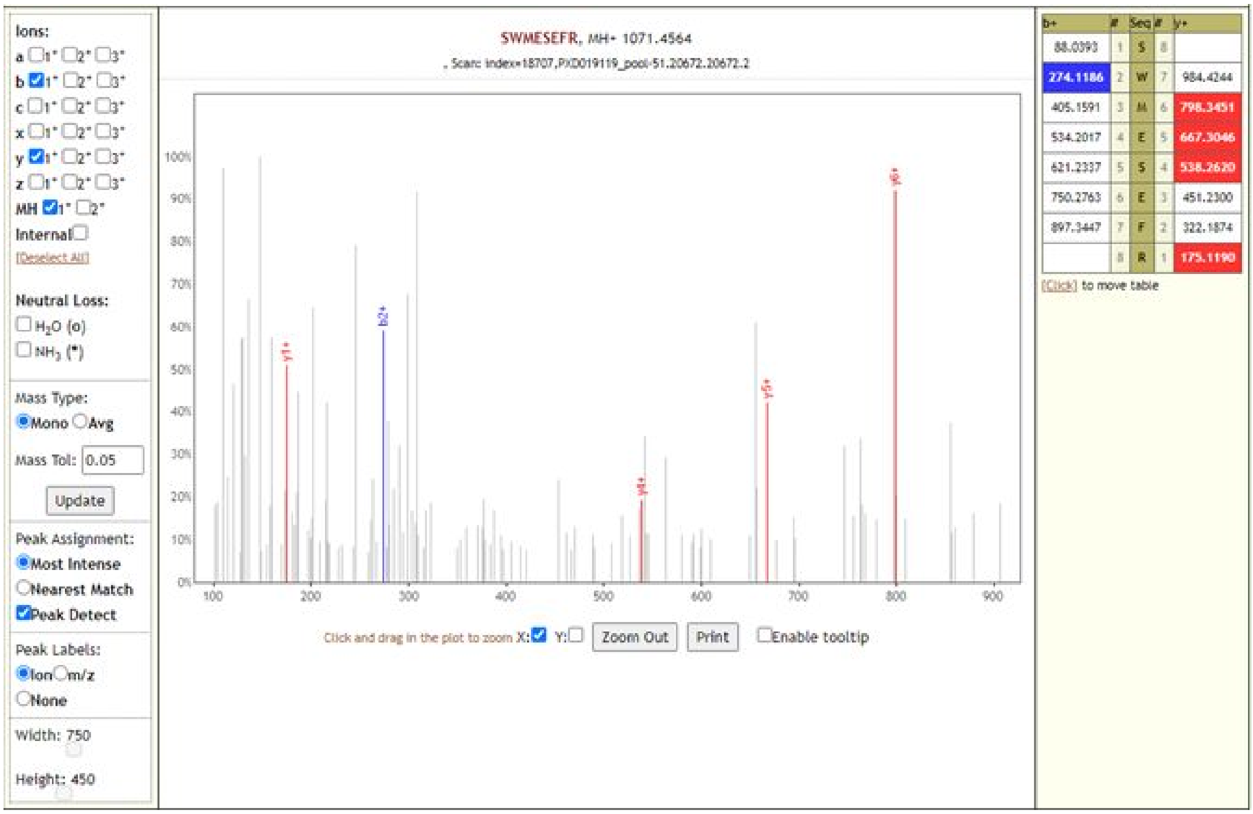

| b+ | # | Seq | # | y+ |
|---|---|---|---|---|
| 88.0393 | 1 | S | 8 | |
| 274.1186 | 2 | W | 7 | 984.4244 |
| 405.1591 | 3 | M | 6 | 798.3451 |
| 534.2017 | 4 | E | 5 | 667.3046 |
| 621.2337 | 5 | S | 4 | 538.2620 |
| 750.2763 | 6 | E | 3 | 451.2300 |
| 897.3447 | 7 | F | 2 | 322.1874 |
| | 8 | R | 1 | 175.1190 |

### TQLPPAYTNSFTR

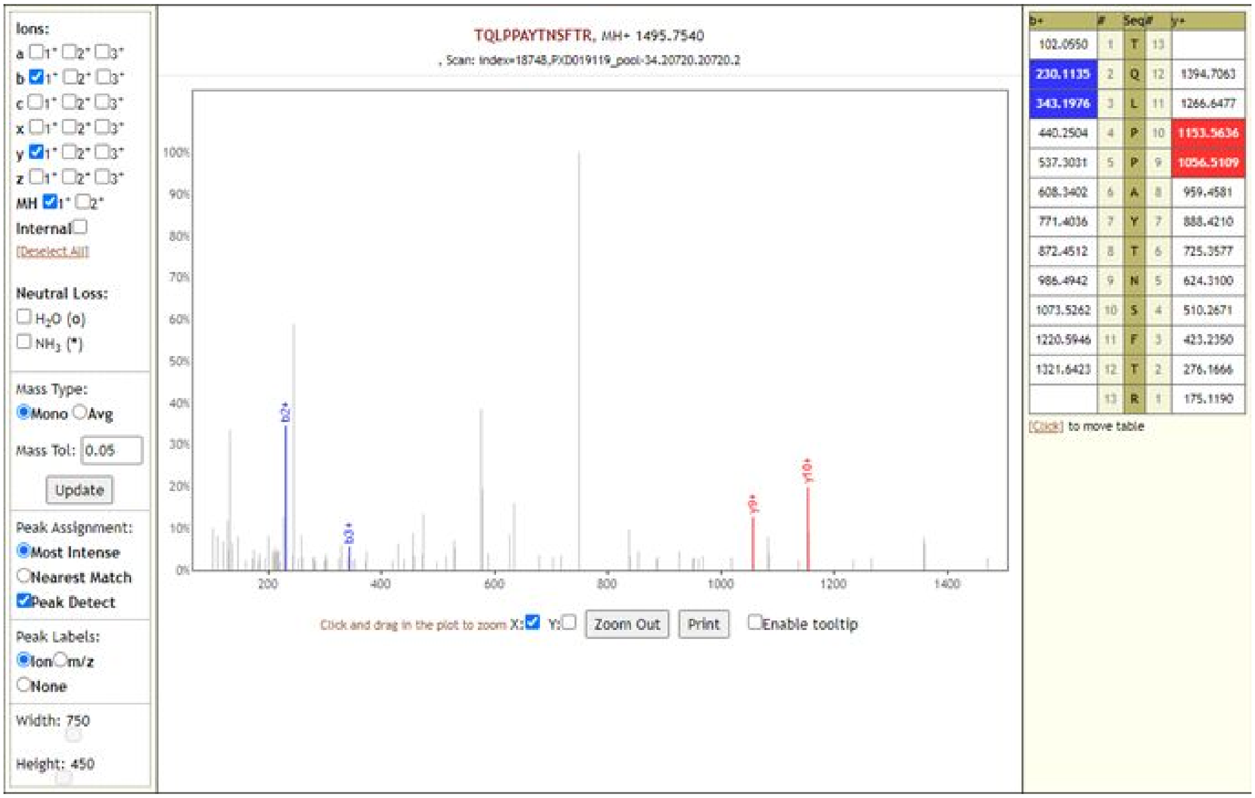

### VGGNYNYLYR

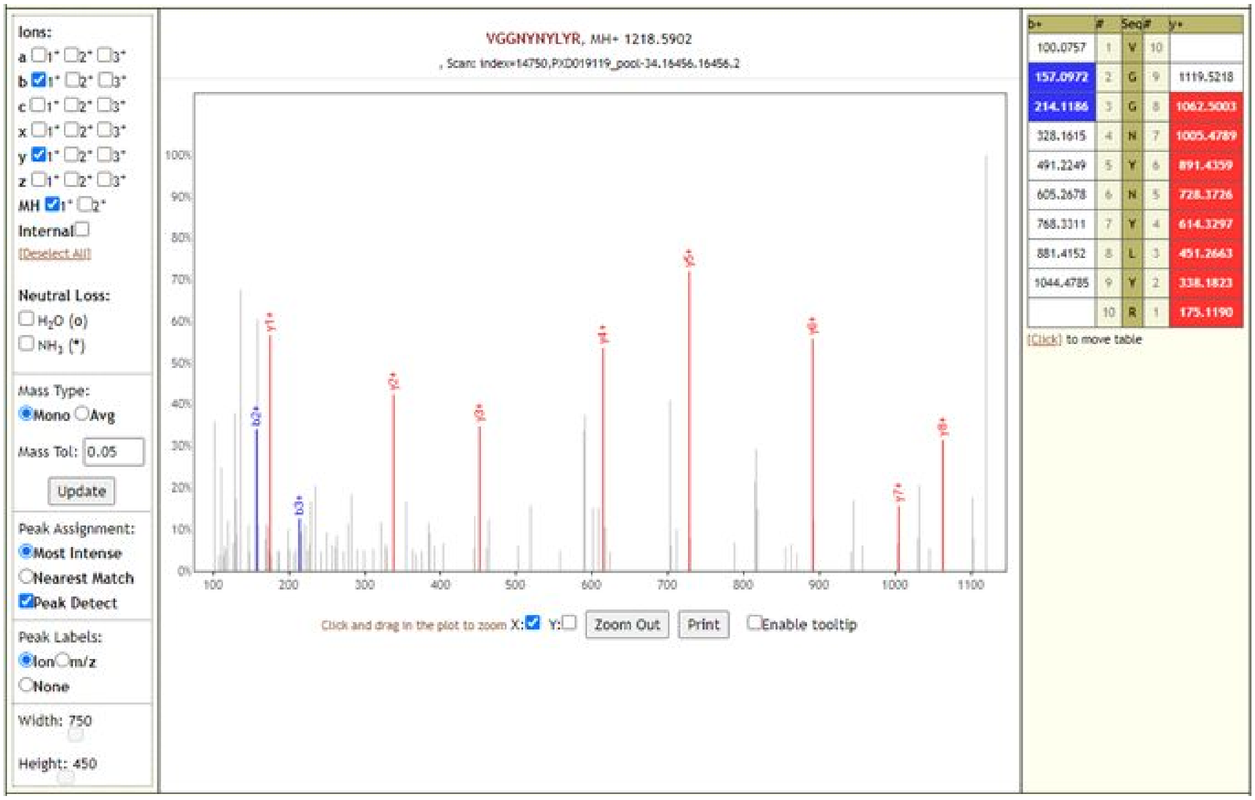

**VYSTGSNVFQTR**

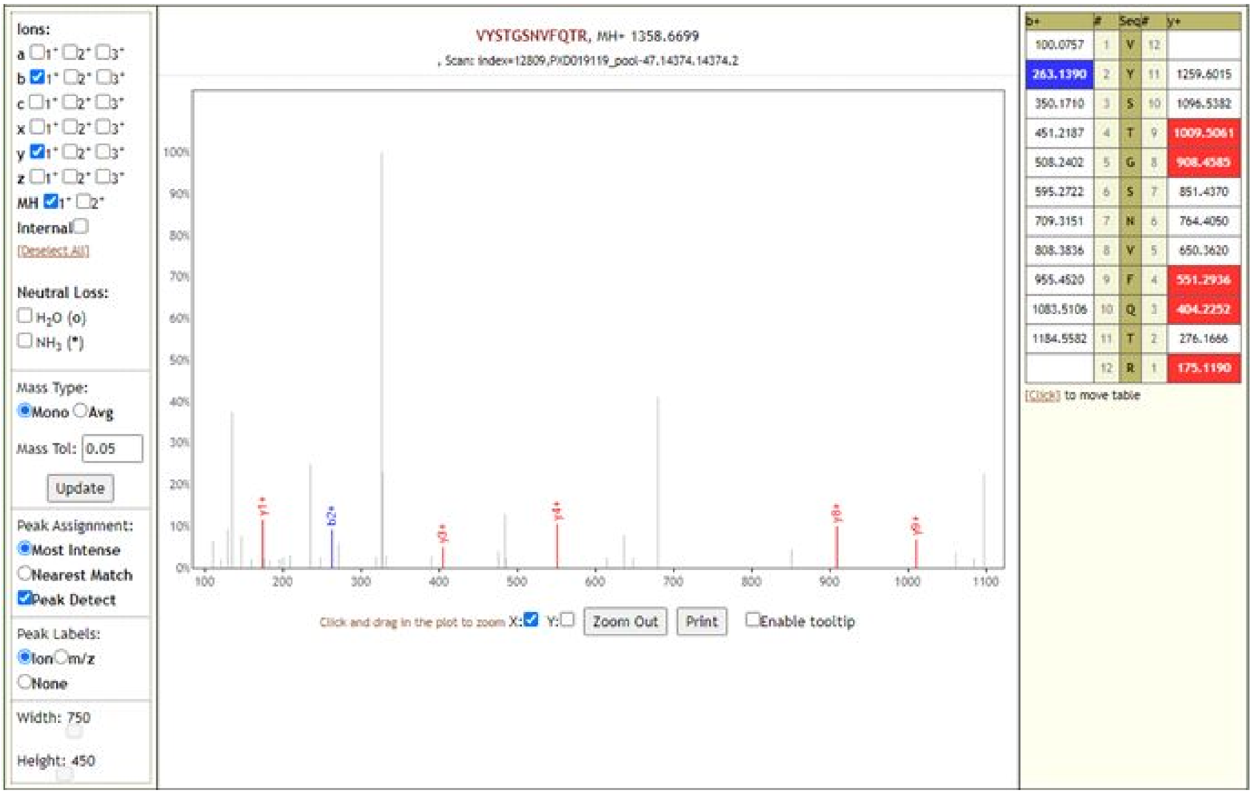

**DFGGFNFSQILPDPSKPSK**

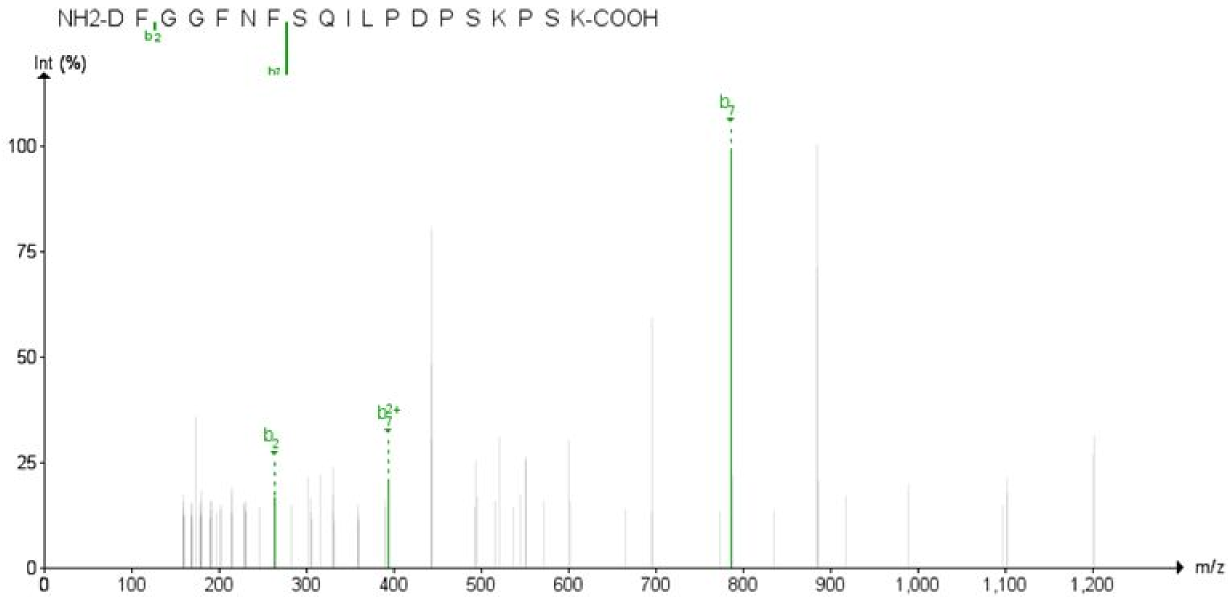

**DGIIWVATEGALNTPKDHIGTR**

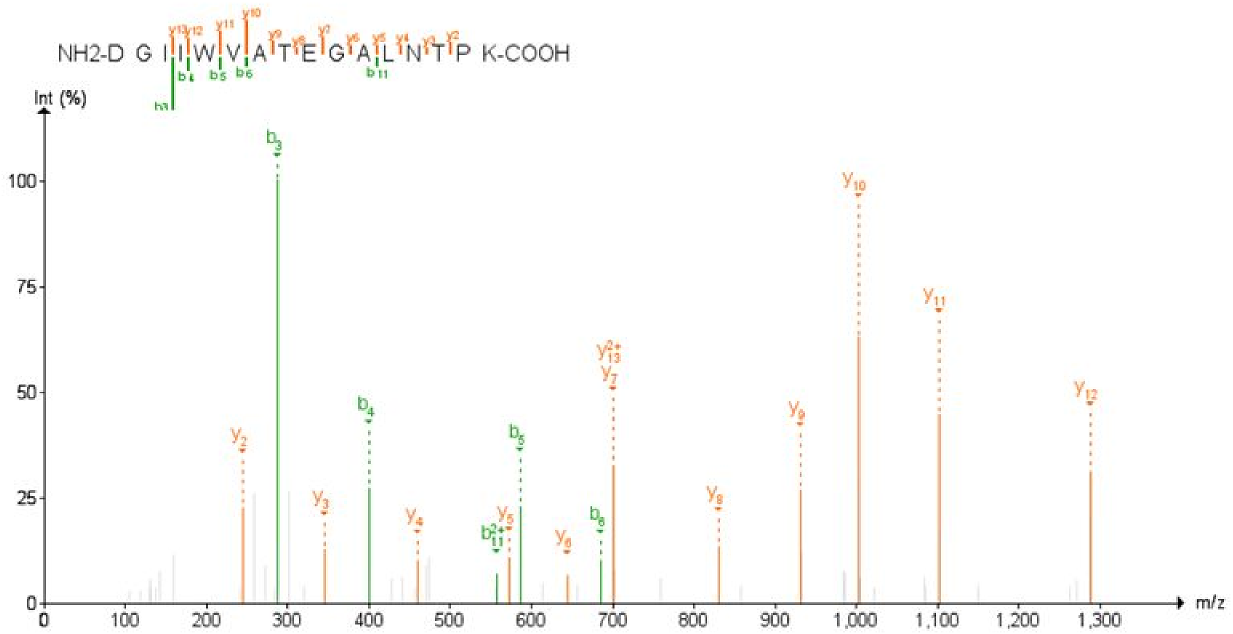

**DHIGTRNPANNAAIVLQLPQGTTLPK**

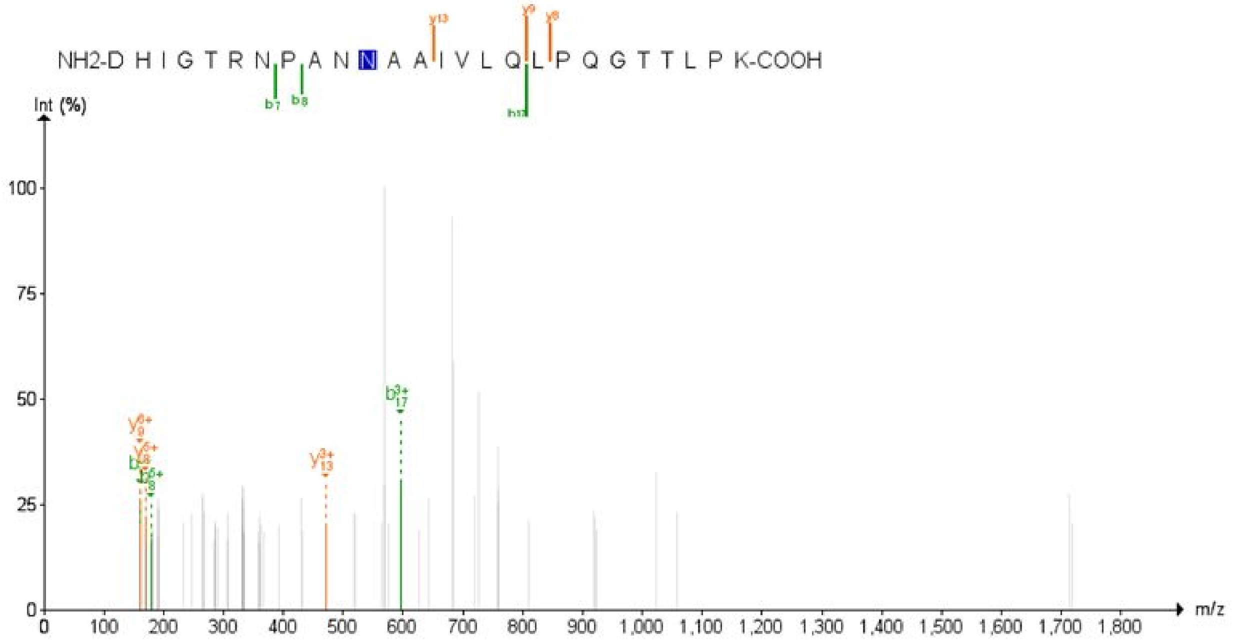

**FPNITNLCPFGEVFNATR**

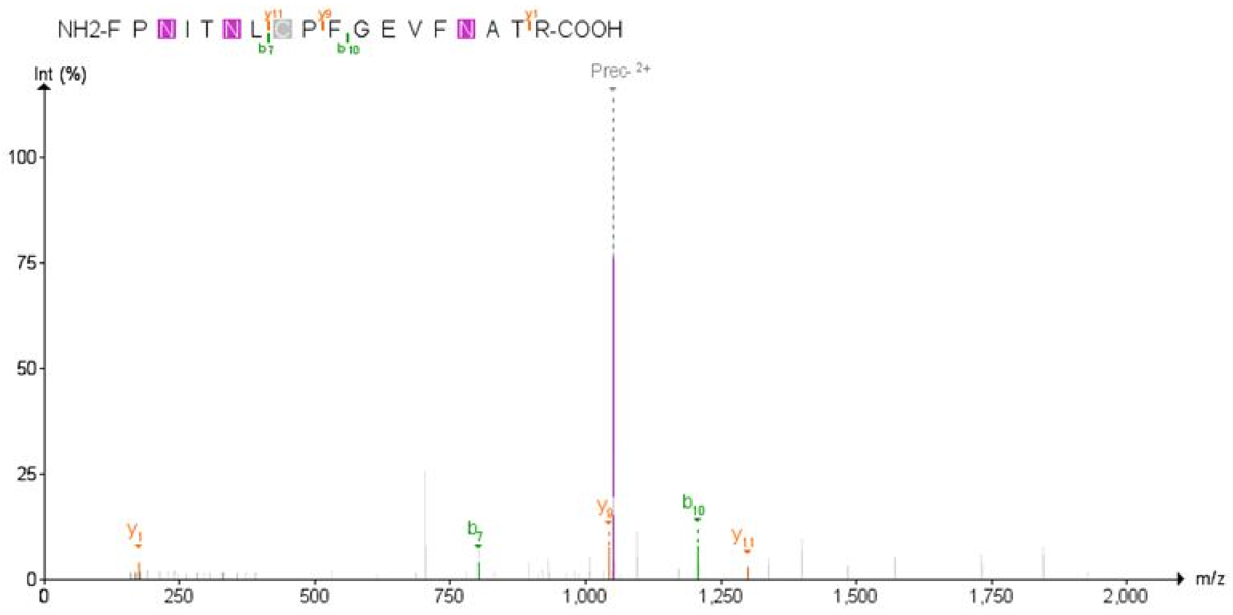

**IYCPACHNSEVGPEHSLAEYHNESGLK**

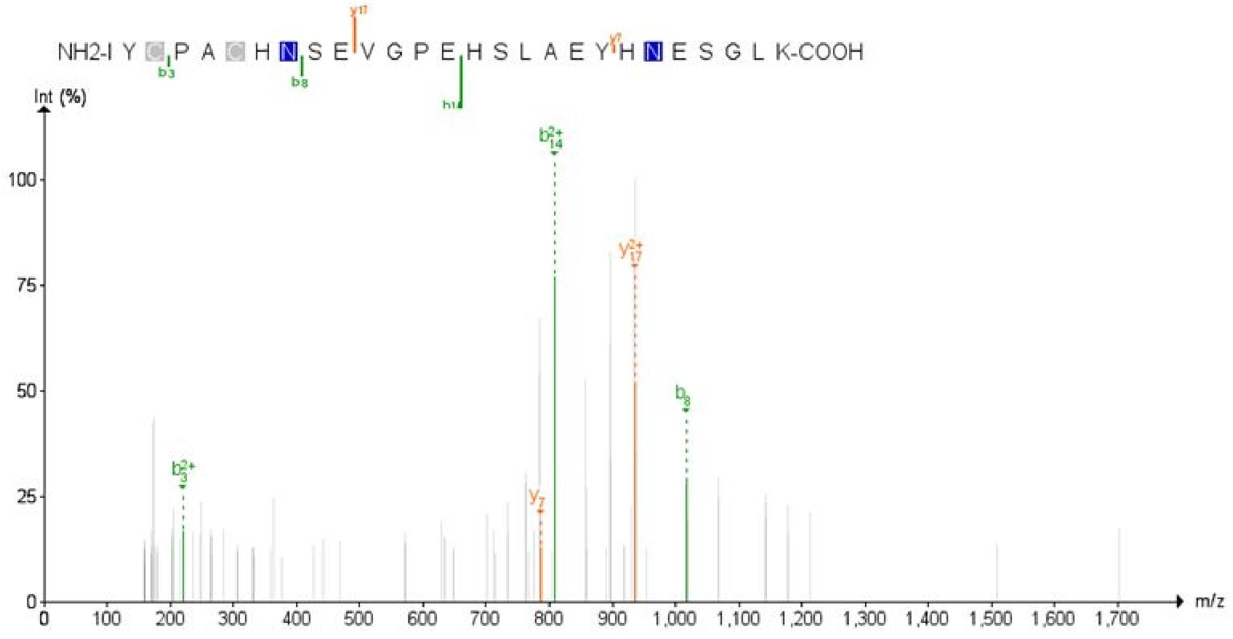

**NPLLYDANYFL**

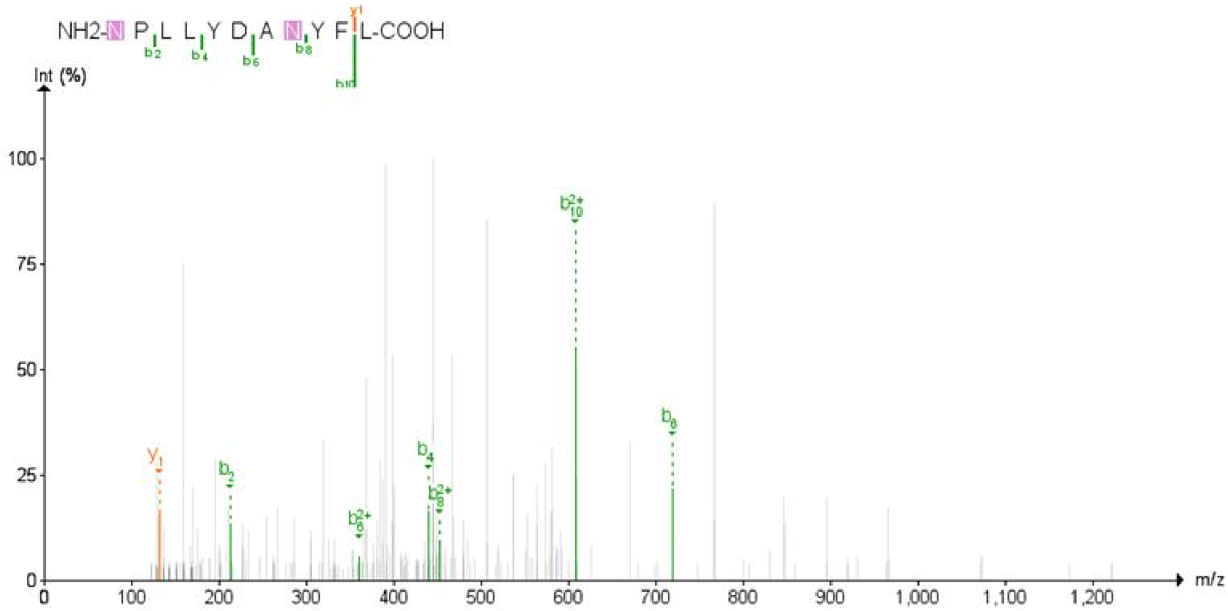

**RGQGVPINTNSSPDDQIGYYR**

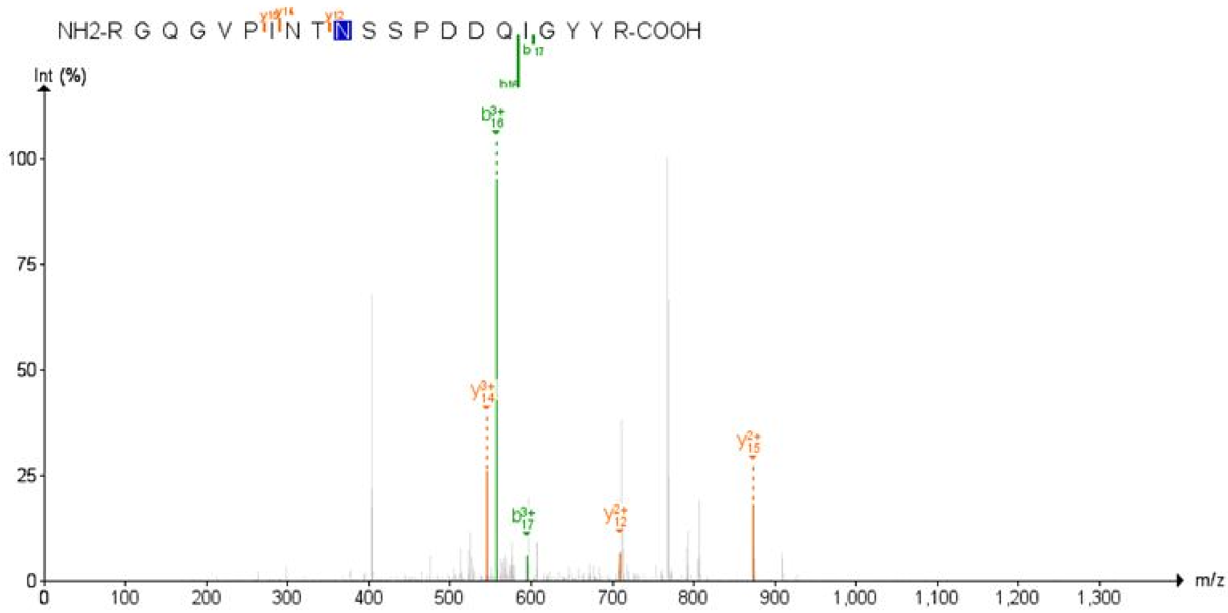

**STDTGVEHVTFFIYNK**

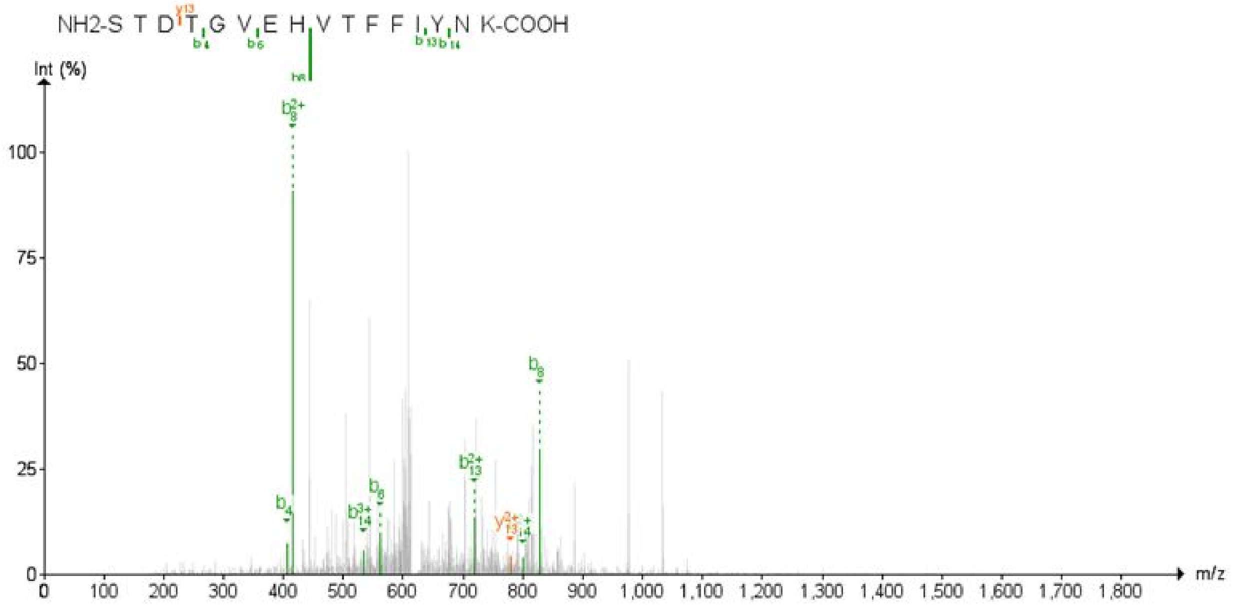

**VAGDSGFAAYSRYR**

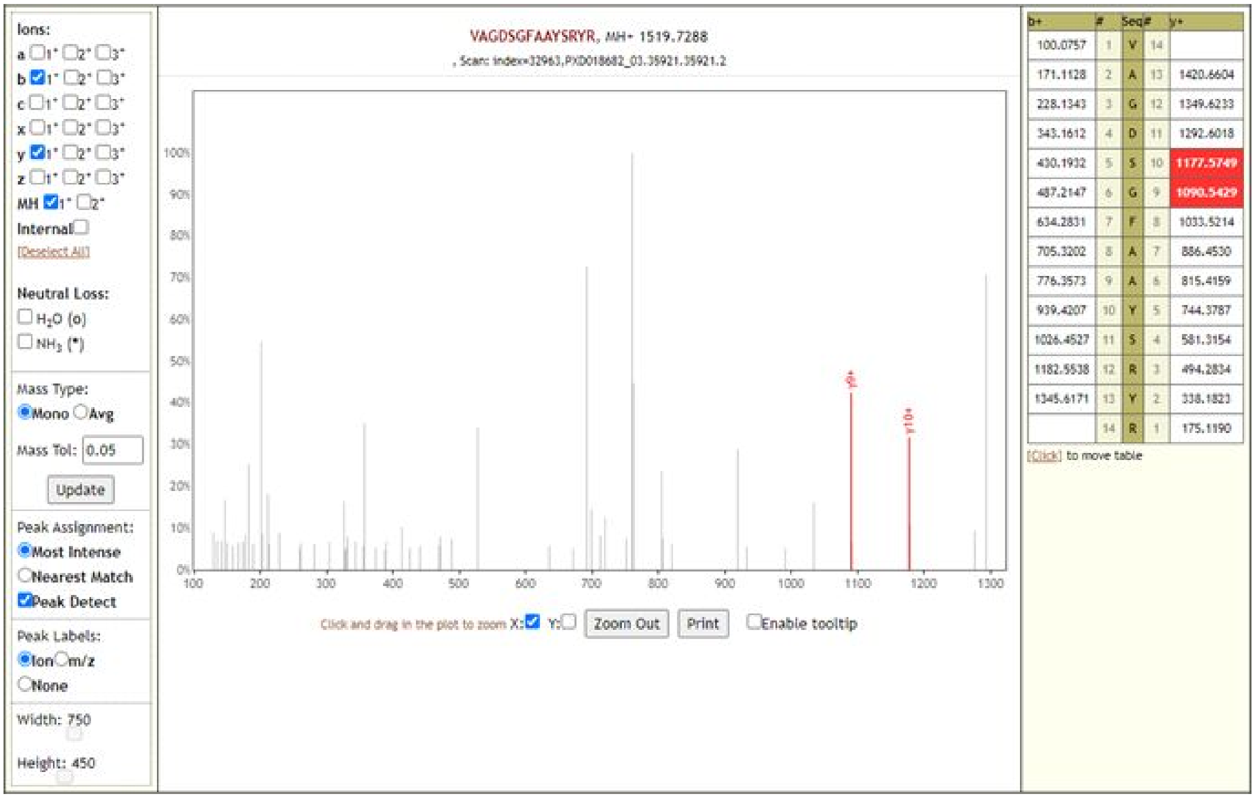

| b+ | # | Seq | # | y+ |
|---|---|---|---|---|
| 100.0757 | 1 | V | 14 | |
| 171.1128 | 2 | A | 13 | 1420.6604 |
| 228.1343 | 3 | G | 12 | 1349.6233 |
| 343.1612 | 4 | D | 11 | 1292.6018 |
| 430.1932 | 5 | S | 10 | 1177.5749 |
| 487.2147 | 6 | G | 9 | 1090.5429 |
| 634.2831 | 7 | F | 8 | 1033.5214 |
| 705.3202 | 8 | A | 7 | 886.4530 |
| 776.3573 | 9 | A | 6 | 815.4159 |
| 939.4207 | 10 | Y | 5 | 744.3787 |
| 1026.4527 | 11 | S | 4 | 581.3154 |
| 1182.5538 | 12 | R | 3 | 494.2834 |
| 1345.6171 | 13 | Y | 2 | 338.1823 |
| | 14 | R | 1 | 175.1190 |

#### ALNLGETFVTHSK

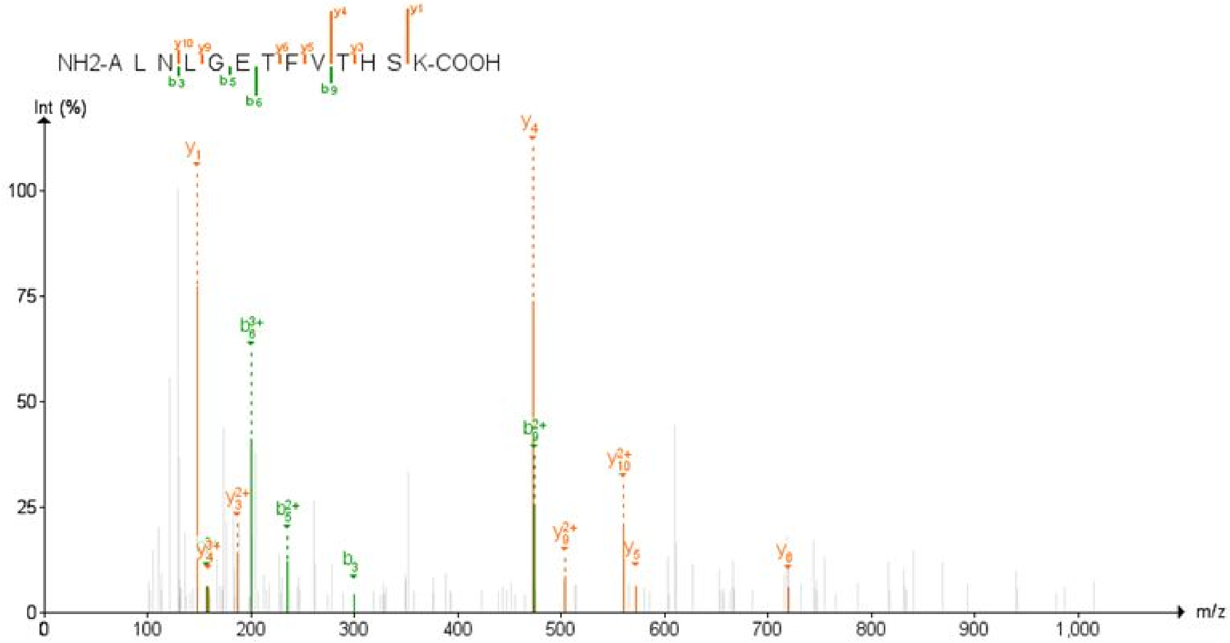

#### ALTGIAVEQDKNTQEVFAQVK

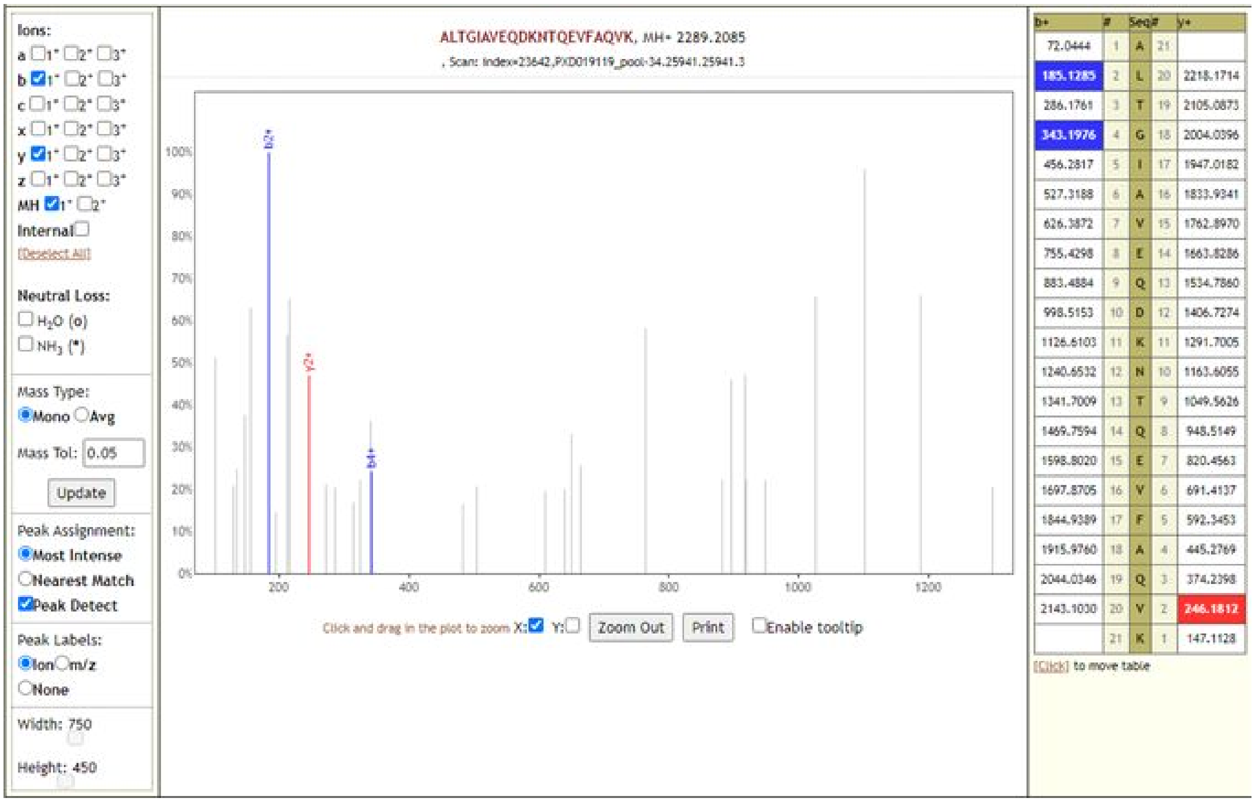

### ALTQHGKEDLK

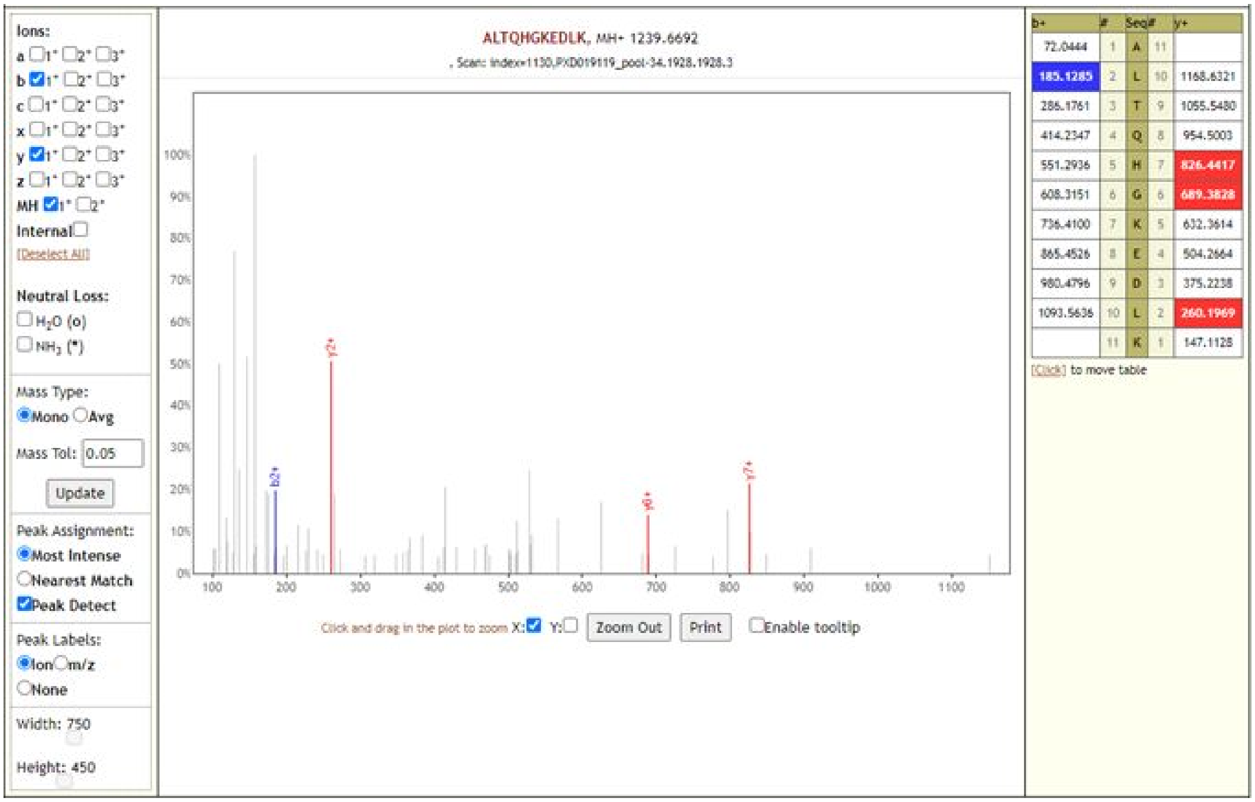

### DGTCGLVEVEK

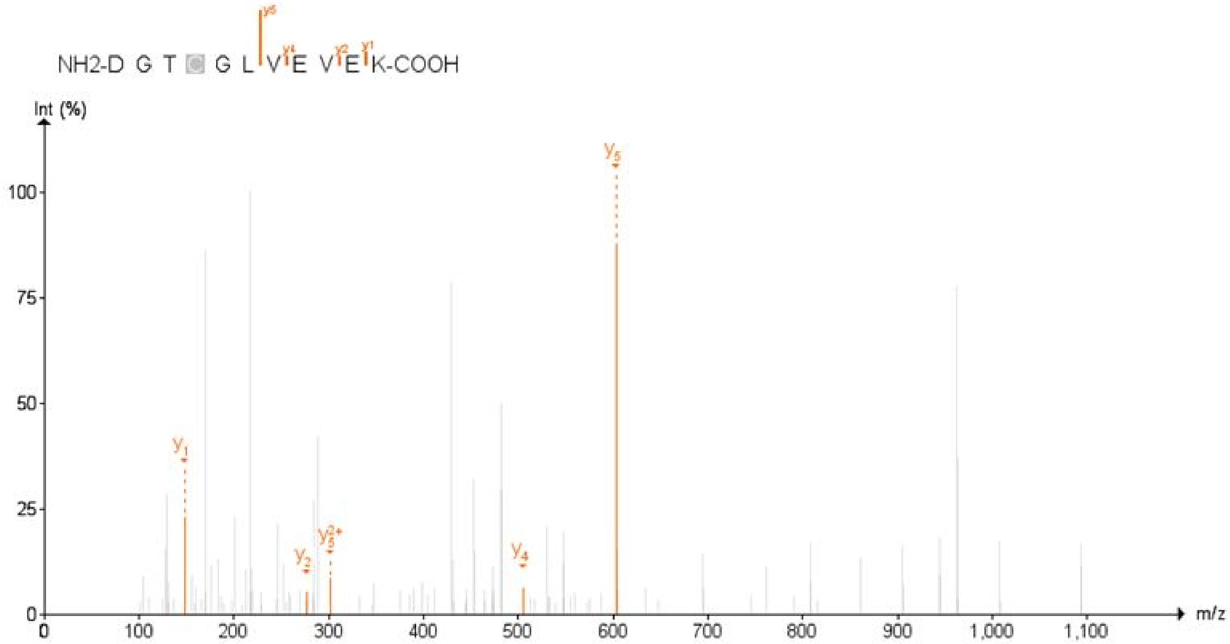

**FTALTQHGKEDLK**

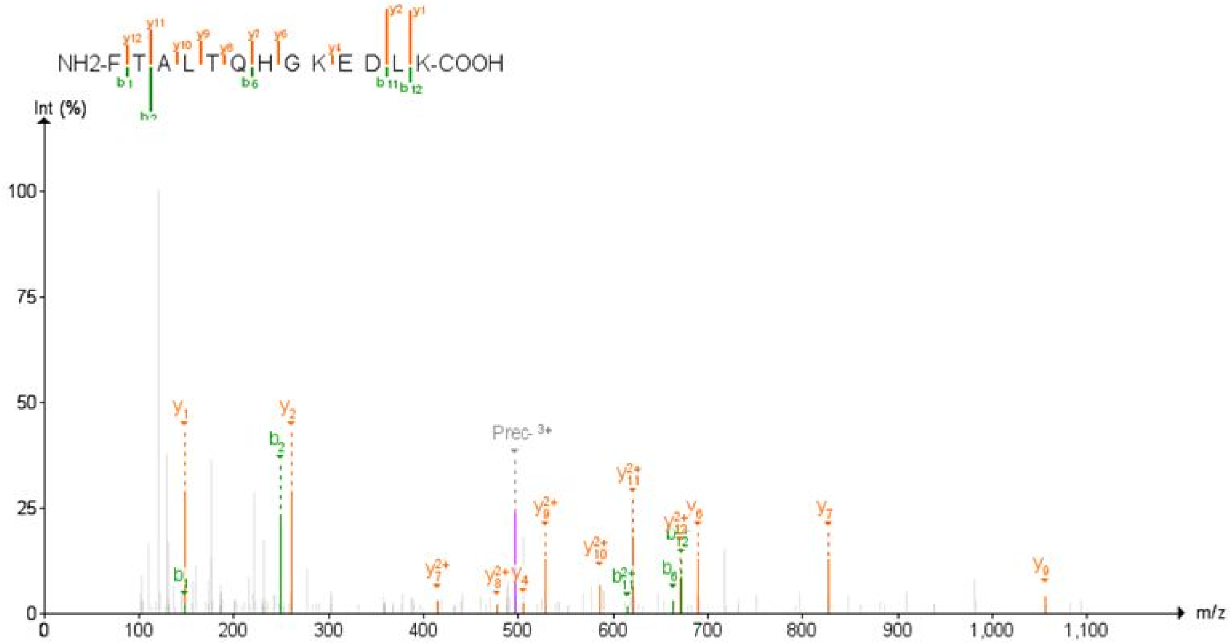

**GVQIPCTCGK**

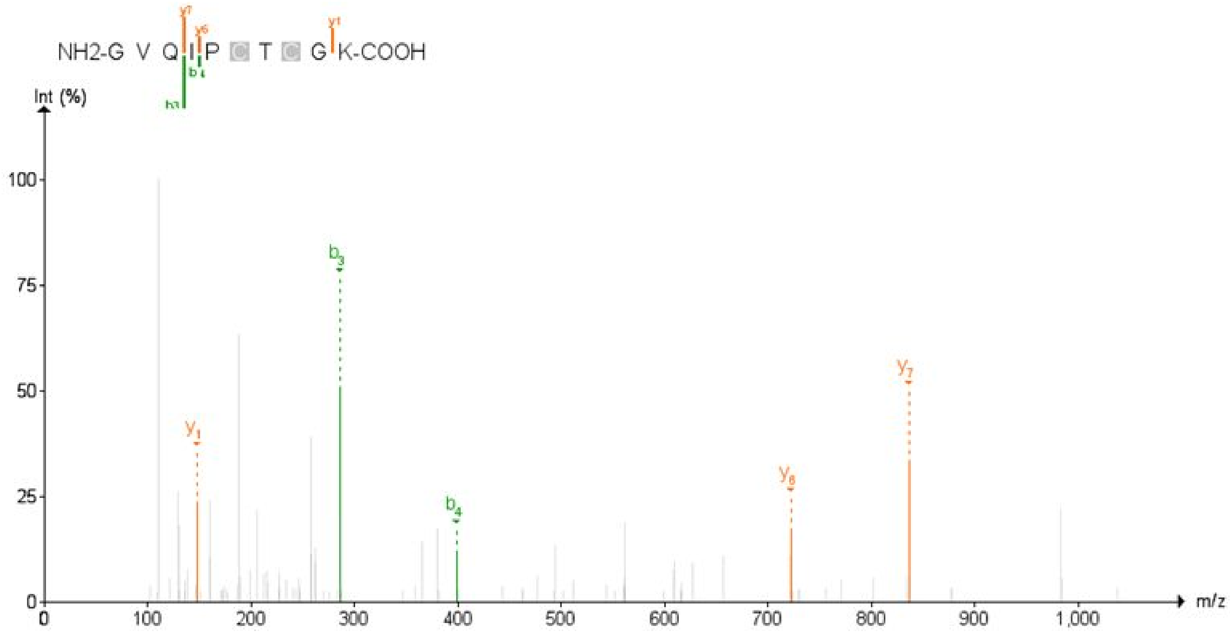

**GVYYPDKVFR**

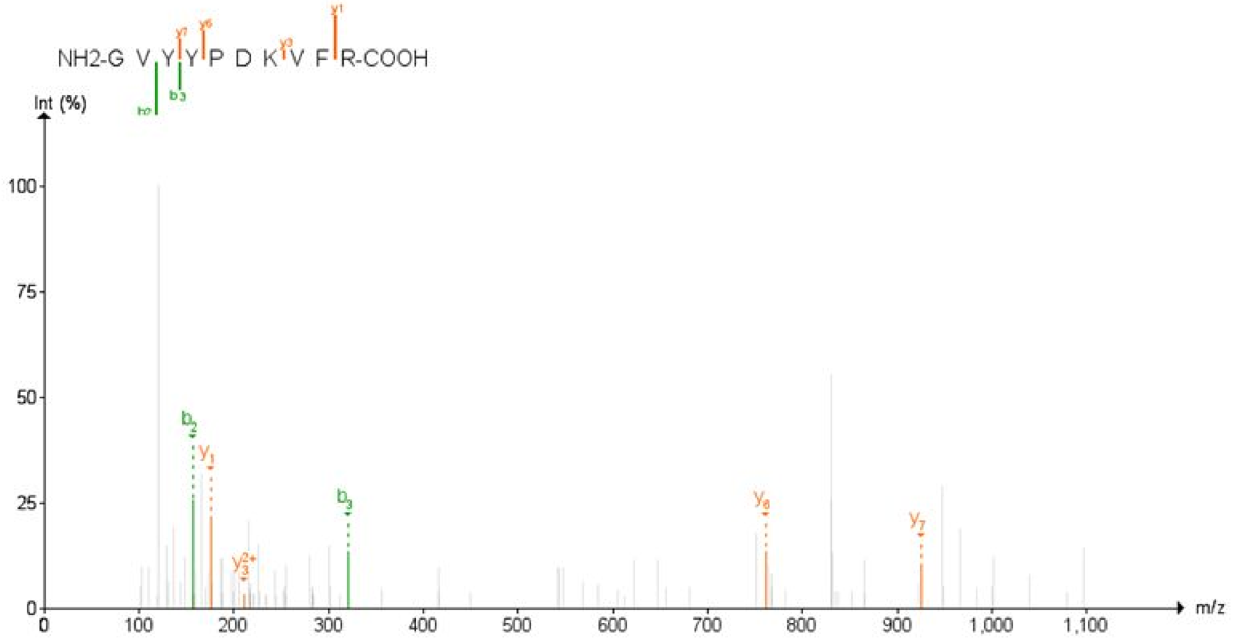

**IVDEPEEHVQIHTID**

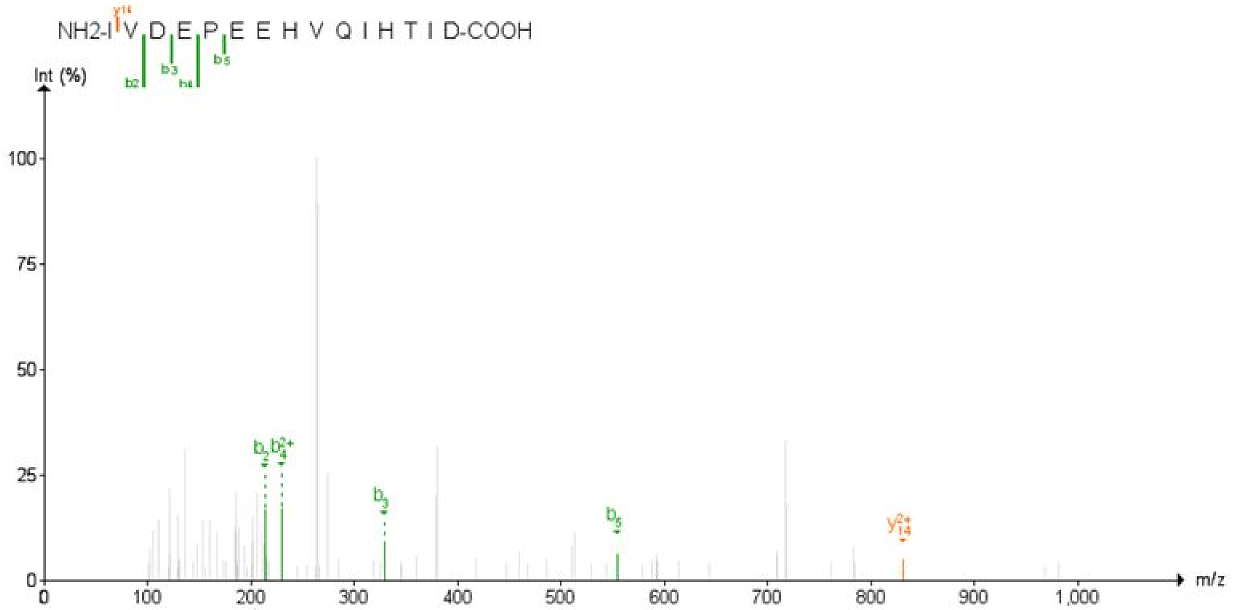

**SPDDQIGYYR**

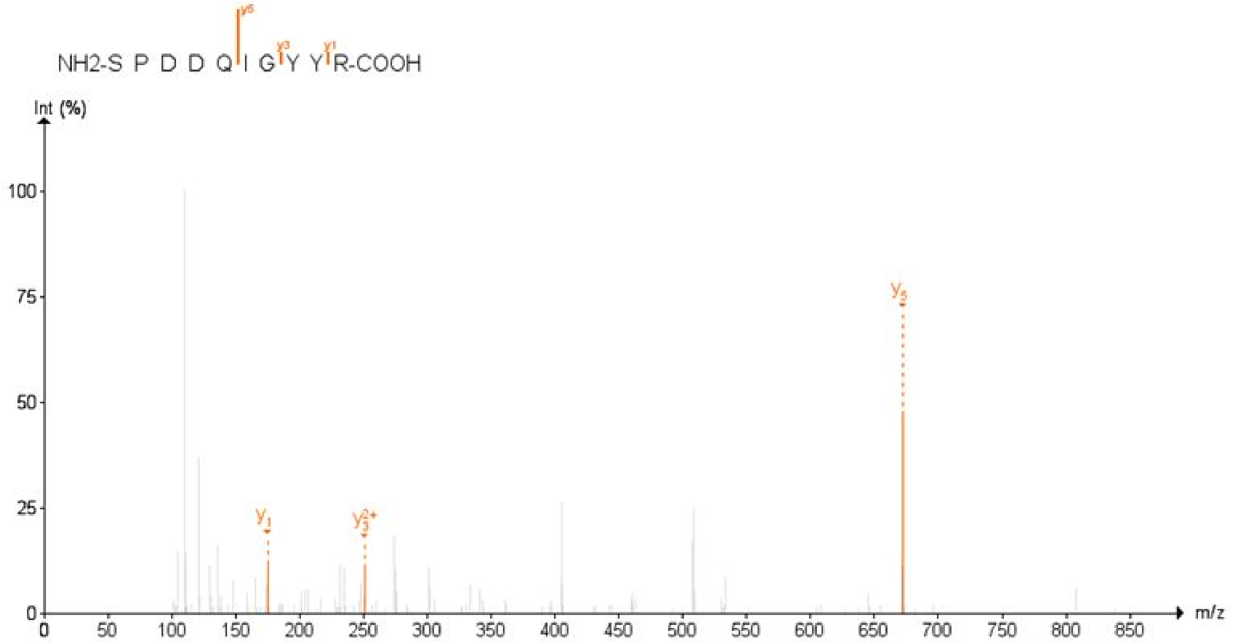

**SVNITFELDER**

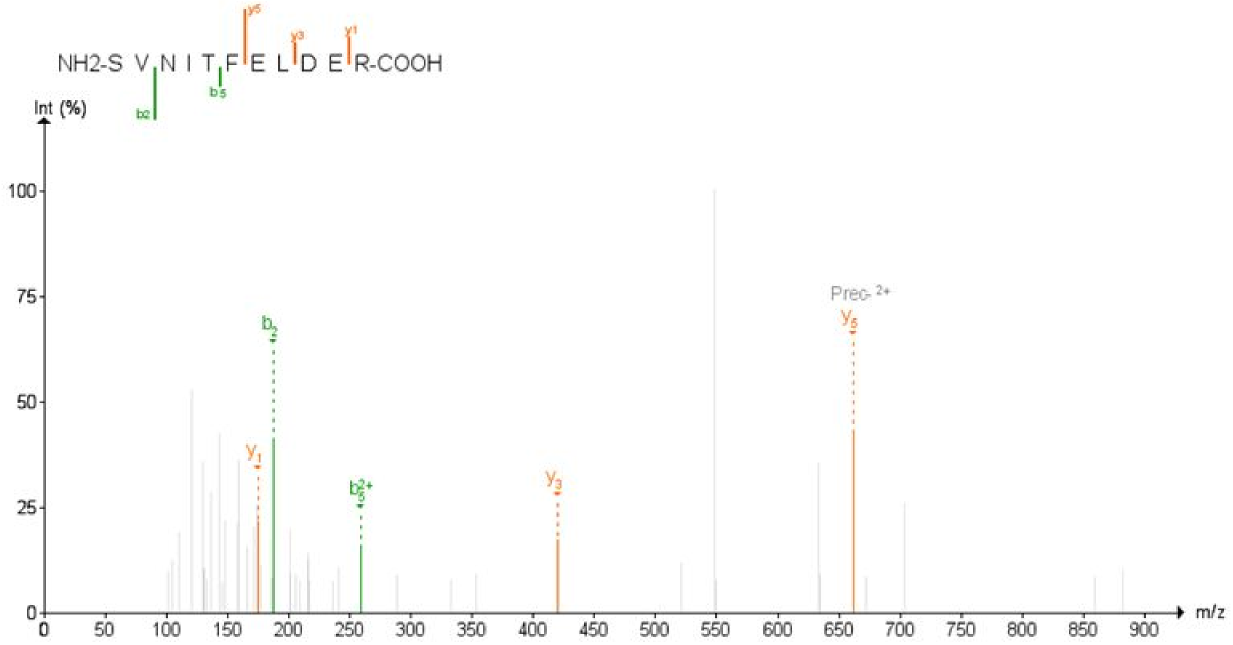

### TAGAAAYYVGYLQPR

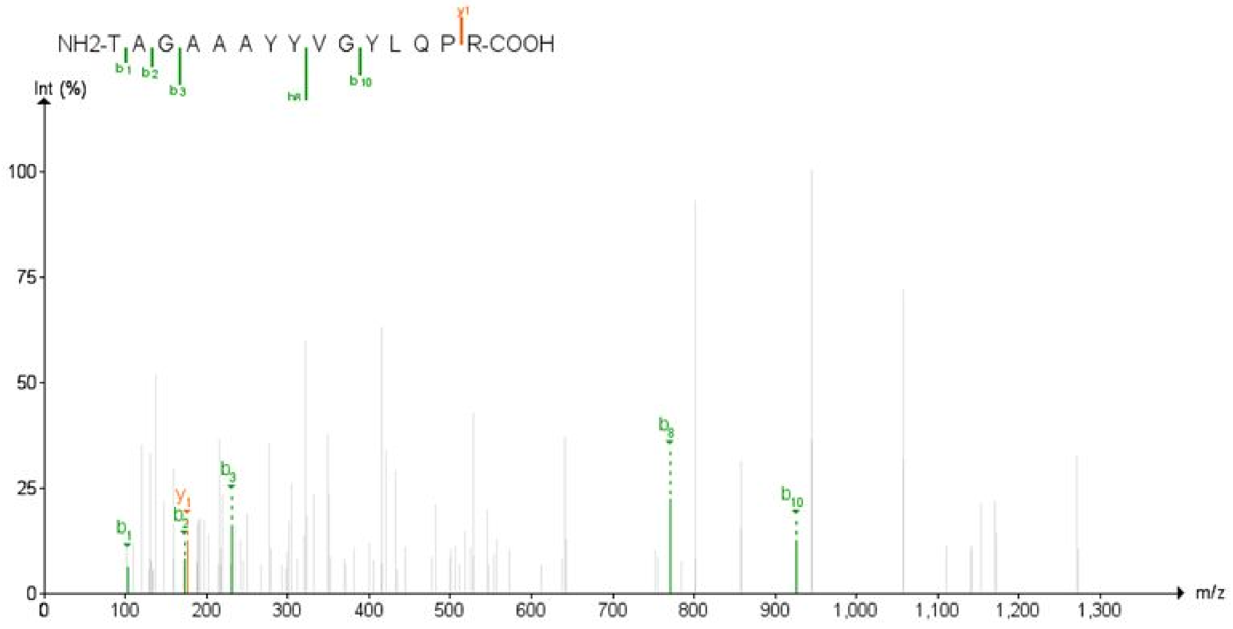

### TALTQHGKEDLKFPR

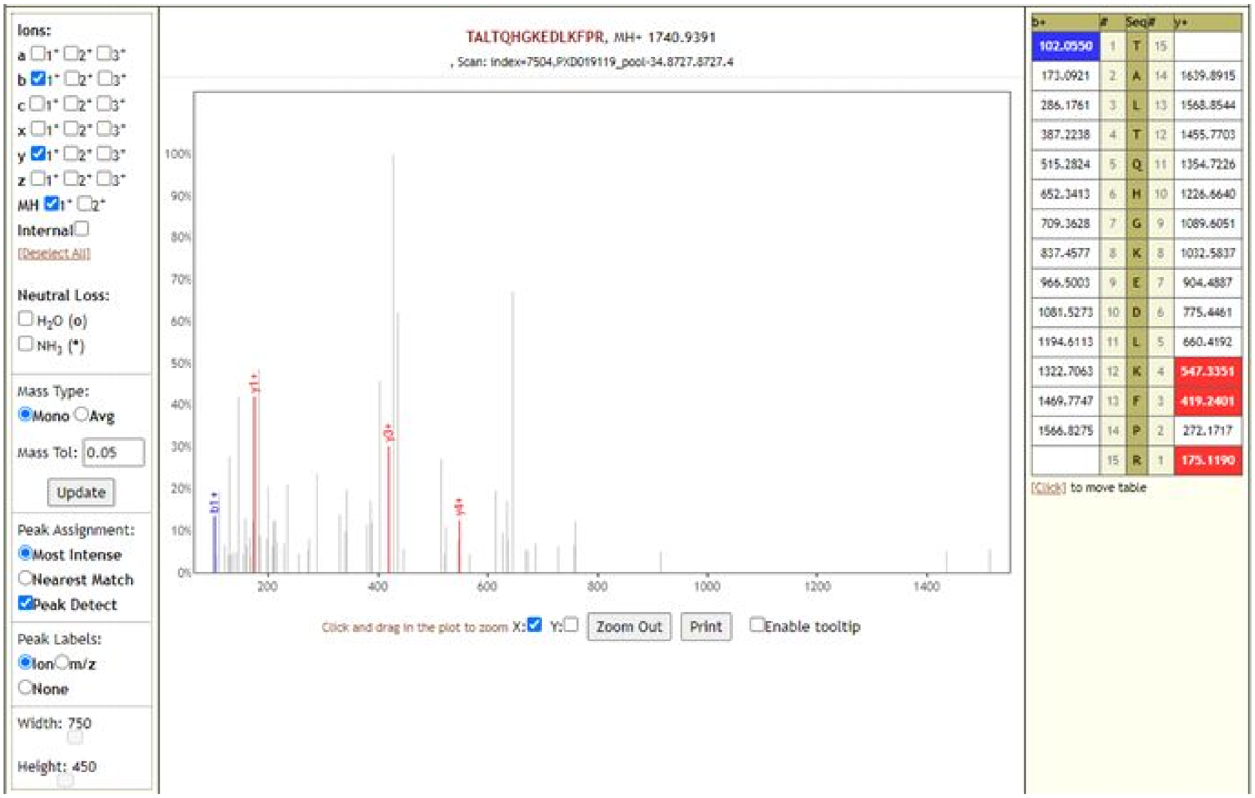

**TLSYYKLGASQRVAGDSGFAAYSR**

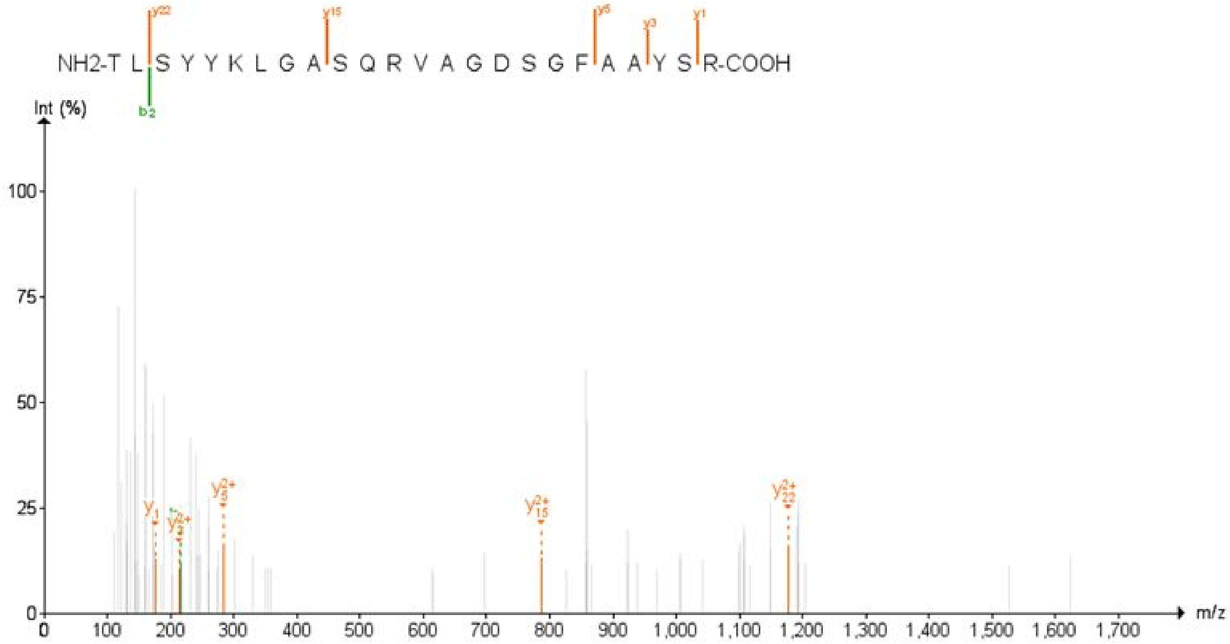

**AFQLTPIAVQMTK**

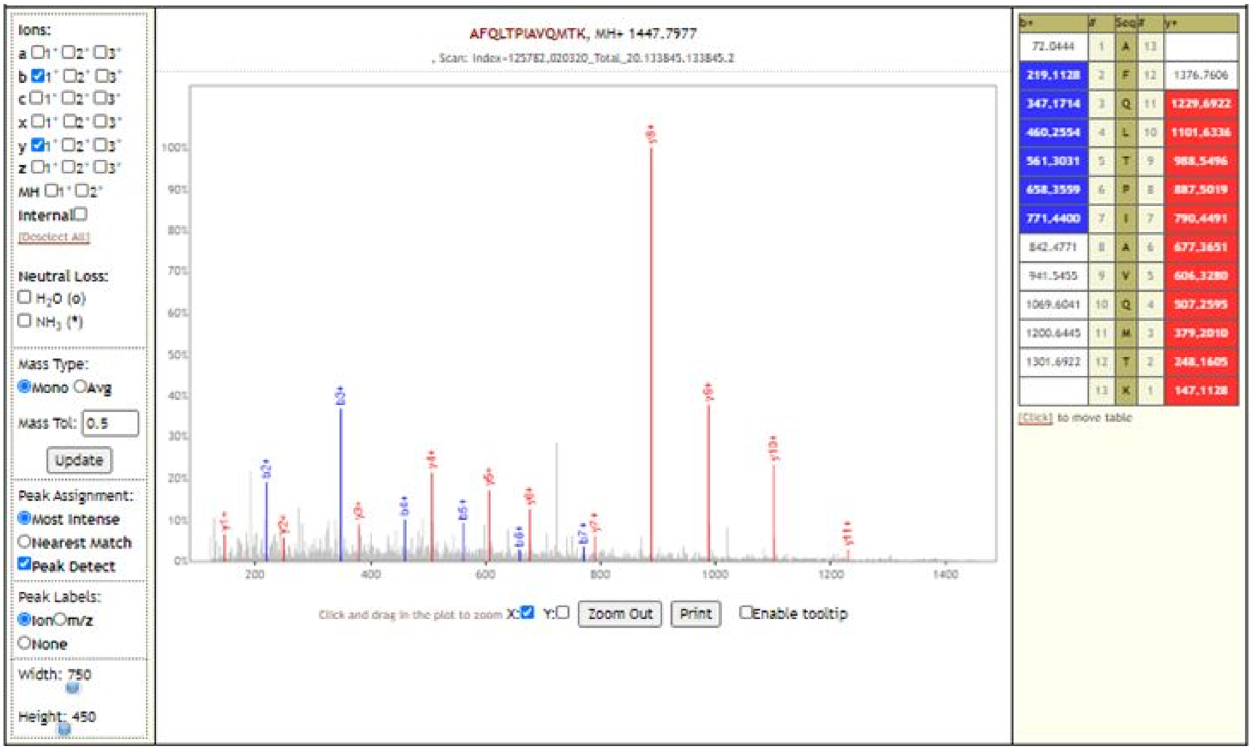

### AFQLTPIAVQMTKLATTEELPDEFVVVTVK

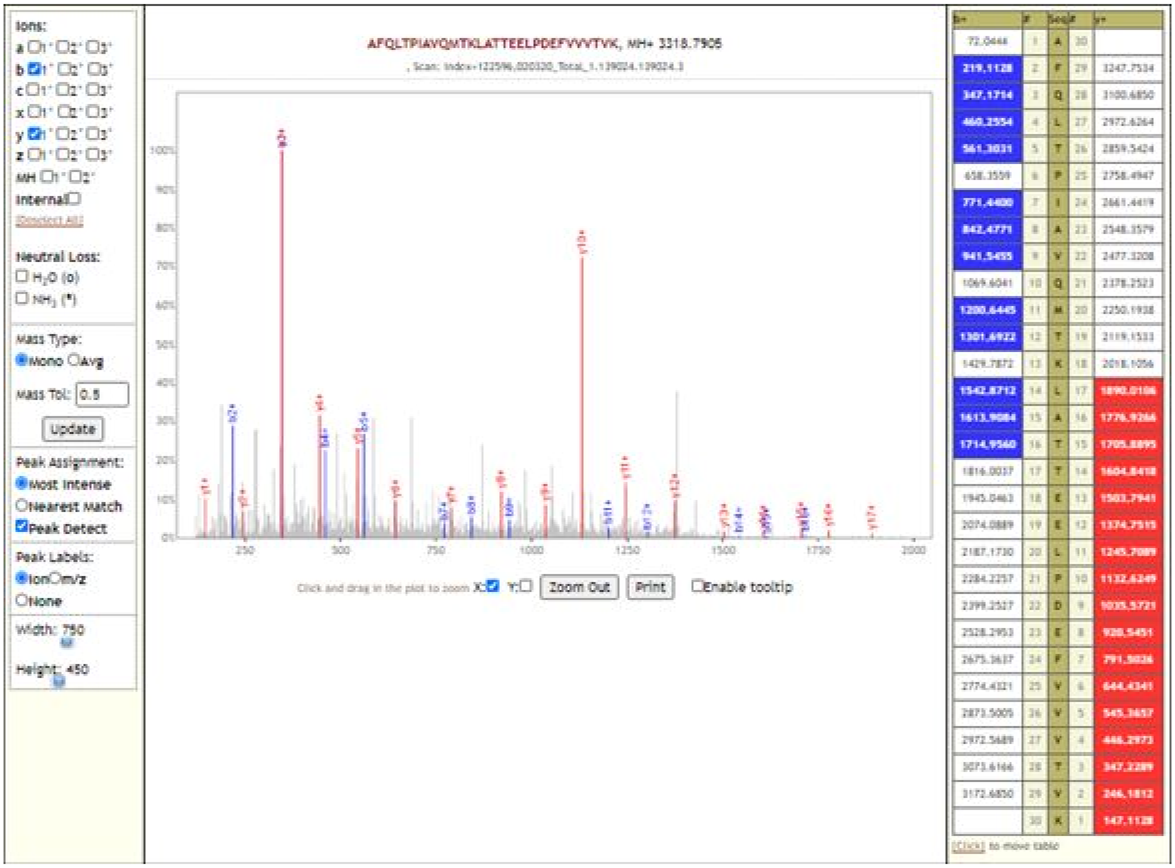

| b+ | # | Seq | # | y+ |
|---|---|---|---|---|
| 72.0444 | 1 | A | 30 | |
| 219.1128 | 2 | F | 29 | 3247.7534 |
| 347.1714 | 3 | Q | 28 | 3100.6850 |
| 460.2554 | 4 | L | 27 | 2972.6264 |
| 561.3031 | 5 | T | 26 | 2859.5424 |
| 658.3559 | 6 | P | 25 | 2758.4947 |
| 771.4400 | 7 | I | 24 | 2661.4419 |
| 842.4771 | 8 | A | 23 | 2548.3579 |
| 941.5455 | 9 | V | 22 | 2477.3208 |
| 1069.6041 | 10 | Q | 21 | 2378.2523 |
| 1200.6445 | 11 | M | 20 | 2250.1938 |
| 1301.6922 | 12 | T | 19 | 2119.1533 |
| 1429.7872 | 13 | K | 18 | 2018.1056 |
| 1542.8712 | 14 | L | 17 | 1890.0106 |
| 1613.9084 | 15 | A | 16 | 1776.9266 |
| 1714.9560 | 16 | T | 15 | 1705.8895 |
| 1816.0037 | 17 | T | 14 | 1604.8418 |
| 1945.0463 | 18 | E | 13 | 1503.7941 |
| 2074.0889 | 19 | E | 12 | 1374.7515 |
| 2187.1730 | 20 | L | 11 | 1245.7089 |
| 2284.2257 | 21 | P | 10 | 1132.6249 |
| 2399.2527 | 22 | D | 9 | 1035.5721 |
| 2528.2953 | 23 | E | 8 | 920.5451 |
| 2675.3637 | 24 | F | 7 | 791.5026 |
| 2774.4321 | 25 | V | 6 | 644.4341 |
| 2873.5005 | 26 | V | 5 | 545.3657 |
| 2972.5689 | 27 | V | 4 | 446.2973 |
| 3073.6166 | 28 | T | 3 | 347.2289 |
| 3172.6850 | 29 | V | 2 | 246.1812 |
| | 30 | K | 1 | 147.1128 |

### ALNDFSNSGSDVLYQPPQTSITSAVLQSGFR

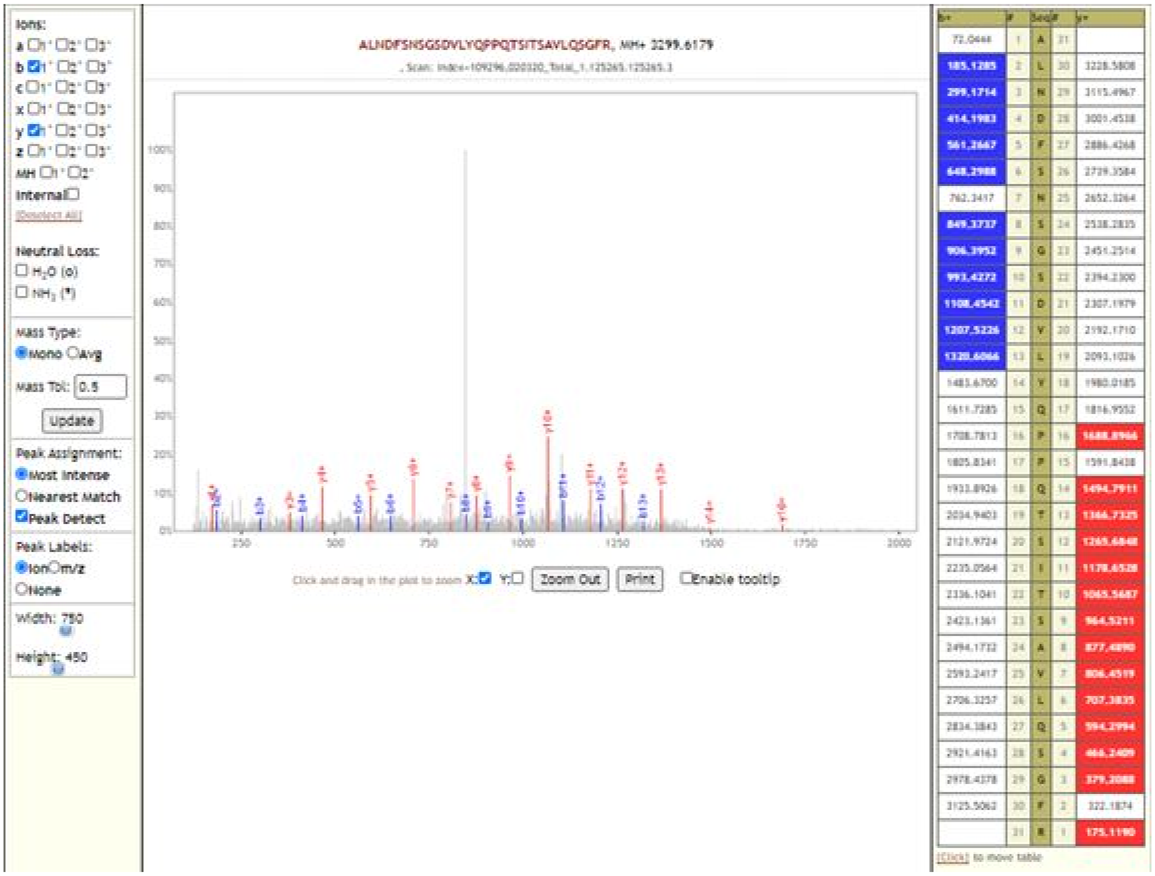

### DQNNVGPKVYPIILR

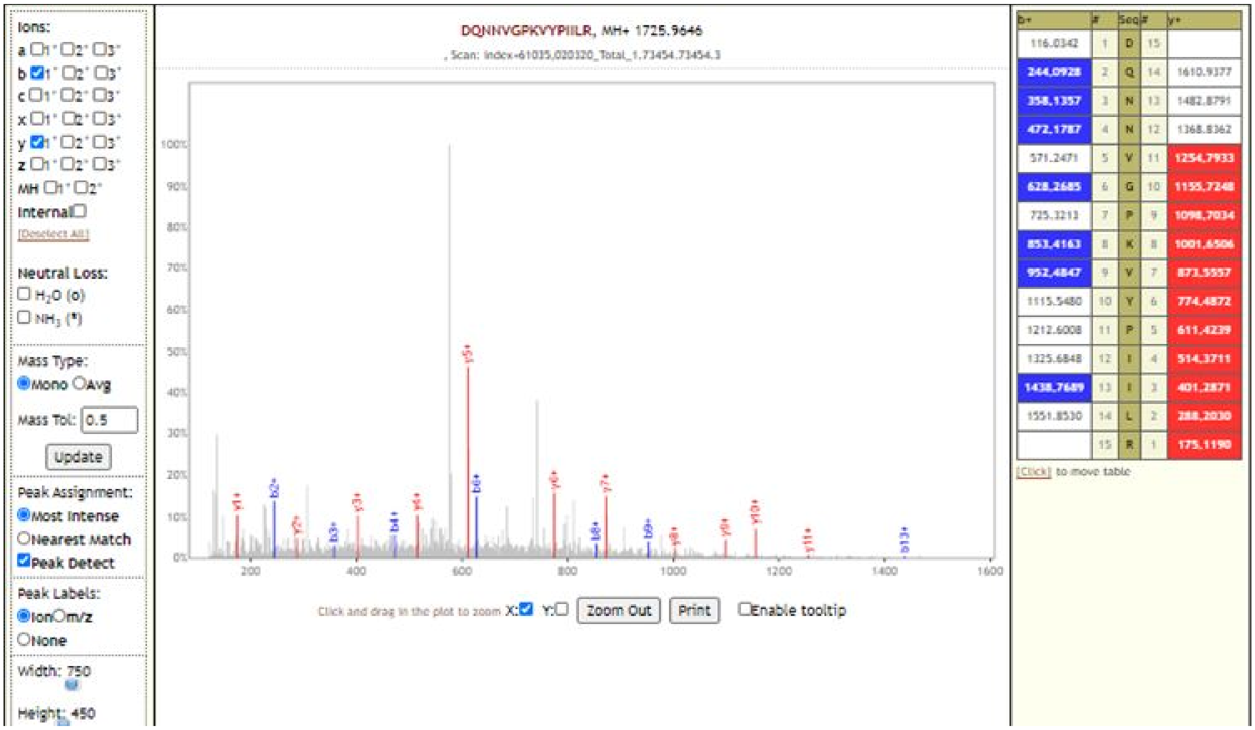

### ISEMHPALR

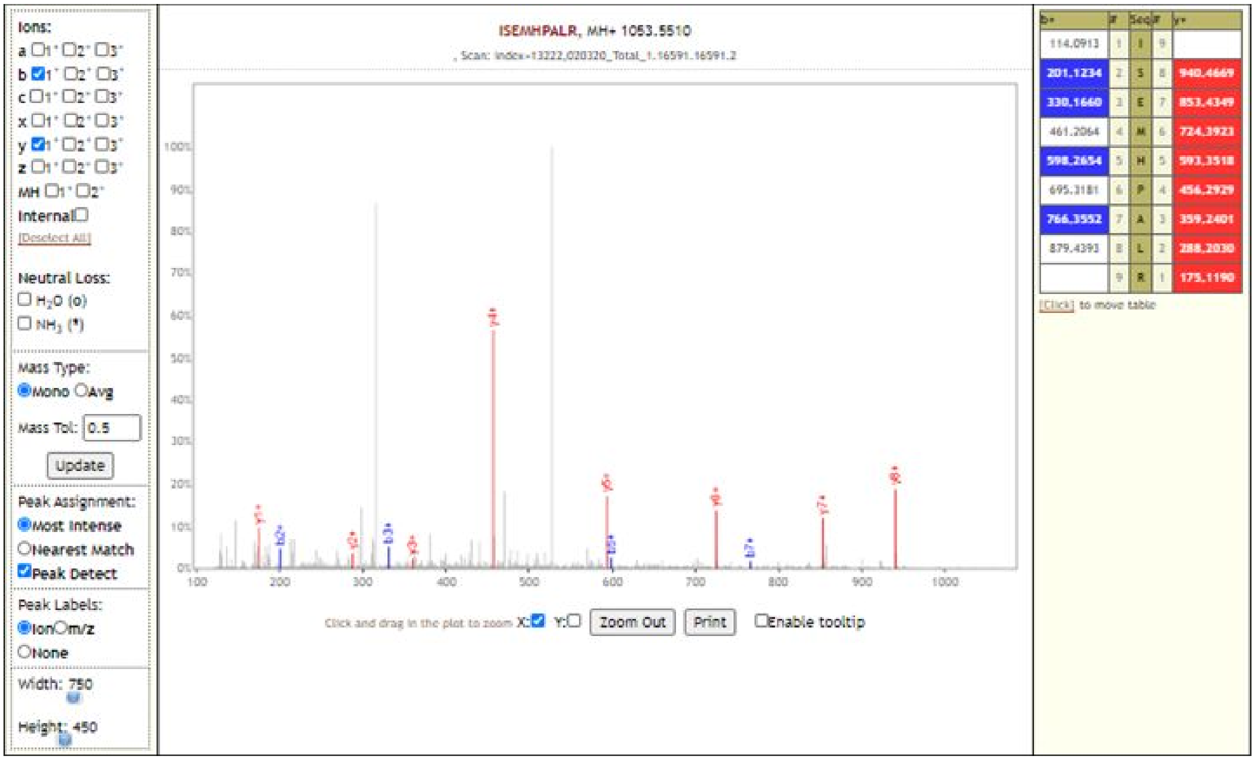

### KTLNSLEDK

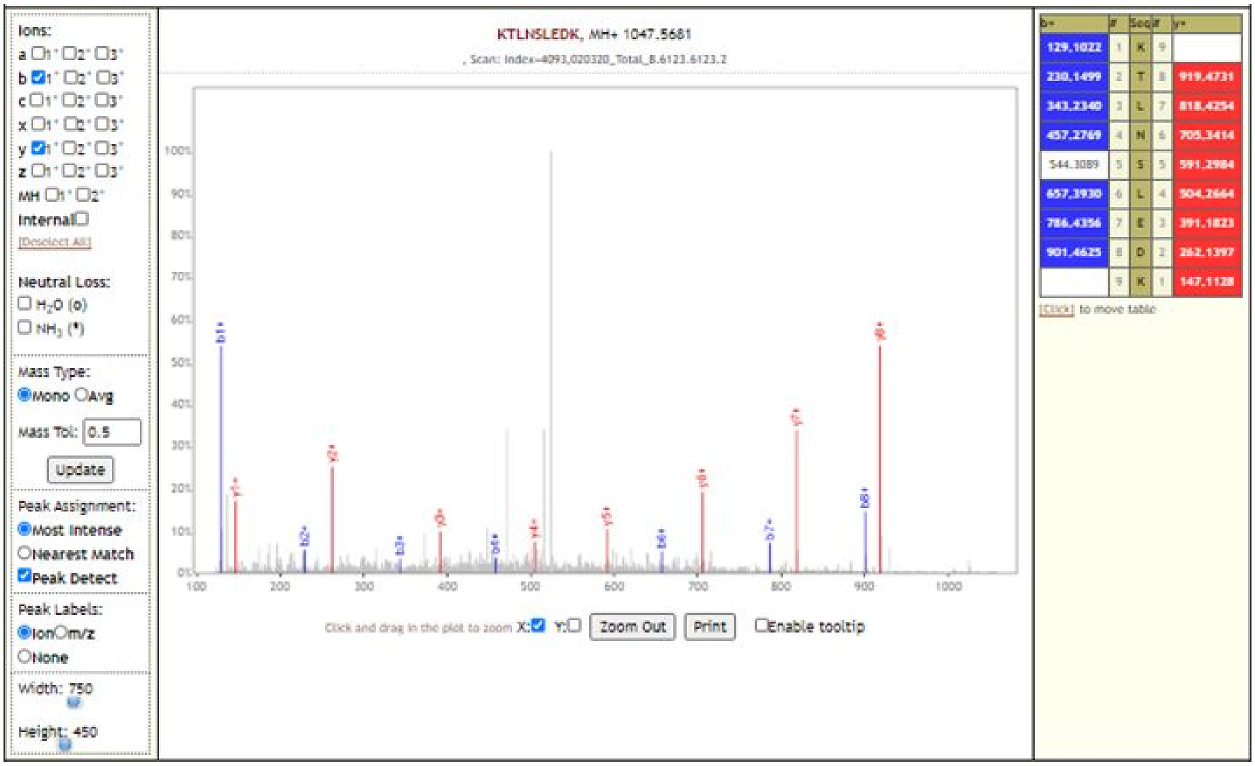

### KTLNSLEDKAFQLTPIAVQMTK

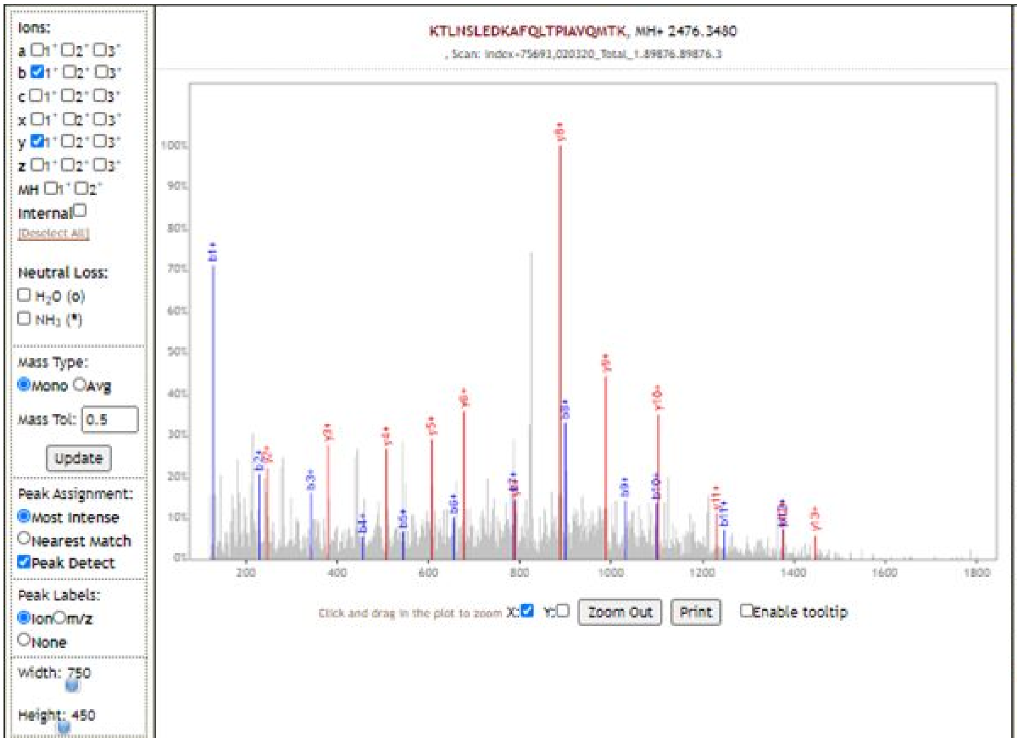

| b+ | # | Seq | # | y+ |
|---|---|---|---|---|
| 129.1022 | 1 | K | 22 | |
| 230.1499 | 2 | T | 21 | 2348.2530 |
| 343.2340 | 3 | L | 20 | 2247.2053 |
| 457.2769 | 4 | N | 19 | 2134.1213 |
| 544.3089 | 5 | S | 18 | 2020.0783 |
| 657.3930 | 6 | L | 17 | 1933.0463 |
| 786.4356 | 7 | E | 16 | 1819.9622 |
| 901.4625 | 8 | D | 15 | 1690.9197 |
| 1029.5575 | 9 | K | 14 | 1575.8927 |
| 1100.5946 | 10 | A | 13 | 1447.7977 |
| 1247.6630 | 11 | F | 12 | 1376.7606 |
| 1375.7216 | 12 | Q | 11 | 1229.6922 |
| 1488.8057 | 13 | L | 10 | 1101.6336 |
| 1589.8533 | 14 | T | 9 | 988.5496 |
| 1686.9061 | 15 | P | 8 | 887.5019 |
| 1799.9902 | 16 | I | 7 | 790.4491 |
| 1871.0273 | 17 | A | 6 | 677.3651 |
| 1970.0957 | 18 | V | 5 | 606.3280 |
| 2098.1543 | 19 | Q | 4 | 507.2595 |
| 2229.1948 | 20 | M | 3 | 379.2010 |
| 2330.2424 | 21 | T | 2 | 248.1605 |
| | 22 | K | 1 | 147.1128 |

[Click] to move table

### LATTEELPDEFVVVTVK

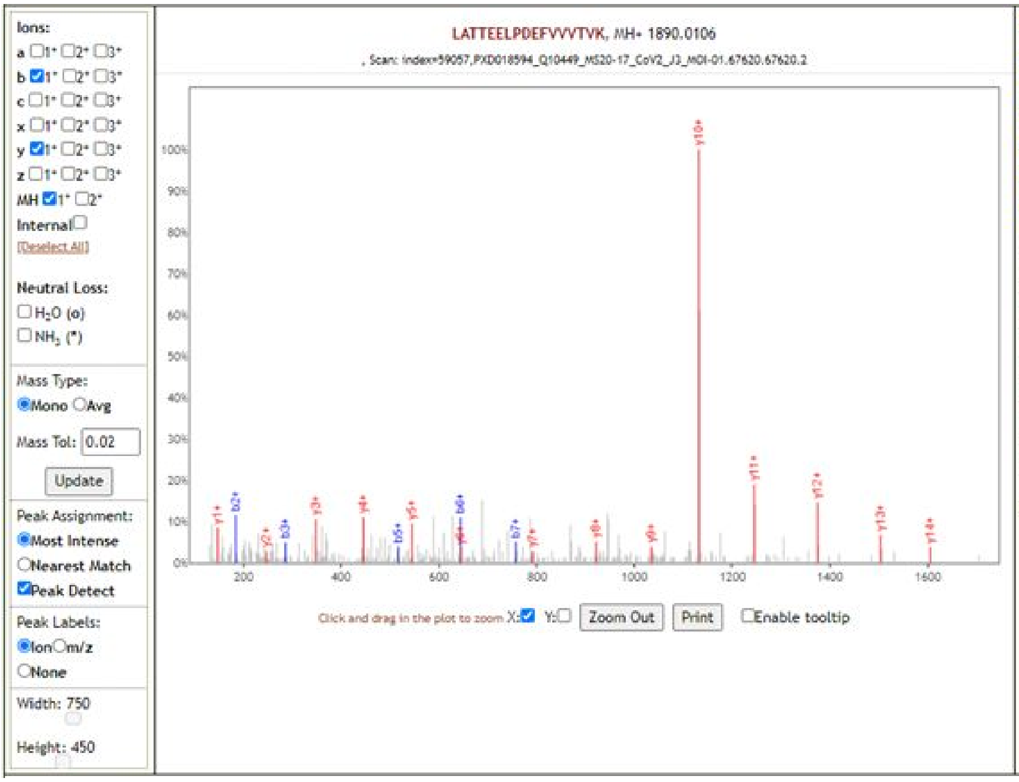

| b+ | # | Seq | # | y+ |
|---|---|---|---|---|
| 114.0913 | 1 | L | 17 | |
| 185.1285 | 2 | A | 16 | 1776.9266 |
| 286.1761 | 3 | T | 15 | 1705.8895 |
| 387.2238 | 4 | T | 14 | 1604.8418 |
| 516.2664 | 5 | E | 13 | 1503.7941 |
| 645.3090 | 6 | E | 12 | 1374.7515 |
| 758.3931 | 7 | L | 11 | 1245.7089 |
| 855.4458 | 8 | P | 10 | 1132.6249 |
| 970.4728 | 9 | D | 9 | 1035.5721 |
| 1099.5154 | 10 | E | 8 | 920.5451 |
| 1246.5838 | 11 | F | 7 | 791.5026 |
| 1345.6522 | 12 | V | 6 | 644.4341 |
| 1444.7206 | 13 | V | 5 | 545.3657 |
| 1543.7890 | 14 | V | 4 | 446.2973 |
| 1644.8367 | 15 | T | 3 | 347.2289 |
| 1743.9051 | 16 | V | 2 | 246.1812 |
| | 17 | K | 1 | 147.1128 |

[Click] to move table

**LGSPLSLNMAR**

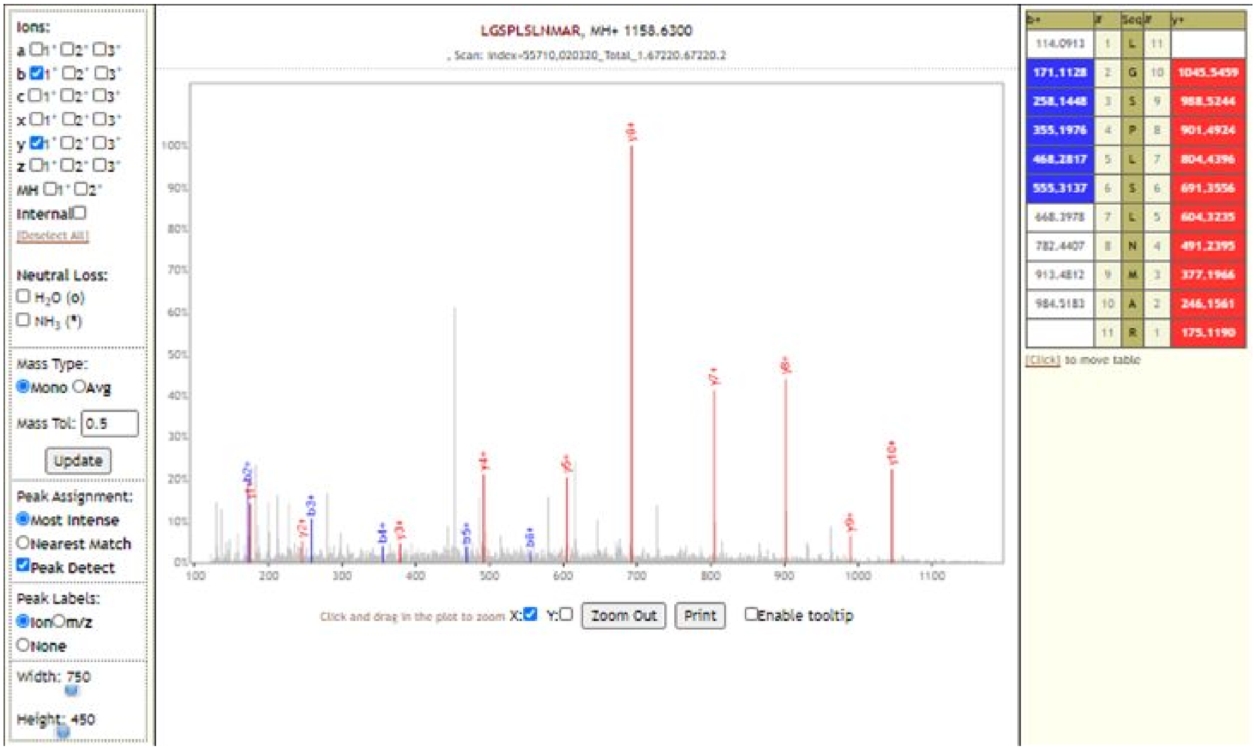

| b+ | # | Seq | # | y+ |
|---|---|---|---|---|
| 114.0913 | 1 | L | 11 | |
| 171.1128 | 2 | G | 10 | 1045.5459 |
| 258.1448 | 3 | S | 9 | 988.5244 |
| 355.1976 | 4 | P | 8 | 901.4924 |
| 468.2817 | 5 | L | 7 | 804.4396 |
| 555.3137 | 6 | S | 6 | 691.3556 |
| 668.3978 | 7 | L | 5 | 604.3235 |
| 782.4407 | 8 | N | 4 | 491.2395 |
| 913.4812 | 9 | M | 3 | 377.1966 |
| 984.5183 | 10 | A | 2 | 246.1561 |
| | 11 | R | 1 | 175.1190 |

**LQAGNATEVPANSTVLSFCAFAVDAAK**

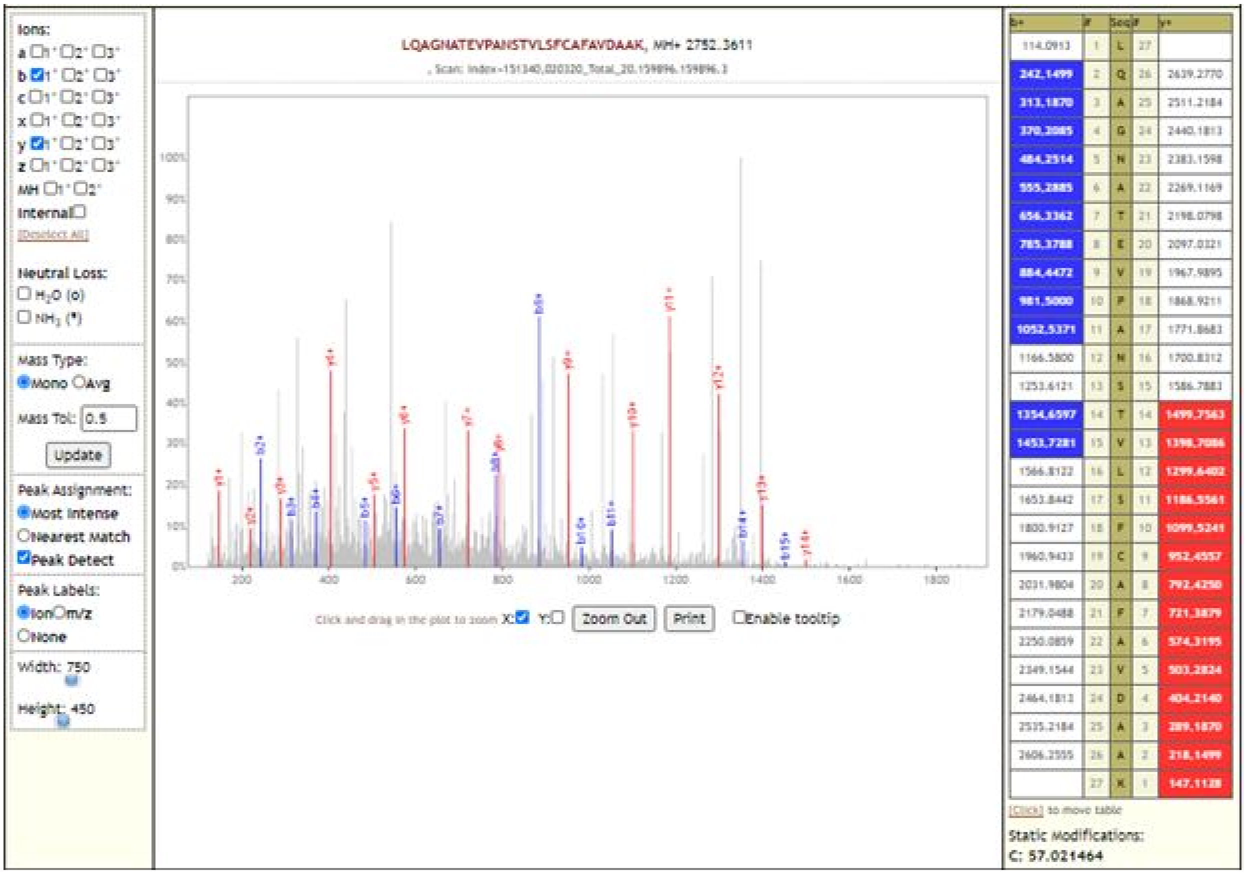

| b+ | # | Seq | # | y+ |
|---|---|---|---|---|
| 114.0913 | 1 | L | 27 | |
| 242.1499 | 2 | Q | 26 | 2639.2770 |
| 313.1870 | 3 | A | 25 | 2511.2184 |
| 370.2085 | 4 | G | 24 | 2440.1813 |
| 484.2514 | 5 | N | 23 | 2383.1598 |
| 555.2885 | 6 | A | 22 | 2269.1169 |
| 656.3362 | 7 | T | 21 | 2198.0798 |
| 785.3788 | 8 | E | 20 | 2097.0321 |
| 884.4472 | 9 | V | 19 | 1967.9895 |
| 981.5000 | 10 | P | 18 | 1868.9211 |
| 1052.5371 | 11 | A | 17 | 1771.8683 |
| 1166.5800 | 12 | N | 16 | 1700.8312 |
| 1253.6121 | 13 | S | 15 | 1586.7883 |
| 1354.6597 | 14 | T | 14 | 1499.7563 |
| 1453.7281 | 15 | V | 13 | 1398.7086 |
| 1566.8122 | 16 | L | 12 | 1299.6402 |
| 1653.8442 | 17 | S | 11 | 1186.5561 |
| 1800.9127 | 18 | F | 10 | 1099.5241 |
| 1960.9433 | 19 | C | 9 | 952.4557 |
| 2031.9804 | 20 | A | 8 | 792.4250 |
| 2179.0488 | 21 | F | 7 | 721.3879 |
| 2250.0859 | 22 | A | 6 | 574.3195 |
| 2349.1544 | 23 | V | 5 | 503.2824 |
| 2464.1813 | 24 | D | 4 | 404.2140 |
| 2535.2184 | 25 | A | 3 | 289.1870 |
| 2606.2555 | 26 | A | 2 | 218.1499 |
| | 27 | K | 1 | 147.1128 |

### LQSLENVAFNVVNK

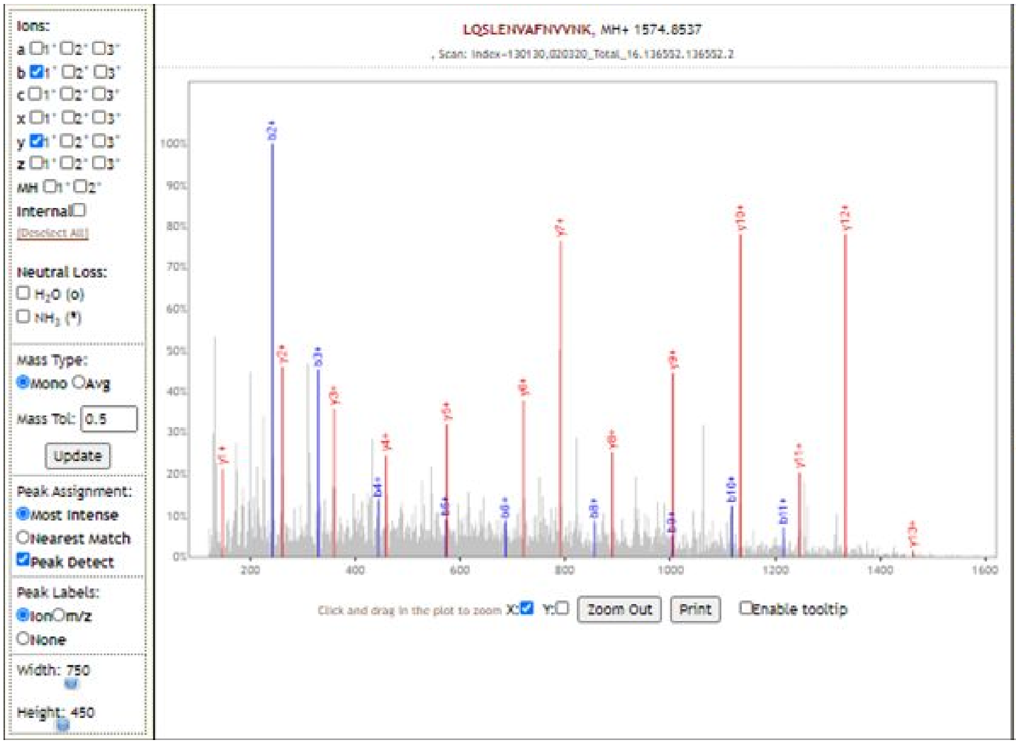

| b+ | # | Seq | # | y+ |
|---|---|---|---|---|
| 114.0913 | 1 | L | 14 | |
| 242.1499 | 2 | Q | 13 | 1461.7696 |
| 329.1819 | 3 | S | 12 | 1333.7110 |
| 442.2660 | 4 | L | 11 | 1246.6790 |
| 571.3086 | 5 | E | 10 | 1133.5949 |
| 685.3515 | 6 | N | 9 | 1004.5524 |
| 784.4199 | 7 | V | 8 | 890.5094 |
| 855.4571 | 8 | A | 7 | 791.4410 |
| 1002.5255 | 9 | F | 6 | 720.4039 |
| 1116.5684 | 10 | N | 5 | 573.3355 |
| 1215.6368 | 11 | V | 4 | 459.2926 |
| 1314.7052 | 12 | V | 3 | 360.2241 |
| 1428.7482 | 13 | N | 2 | 261.1557 |
| | 14 | K | 1 | 147.1128 |

[Click] to move table

### LVDPQIQLAVTR

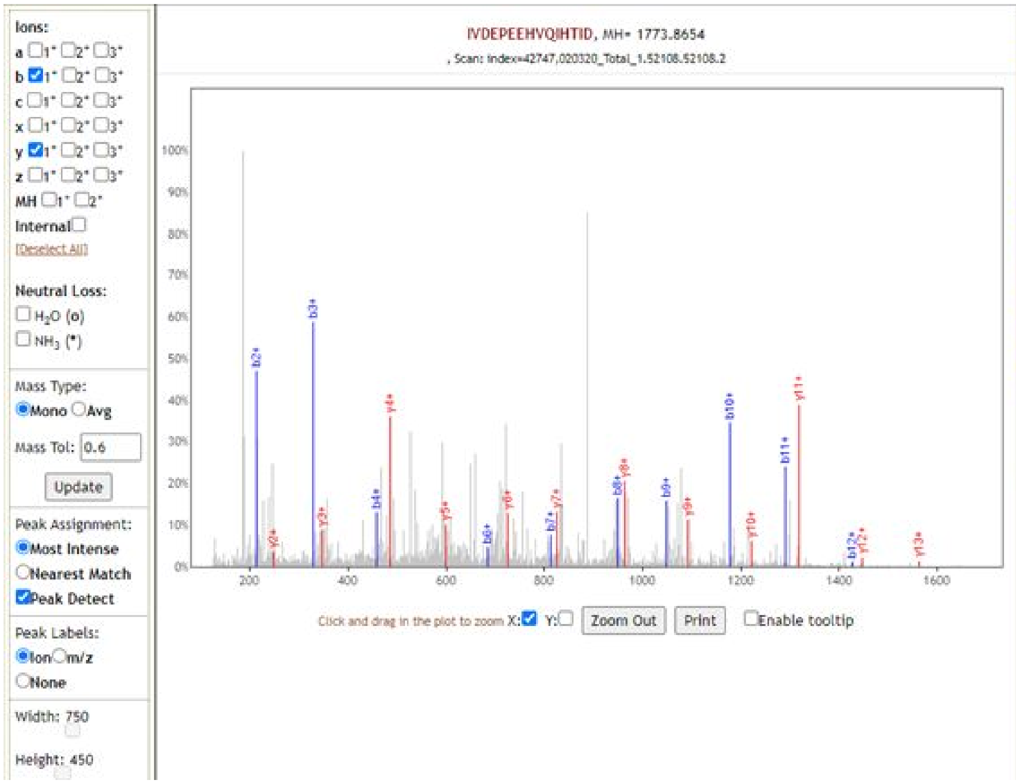

| b+ | # | Seq | # | y+ |
|---|---|---|---|---|
| 114.0913 | 1 | I | 15 | |
| 213.1598 | 2 | V | 14 | 1660.7813 |
| 328.1867 | 3 | D | 13 | 1561.7129 |
| 457.2293 | 4 | E | 12 | 1446.6859 |
| 554.2821 | 5 | P | 11 | 1317.6434 |
| 683.3246 | 6 | E | 10 | 1220.5906 |
| 812.3672 | 7 | E | 9 | 1091.5480 |
| 949.4262 | 8 | H | 8 | 962.5054 |
| 1048.4946 | 9 | V | 7 | 825.4465 |
| 1176.5531 | 10 | Q | 6 | 726.3781 |
| 1289.6372 | 11 | I | 5 | 598.3195 |
| 1426.6961 | 12 | H | 4 | 485.2354 |
| 1527.7438 | 13 | T | 3 | 348.1765 |
| 1640.8279 | 14 | I | 2 | 247.1288 |
| | 15 | D | 1 | 134.0448 |

[Click] to move table

### MENAVGRDQNNVGPK

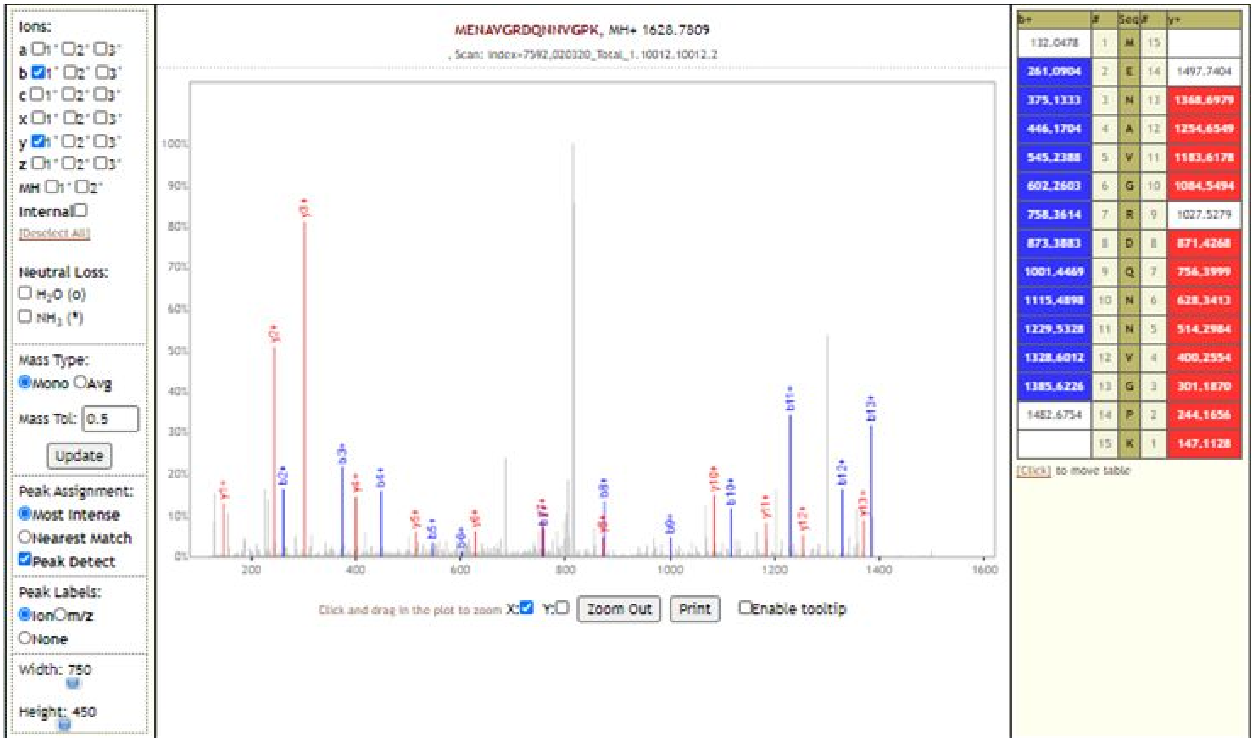

### TLNSLEDK

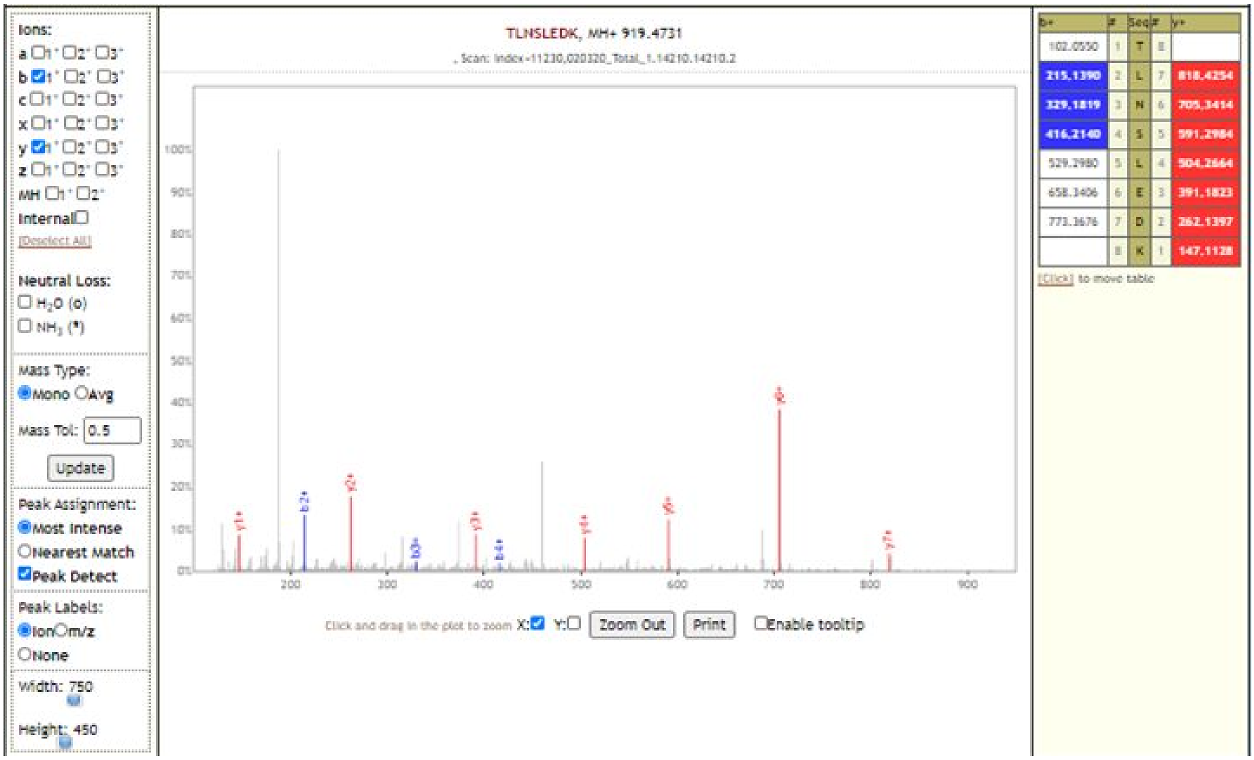

**TLNSLEDKAFQLTPIAVQMTK**

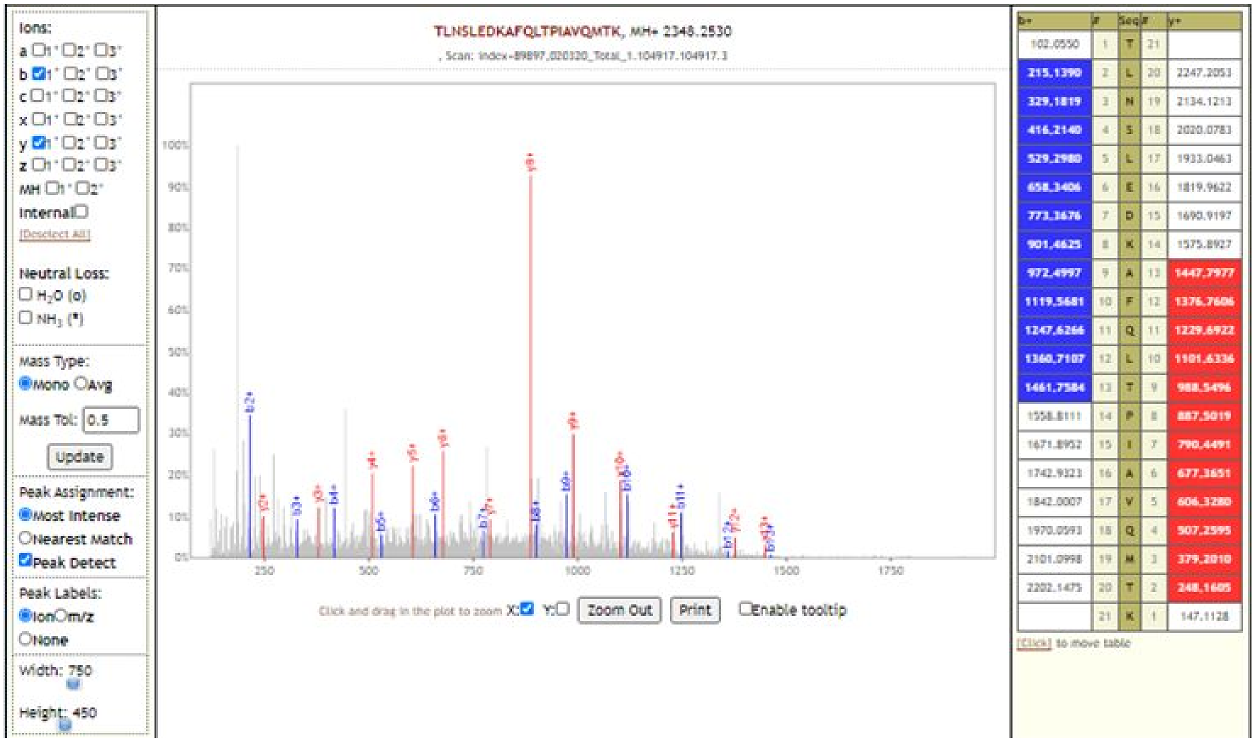

**TLNSLEDKAFQLTPIAVQMTKLATTEELPDEFVVVTVK**

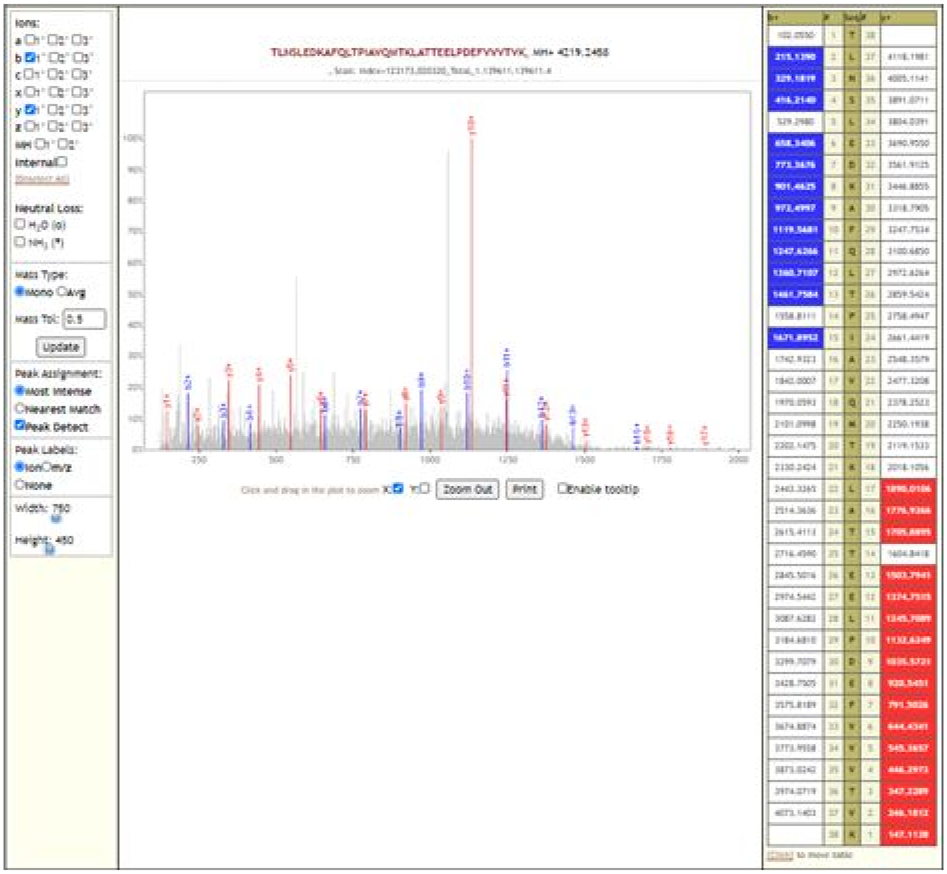

### VYPIILRL

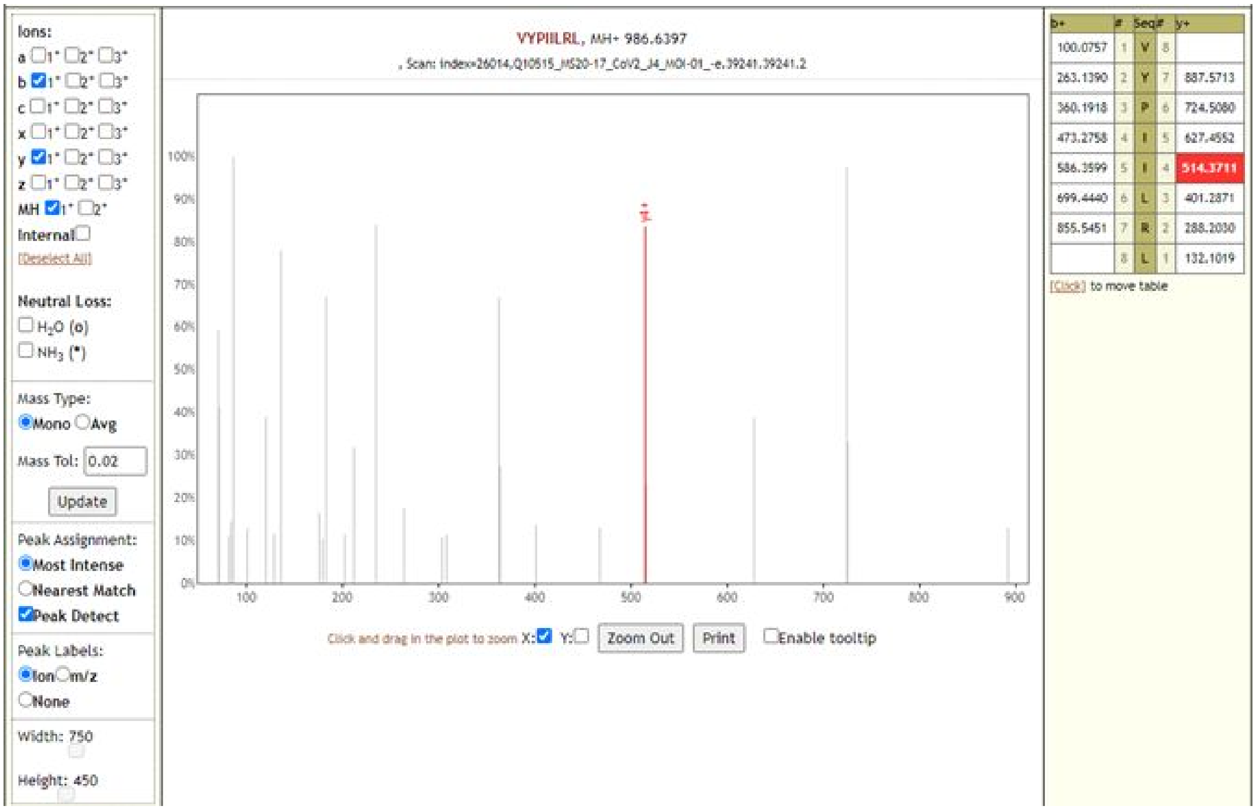

### CVNFNFNGLTGTGVLTESNK

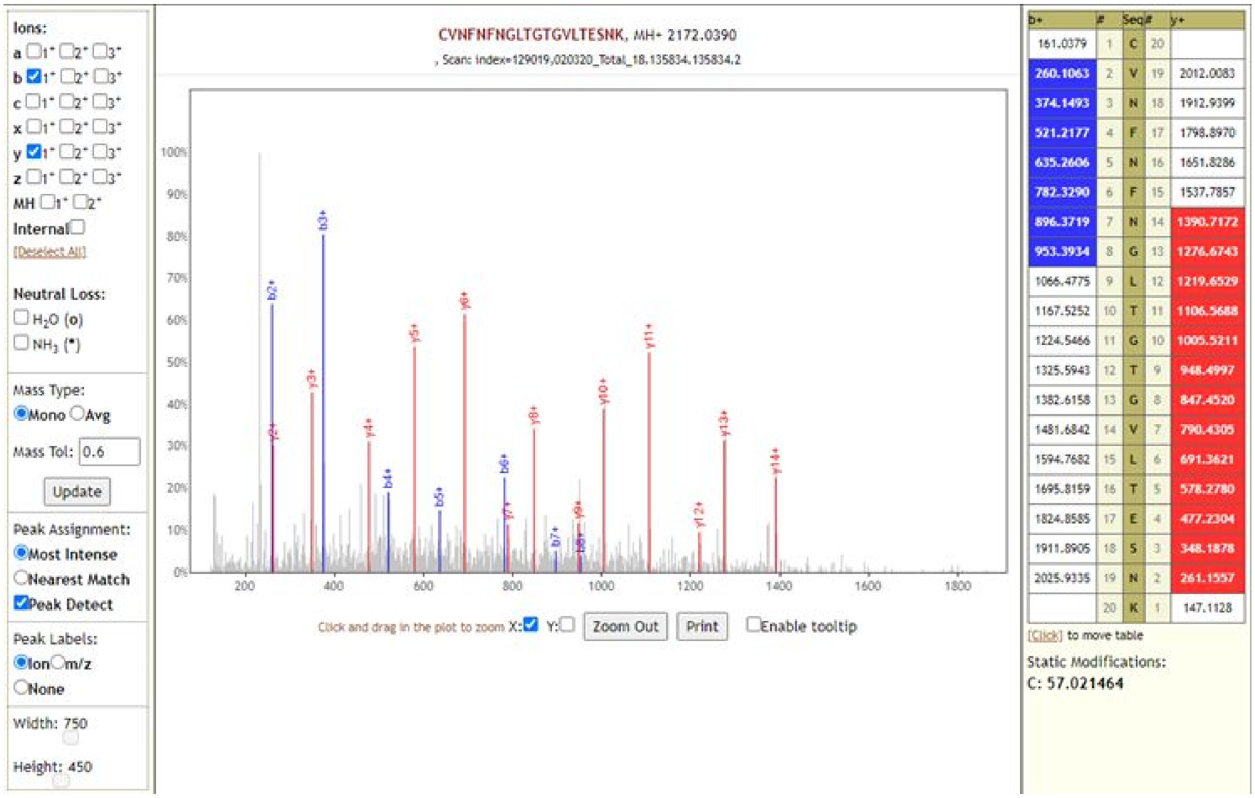

### DIADTTDAVR

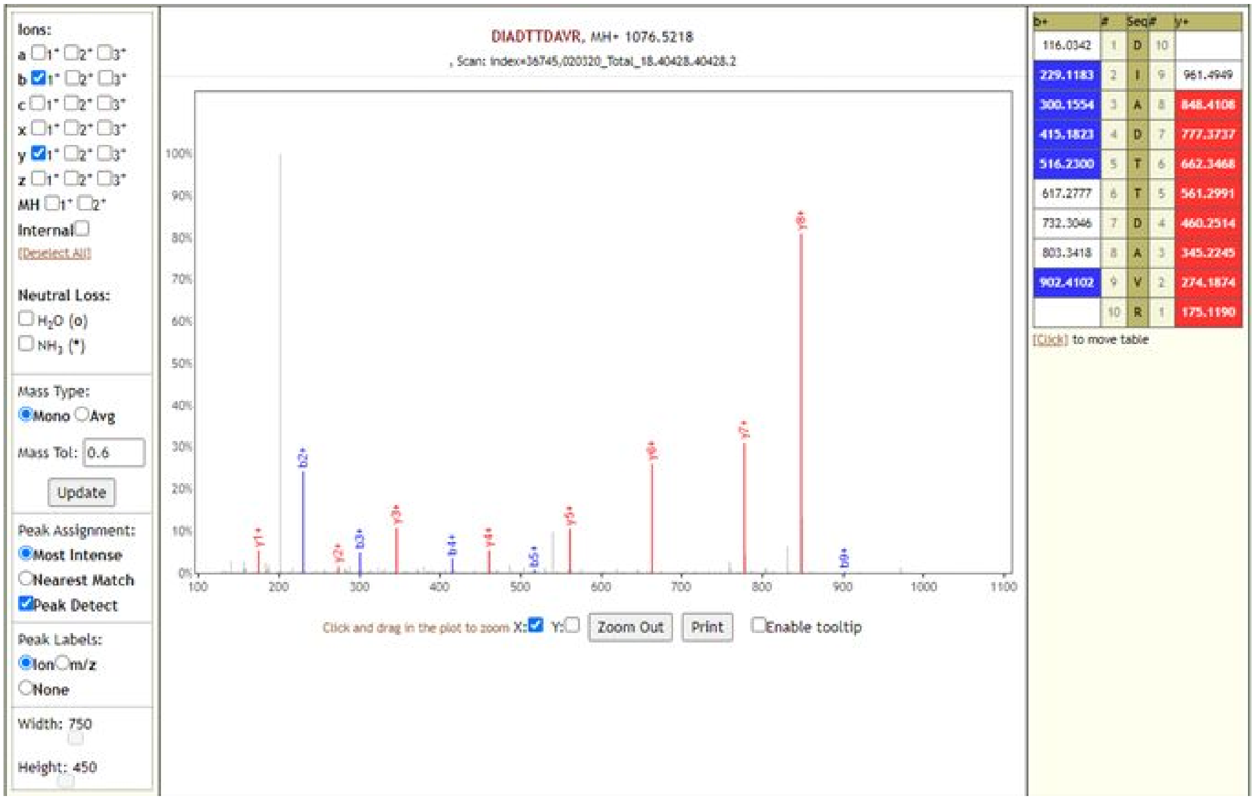

### HTPINLVR

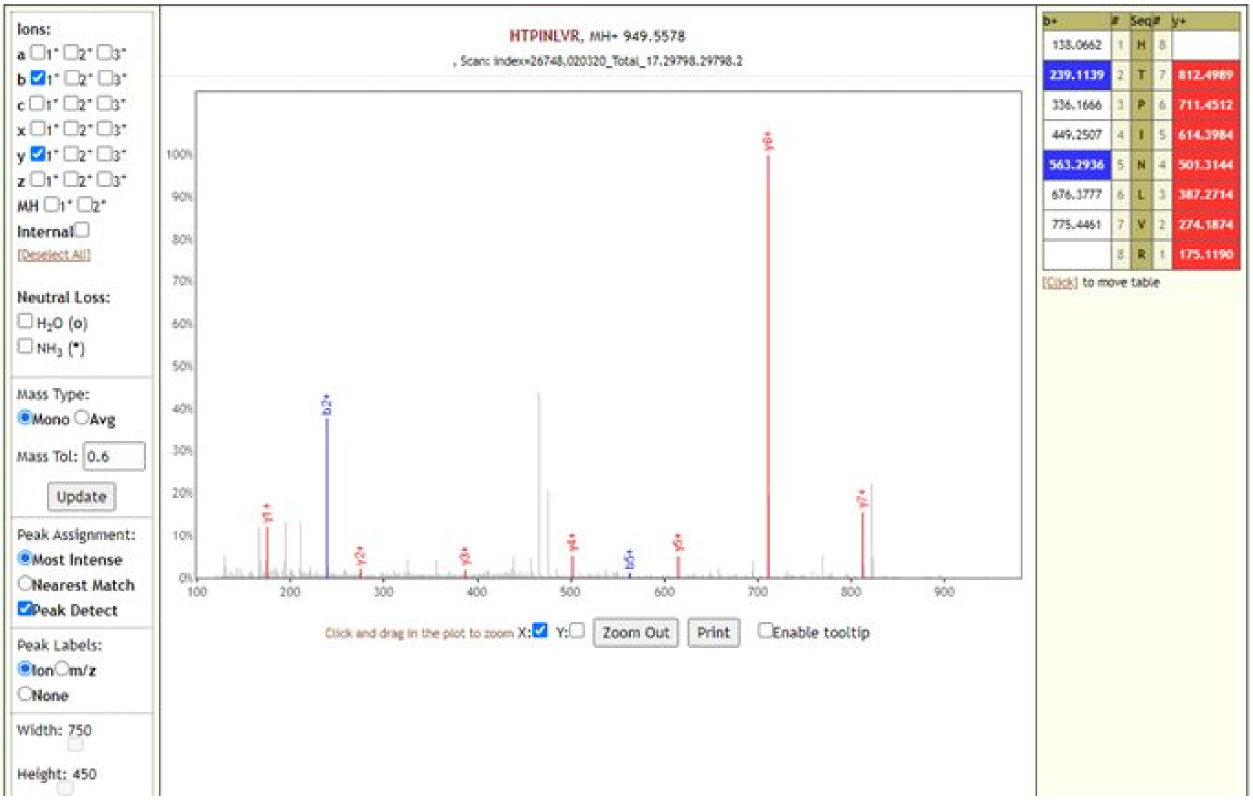

### LPDDFTGCVIAWNSNNLDSK

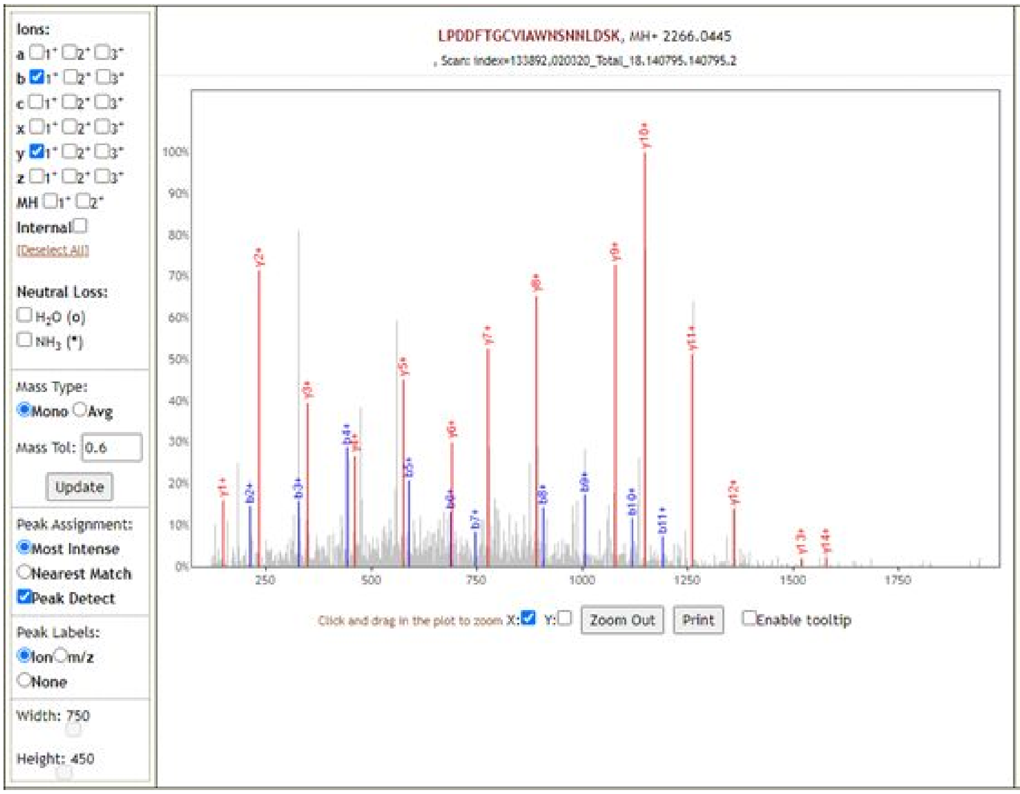

| b+ | # | Seq | # | y+ |
|---|---|---|---|---|
| 114.0913 | 1 | L | 20 | |
| 211.1441 | 2 | P | 19 | 2152.9604 |
| 326.1710 | 3 | D | 18 | 2055.9076 |
| 441.1980 | 4 | D | 17 | 1940.8807 |
| 588.2664 | 5 | F | 16 | 1825.8538 |
| 689.3141 | 6 | T | 15 | 1678.7853 |
| 746.3355 | 7 | G | 14 | 1577.7377 |
| 906.3662 | 8 | C | 13 | 1520.7162 |
| 1005.4346 | 9 | V | 12 | 1360.6856 |
| 1118.5187 | 10 | I | 11 | 1261.6171 |
| 1189.5558 | 11 | A | 10 | 1148.5331 |
| 1375.6351 | 12 | W | 9 | 1077.4960 |
| 1489.6780 | 13 | N | 8 | 891.4167 |
| 1576.7101 | 14 | S | 7 | 777.3737 |
| 1690.7530 | 15 | N | 6 | 690.3417 |
| 1804.7959 | 16 | N | 5 | 576.2988 |
| 1917.8800 | 17 | L | 4 | 462.2558 |
| 2032.9069 | 18 | D | 3 | 349.1718 |
| 2119.9389 | 19 | S | 2 | 234.1448 |
| | 20 | K | 1 | 147.1128 |

[Click] to move table

Static Modifications:
C: 57.021464

### QIAPGQTGK

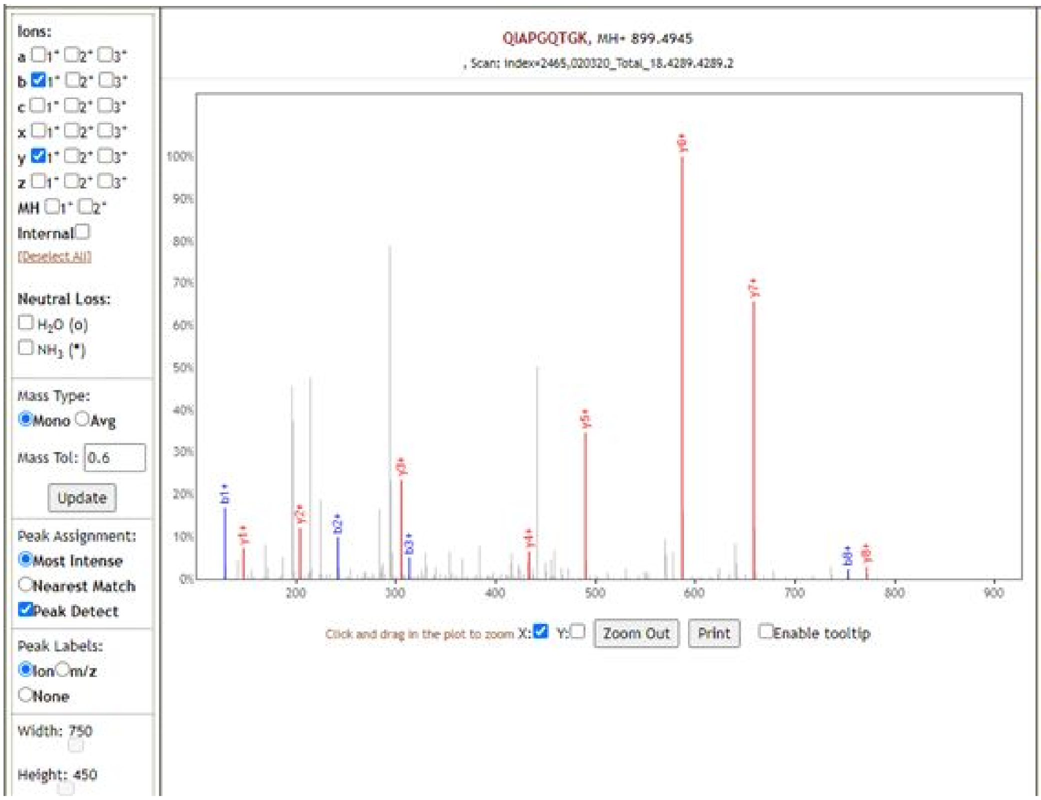

| b+ | # | Seq | # | y+ |
|---|---|---|---|---|
| 129.0659 | 1 | Q | 9 | |
| 242.1499 | 2 | I | 8 | 771.4359 |
| 313.1870 | 3 | A | 7 | 658.3519 |
| 410.2398 | 4 | P | 6 | 587.3148 |
| 467.2613 | 5 | G | 5 | 490.2620 |
| 595.3198 | 6 | Q | 4 | 433.2405 |
| 696.3675 | 7 | T | 3 | 305.1819 |
| 753.3890 | 8 | G | 2 | 204.1343 |
| | 9 | K | 1 | 147.1128 |

[Click] to move table

### TQSLLIVNNATNVVIK

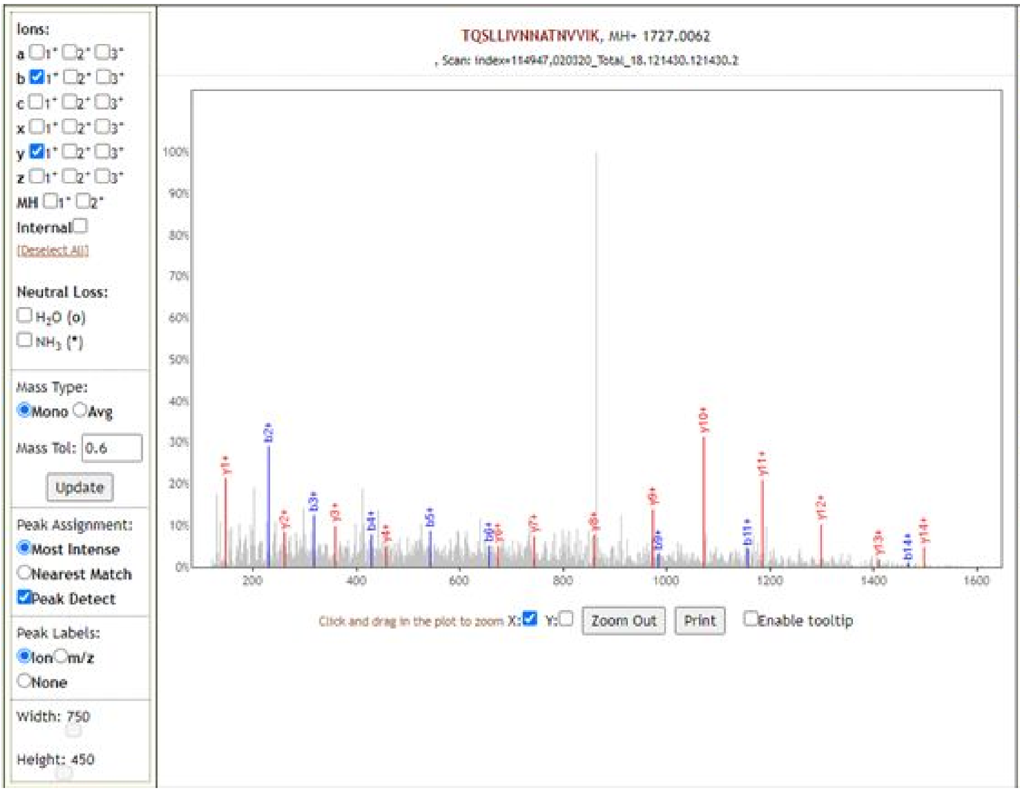

| b+ | # | Seq | # | y+ |
|---|---|---|---|---|
| 102.0550 | 1 | T | 16 | |
| 230.1135 | 2 | Q | 15 | 1625.9585 |
| 317.1456 | 3 | S | 14 | 1497.8999 |
| 430.2296 | 4 | L | 13 | 1410.8679 |
| 543.3137 | 5 | L | 12 | 1297.7838 |
| 656.3978 | 6 | I | 11 | 1184.6997 |
| 755.4662 | 7 | V | 10 | 1071.6157 |
| 869.5091 | 8 | N | 9 | 972.5473 |
| 983.5520 | 9 | N | 8 | 858.5043 |
| 1054.5891 | 10 | A | 7 | 744.4614 |
| 1155.6368 | 11 | T | 6 | 673.4243 |
| 1269.6797 | 12 | N | 5 | 572.3766 |
| 1368.7482 | 13 | V | 4 | 458.3337 |
| 1467.8166 | 14 | V | 3 | 359.2653 |
| 1580.9006 | 15 | I | 2 | 260.1969 |
| | 16 | K | 1 | 147.1128 |

[Click] to move table

### VCEFQFCNDPFLGVYYHK

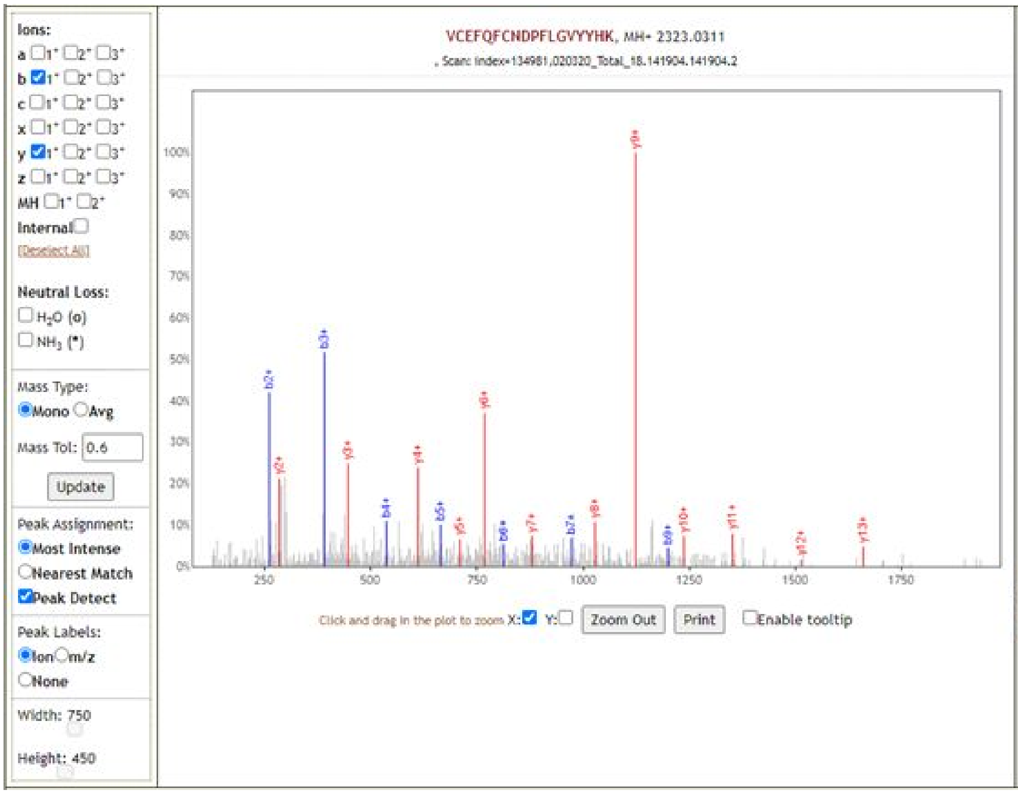

| b+ | # | Seq | # | y+ |
|---|---|---|---|---|
| 100.0757 | 1 | V | 18 | |
| 260.1063 | 2 | C | 17 | 2223.9627 |
| 389.1489 | 3 | E | 16 | 2063.9320 |
| 536.2173 | 4 | F | 15 | 1934.8894 |
| 664.2759 | 5 | Q | 14 | 1787.8210 |
| 811.3443 | 6 | F | 13 | 1659.7624 |
| 971.3750 | 7 | C | 12 | 1512.6940 |
| 1085.4179 | 8 | N | 11 | 1352.6634 |
| 1200.4449 | 9 | D | 10 | 1238.6204 |
| 1297.4976 | 10 | P | 9 | 1123.5935 |
| 1444.5660 | 11 | F | 8 | 1026.5407 |
| 1557.6501 | 12 | L | 7 | 879.4723 |
| 1614.6716 | 13 | G | 6 | 766.3883 |
| 1713.7400 | 14 | V | 5 | 709.3668 |
| 1876.8033 | 15 | Y | 4 | 610.2984 |
| 2039.8666 | 16 | Y | 3 | 447.2350 |
| 2176.9255 | 17 | H | 2 | 284.1717 |
| | 18 | K | 1 | 147.1128 |

[Click] to move table

Static Modifications:
C: 57.021464
C: 57.021464

**YNENGTITDAVDCALDPLSETK**

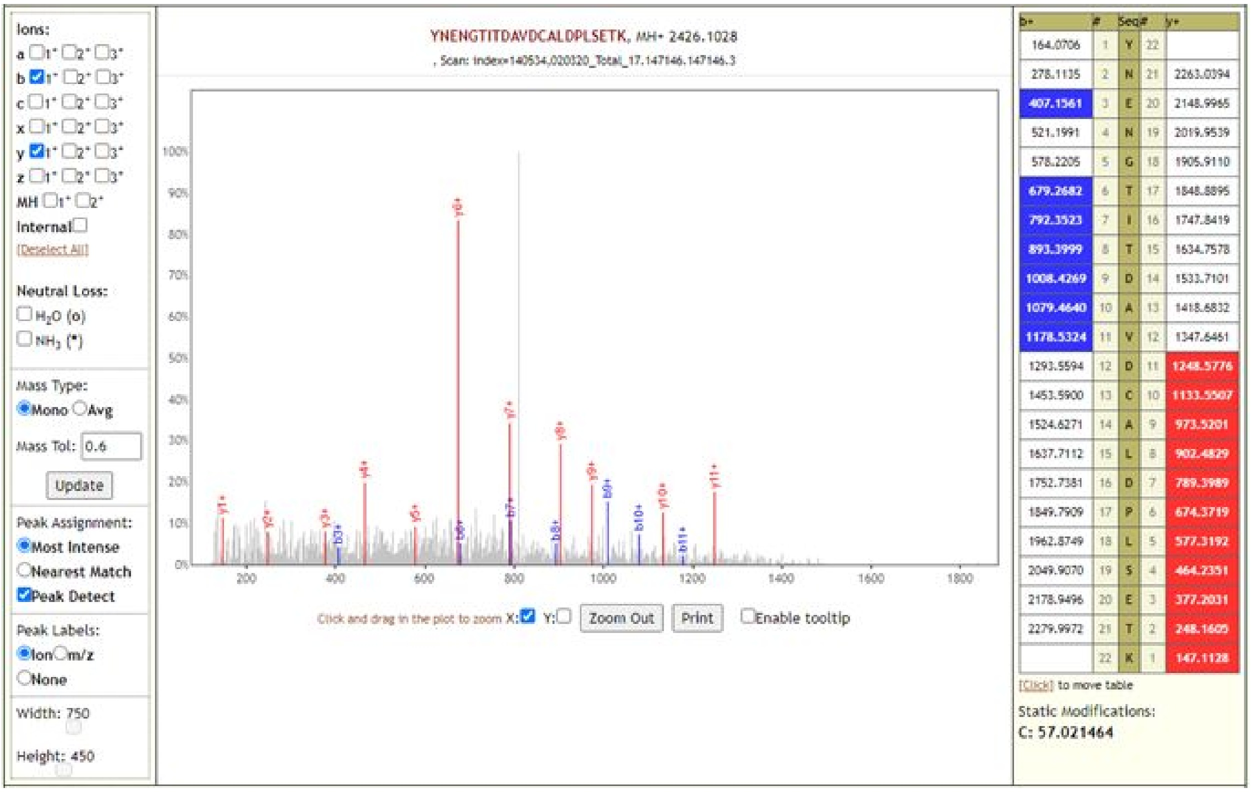

**Supplementary Table 1: The distribution of panel of SARS-CoV-2 peptides detected via sequence database searching workflow and *in silico* analysis.**

The table enlists the datasets in which each of the 639 SARS-CoV-2 peptides were detected. The peptides detected via Search Workflow in cell culture or clinical datasets have been enlisted. Additionally, peptides detected from clinical datasets by running the Validation Workflow against the 639 peptides panel. The table also enlists peptides predicted via *in silico* analysis by Orsburn *et al* (https://doi.org/10.1101/2020.03.08.980383) and those detected by Gouveia *et al* (https://doi.org/10.1002/pmic.202000107) in deep proteomic analysis of CoviD-19 virus infected Vero cells.

| **Peptide** | **Detected via Search Workflow** | **Detected via Validation Workflow** | **Predicted via *in silico* analysis** | **Detected as Target Peptides in the Deep proteomic analysis of CoviD-19 virus infected Vero cells.** |
|---|---|---|---|---|
| AAITILDGISQYSLR | PXD018241 (cell culture) | | | |
| AAIVLQLPQGTTLPK | PXD018804, PXD018594 and PXD018241 (cell culture) | | | |
| AALALLLLDR | PXD018241 (cell culture) | | | |
| AALALLLLDRLNQLESK | PXD018241 (cell culture) | | | |
| AALLADKFPVLHDIGNPK | PXD018241 (cell culture) | | | |
| ACPLIAAVITR | PXD018241 (cell culture) | | | |
| ACVEEVTTTLEETK | PXD018241 (cell culture) | | | |
| ADETQALPQR | PXD018804, PXD018594 and PXD018241 (cell culture); PXD021328 (clinical) | PXD021328 (clinical) | Yes | Yes |
| ADETQALPQRQ | PXD018804 (cell culture) | | | |
| ADETQALPQRQK | PXD018804 and PXD018594 (cell culture) | | | Yes |
| ADETQALPQRQKK | PXD018804 and PXD018594 (cell culture) | | | Yes |
| ADSNGTITVEELK | PXD018241 (cell culture) | | | |
| ADSNGTITVEELKK | PXD018804 and PXD018241 (cell culture) | | | |
| AENVTGLFK | PXD018241 (cell culture) | | | |
| AFDIYNDK | PXD018241 (cell culture) | | | |
| AFDIYNDKVAGFAK | PXD018804 and PXD018594 (cell culture) | | | |
| AFQLTPIAVQMTK | PXD018804, PXD018594 and PXD018241 (cell culture) | | | |
| AFQLTPIAVQMTKLATTEEL PDEFVVVTVK | PXD018241 (cell culture) | | | |
| AGEAANFCALILAYCNK | PXD018241 (Cell culture) | | | |
| AGGTTEMLAK | PXD018241 (Cell culture) | | | |

| | | | | |
|---|---|---|---|---|
| AGNATEVPANSTVLSFCAF AVDAAK | PXD018241 (Cell culture) | | | |
| AGNGGDAALALLLLDR | PXD018241 (Cell culture); PXD021328 (Clinical) | PXD021328 (Clinical) | | |
| AGNGGDAALALLLLDRLNQ LESK | PXD021328 (Clinical) | PXD021328 (Clinical) | | |
| AISSVLNDILSR | PXD018241 (Cell culture) | | | |
| AIVSTIQR | PXD018241 (Cell culture) | | | |
| ALNDFSNSGSDVLYQPPQT SITSAVLQ | PXD018241 (Cell culture) | | | |
| ALNDFSNSGSDVLYQPPQT SITSAVLQSGFR | PXD018241 (Cell culture) | | | |
| ALNLGETFVTHSK | PXD018241 (Cell culture) | PXD021328 (Clinical) | | |
| ALTAESHVDTDLTKPYIK | PXD018241 (Cell culture) | | | |
| ALTGIAVEQDK | PXD018241 (Cell culture) | | | |
| ALTGIAVEQDKNTQEVFAQ VK | PXD018804, PXD018594 and PXD018241 (cell culture) | PXD021328 (clinical) | | |
| ALTQHGKEDLK | PXD018241 (Cell culture) | PXD021328 (Clinical) | | |
| ANNAAIVLQLPQGTTLPK | PXD018241 (Cell culture) | | | |
| APSASAFFGMSR | PXD018241 (Cell culture) | | | |
| ASANIGCNHTGVVGEGSEG LNDNLLEILQK | PXD018241 (Cell culture) | | | |
| ASANLAATK | PXD018804 and PXD018241 (cell culture) | | | |
| ASCTLSEQLDFIDTK | PXD018241 (Cell culture) | | | |
| ASCTLSEQLDFIDTKR | PXD018241 (Cell culture) | | | |
| ASMPTTIAK | PXD018241 (Cell culture) | | | |
| ASWFTALTQHGK | PXD018241 (Cell culture) | | | |
| ASWFTALTQHGKEDLK | PXD018241 (Cell culture) | | | |
| ASYQTQTNSPR | PXD018241 (Cell culture) | | | |
| ATCEFCGTENLTK | PXD018241 (Cell culture) | | | |
| ATNNAMQVESDDYIATNG PLK | PXD018241 (Cell culture) | | | |
| AVDCALDPLSETK | PXD018241 (Cell culture) | | | |
| AVFISPYNSQNAVASK | PXD018241 (Cell culture) | | | |
| AVGACVLCNSQTSLR | PXD018241 (Cell culture) | | | |
| AYKDYLASGGQPITNCVK | PXD018241 (Cell culture) | | | |
| AYNVTQAFGR | PXD018804, PXD018594 and PXD018241 (cell culture); PXD021328, PXD019423 and PXD020493 (clinical) | PXD021328, PXD019423 and PXD020493 (clinical) | | Yes |
| AYNVTQAFGRR | PXD018804 (Cell culture) | | | |

| | | | | |
|---|---|---|---|---|
| CAGSTFISDEVAR | PXD018241 (Cell culture); PXD020493 (Clinical) | | | |
| CDHCGETSWQTGDFVK | PXD018241 (Cell culture) | | | |
| CDIKDLPK | PXD018804, PXD018594 and PXD018241 (cell culture) | | | |
| CDIKDLPKEITVATSR | PXD018241 (Cell culture) | | | |
| CDLQNYGDSATLPK | PXD018241 (Cell culture) | PXD020493 (Clinical) | | |
| CLWSTKPVETSNSFDVLK | PXD018241 (Cell culture) | | | |
| CPAEIVDTVSALVYDNK | PXD018241 (Cell culture) | | | |
| CSFYEDFLEYHDVR | PXD018804 and PXD018241 (cell culture) | | | |
| CTSVVLLSVLQQLR | PXD018241 (Cell culture) | | | |
| CVNFNFNGLTGTGVLTESNK | PXD018241 (Cell culture) | | Yes | |
| CVNFNFNGLTGTGVLTESNKK | PXD018241 (Cell culture) | | | |
| CYGVSPTK | PXD018241 (Cell culture) | | | |
| DAPAHISTIGVCSMTDIAK | PXD018241 (Cell culture) | | | |
| DAPYIVGDVVQEGVLTAVVIPTK | PXD018241 (Cell culture) | | | |
| DAPYIVGDVVQEGVLTAVVIPTKK | PXD018594 (Cell culture) | | | |
| DASGKPVPYCYDTNVLEGSVAYESLRPDTR | PXD018241 (Cell culture) | | | |
| DATPSDFVR | PXD018241 (Cell culture) | | | |
| DDKDPNFKDQVILLNK | PXD018241 (Cell culture); PXD021328 (Clinical) | PXD021328 (Clinical) | | |
| DFGGFNFSQILPDPSKPSK | PXD018241 (Cell culture) | PXD019423 (Clinical) | | |
| DFMSLSEQLR | PXD018241 (Cell culture) | | | |
| DFMSLSEQLRK | PXD018241 (Cell culture) | | | |
| DFYDFAVSK | PXD018241 (Cell culture) | | | |
| DGCVPLNIIPLTTAAK | PXD018241 (Cell culture) | | | |
| DGHVETFYPK | PXD018241 (Cell culture) | | | |
| DGIIWVATEGALNTPK | PXD018804, PXD018594 and PXD018241 (cell culture); PXD021328 and PXD020493 (clinical) | PXD021328 and PXD020493 (clinical) | | |
| DGIIWVATEGALNTPKDHIGTR | PXD018804 and PXD018241 (cell culture) | PXD019423(Clinical) | | |
| DGIIWVATEGALNTPKDHIGTRNPANNAAIVLQLPQGTTLPK | PXD018241 (Cell culture) | | | |
| DGTCGLVEVEK | PXD018241 (Cell culture) | PXD021328 (Clinical) | | |
| DHIGTRNPANNAAIVLQLPQGTTLPK | PXD018804, PXD018594 and PXD018241 (cell culture) | PXD019423 (clinical) | | Yes |

| | | | |
|---|---|---|---|
| DIADTTDAVR | PXD018241 (Cell culture) | | Yes |
| DIADTTDAVRD | PXD018241 (Cell culture) | | |
| DIADTTDAVRDPQ | PXD018241 (Cell culture) | | |
| DIADTTDAVRDPQTLEILDIT PC | PXD018241 (Cell culture) | | |
| DIASTDTCFANK | PXD018241 (Cell culture) | | |
| DLGACIDCSAR | PXD018241 (Cell culture) | | |
| DLPKEITVATSR | PXD018241 (Cell culture) | | |
| DLPQGFSALEPLVDLPIGINI TR | Not Detected | | Yes |
| DLSPRWYFYYLGTGPEAGLP YGANK | PXD018241 (Cell culture) | | |
| DLSPRWYFYYLGTGPEAGLP YGANKDGIIWVATEGALNT PK | PXD018241 (Cell culture) | | |
| DLYDKLQFTSLEIPR | PXD018241 (Cell culture) | | |
| DNSYFTEQPIDLVPNQPYPN ASFDNFK | PXD018241 (Cell culture) | | |
| DPNFKDQVILLNK | PXD018241 (Cell culture) | | |
| DPQTLEILDITPC | PXD018241 (Cell culture) | | |
| DQNNVGPKVYPIILR | PXD018241 (Cell culture) | | |
| DQVILLNK | Not Detected | | Yes |
| DQVILLNKHIDAYK | PXD018241 (Cell culture) | | |
| DSNGTITVEELKK | PXD018241 (Cell culture) | | |
| DVDTDFVNEFYAYLR | PXD018241 (Cell culture) | | |
| DWSYSGQSTQLGIEFLK | PXD018241 (Cell culture) | | |
| DWYDFVENPDILR | PXD018241 (Cell culture) | | |
| DYLASGGQPITNCVK | PXD018241 (Cell culture) | | |
| EAPAHVSTIGVCTMTDIAK | PXD018241 (Cell culture) | | |
| EETGLLMPLKAPK | PXD018241 (Cell culture) | | |
| EEVKPFITESKPSVEQR | PXD018241 (Cell culture) | | |
| EFVFKNIDGYFK | PXD018804 and PXD018241 (cell culture) | | |
| EGATTCGYLPQNAVVK | PXD018241 (Cell culture) | | |
| EGFFTYICGFIQQK | PXD018241 (Cell culture) | | |
| EGIVWVATEGALNTPK | PXD018241 (Cell culture) | | |
| EGIVWVATEGALNTPKDHI GTR | PXD018241 (Cell culture) | | |
| EGQINDMILSLLSK | PXD018241 (Cell culture) | | |

| | | | | |
|---|---|---|---|---|
| EGSSVELK | PXD018241 (Cell culture) | | | |
| EGVFVSNGTHWFVTQR | PXD018804, PXD018594 and PXD018241 (cell culture) | | | |
| EHEHEIAWYTER | PXD018241 (Cell culture) | | | |
| EIDRLNEVAK | PXD018804, PXD018594 and PXD018241 (cell culture) | | | |
| EIIFLEGETLPTEVLTEEVVLK | PXD018241 (Cell culture) | | | |
| EITVATSR | PXD018804, PXD018594 and PXD018241 (cell culture); PXD021328 (clinical) | | | Yes |
| EITVATSRTLSYYK | PXD018804 and PXD018594 (Cell culture) | | | Yes |
| EITVATSRTLSYYKLGASQR | PXD018241 (Cell culture) | | | |
| EKVNINIVGDFK | PXD018241 (Cell culture) | | | |
| ELGVVHNQDVNLHSSR | PXD018241 (Cell culture) | | | |
| ELLQNGMNGR | PXD018241 (Cell culture) | | | |
| ELLVYAADPAMHAASGNLLLDKR | PXD018241 (Cell culture) | | | |
| ELYHYQECVR | PXD018241 (Cell culture) | | | |
| EMLAHAEETR | PXD018241 (Cell culture) | | | |
| EMLAHAEETRK | PXD018804, PXD018594 and PXD018241 (cell culture) | | | |
| ENDSKEGFFTYICGFIQQK | PXD018241 (Cell culture) | | | |
| ENSYTTTIKPVTYK | PXD018241 (Cell culture) | | | |
| EPCSSGTYEGNSPFHPLADNK | PXD018241 (Cell culture) | | | |
| EQIDGYVMHANYIFWR | PXD018241 (Cell culture) | | | |
| ESPFELEDFIPMDSTVK | PXD018241 (Cell culture) | | | |
| ESVQTFFK | PXD018241 (Cell culture) | | | |
| ETLYCIDGALLTK | PXD018241 (Cell culture) | | | |
| ETMSYLFQHANLDSCK | PXD018241 (Cell culture) | | | |
| ETMSYLFQHANLDSCKR | PXD018241 (Cell culture) | | | |
| EVGFVVPGLPGTILR | PXD018804, PXD018594 and PXD018241 (cell culture) | | | |
| FADDLNQLTGYK | PXD018241 (Cell culture) | | | |
| FADDLNQLTGYKKPASR | PXD018241 (Cell culture) | | | |
| FALTCFSTQFAFACPDGVK | PXD018241 (Cell culture) | | | |
| FAPSASAFFGMSR | PXD018241 (Cell culture) | | | |
| FASVYAWNR | PXD018804, PXD018594 and PXD018241 (cell culture) | | | |
| FCLEASFNYLK | PXD018241 (Cell culture) | | | |

| | | | | |
|---|---|---|---|---|
| FDEDDSEPVLK | PXD018241 (Cell culture) | | | |
| FDEDDSEPVLKG | PXD018241 (Cell culture) | | | |
| FDEDDSEPVLKGVK | PXD018241 (Cell culture) | | | |
| FDNPVLPFNDGVYFASTEK | PXD018804, PXD018594 and PXD018241 (Cell culture); PXD021328 (Clinical) | PXD021328 (Clinical) | Yes | |
| FDTFNGECPNFVFPLNSIIK | PXD018241 (Cell culture) | | | |
| FGDQELIR | PXD018804 (Cell culture) | | | |
| FGGPSDSTGSNQNGER | PXD018804 and PXD018241 (cell culture) | | | |
| FISTCACEIVGGQIVTCAK | PXD018241 (Cell culture) | | | |
| FKEGVEFLR | PXD018241 (Cell culture) | | | |
| FKESPFELEDFIPMDSTVK | PXD018241 (Cell culture) | | | |
| FKTEGLCVDIPGIPK | PXD018241 (Cell culture) | | | |
| FLALCADSIIIGGAK | PXD018241 (Cell culture) | | | |
| FLPFQQFGR | PXD018804, PXD018594 and PXD018241 (cell culture); PXD021328 (clinical) | PXD021328 (clinical) | | |
| FNFNGLTGTGVLTESNK | PXD018241 (Cell culture) | | | |
| FNGIGVTQNVLYENQK | PXD018804, PXD018594 and PXD018241 (cell culture); PXD021328 (clinical) | | | |
| FNGLTGTGVLTESNK | PXD018241 (Cell culture) | | | |
| FNGLTVLPPLLTD | PXD018241 (Cell culture) | | | |
| FNPPALQDAYYR | PXD018241 (Cell culture) | | | |
| FPNITNLCPFGEVFNATR | PXD018241 (Cell culture) | PXD019423 (Clinical) | | |
| FQEKDEDDNLIDSYFVVK | PXD018241 (Cell culture) | | | |
| FQPTNGVGYQPYR | PXD018241 (Cell culture) | | | |
| FQTLLALHR | PXD018804, PXD018594 and PXD018241 (cell culture) | | | Yes |
| FTALTQHGK | PXD018241 (Cell culture) | | | |
| FTALTQHGKEDLK | PXD018241 (Cell culture) | PXD021328 (Clinical) | | |
| FTALTQHGKEDLKFPR | PXD018241 (Cell culture) | | | |
| FTTTLNDFNLVAMK | PXD018241 (Cell culture) | | | |
| FVLALLSDLQDLK | PXD018241 (Cell culture) | | | |
| FVSLAIDAYPLTK | PXD018241 (Cell culture) | | | |
| FYDAQPCSDK | PXD018241 (Cell culture) | | | |
| GAGGHSYGADLK | PXD018241 (Cell culture) | | | |
| GANKDGIIWVATEGALNTPK | PXD018241 (Cell culture) | | | |
| GAWNIGEQK | PXD018241 (Cell culture) | | | |

| | | | | |
|---|---|---|---|---|
| GCCSCGSCCKFDEDDSEPVLK | PXD018241 (Cell culture) | | | |
| GDAALALLLLDR | PXD018241 (Cell culture) | | | |
| GDYGDAVVYR | PXD018241 (Cell culture) | | | |
| GEDIQLLK | PXD018241 (Cell culture) | | | |
| GFGDSVEEVLSEAR | PXD018241 (Cell culture) | | | |
| GFQPTNGVGYQPYR | PXD018241 (Cell culture); PXD021328 (Clinical) | PXD021328 (Clinical) | | |
| GFYAEGSR | PXD018804, PXD018594 and PXD018241 (cell culture); PXD021328 (clinical) | | | Yes |
| GFYAEGSRG | PXD018804 (Cell culture) | | | |
| GFYAEGSRGG | PXD018804 (Cell culture) | | | |
| GFYAEGSRGGSQA | PXD018804 (Cell culture) | | | |
| GFYAEGSRGGSQAS | PXD018804 (Cell culture) | | | |
| GFYAEGSRGGSQASSR | PXD018804, PXD018594 and PXD018241 (cell culture) | | | Yes |
| GGDAALALLLLDR | PXD018241 (Cell culture); PXD021328 (Clinical) | PXD021328 (Clinical) | | |
| GGDAALALLLLDRLNQLESK | PXD018241 (Cell culture) | | | |
| GGDGKMKDLSPR | PXD018804, PXD018594 and PXD018241 (cell culture) | | | |
| GGSYTNDKACPLIAAVITR | PXD018241 (Cell culture) | | | |
| GHFDGQQGEVPVSIINNTVYTK | PXD018241 (Cell culture) | | | |
| GIGVTQNVLYENQK | PXD018241 (Cell culture) | | | |
| GIIWVATEGALNTPK | PXD018241 (Cell culture) | | | |
| GIYQTSNFR | PXD018804, PXD018594 and PXD018241 (cell culture) | | | |
| GLPNNTASWFTALTQHGK | PXD018241 (Cell culture) | | | |
| GLPWNVVR | PXD018241 (Cell culture) | | | |
| GLTGTGVLTESNK | PXD018241 (Cell culture) | | | |
| GMVLGSLAATVR | PXD018804 and PXD018241 (cell culture) | | | |
| GNGGDAALALLLLDR | PXD018241 (Cell culture) | | | |
| GPEQTQGNFGDQELIR | PXD018804, PXD018594 and PXD018241 (cell culture); PXD021328 (clinical) | PXD021328 (Clinical) | | Yes |
| GPHEFCSQHTMLVK | PXD018241 (Cell culture) | | | |
| GPITDVFYK | PXD018241 (Cell culture) | | | |
| GPITDVFYKENSYTTTIKPVTYK | PXD018241 (Cell culture) | | | |
| GQGVPINTNSSPDDQIGY | PXD021328 (Clinical) | PXD021328 (Clinical) | | |
| GQGVPINTNSSPDDQIGYY | PXD021328 (Clinical) | PXD021328 (Clinical) | | |

| | | | | |
|---|---|---|---|---|
| GQGVPINTNSSPDDQIGYYR | PXD018804, PXD018594 and PXD018241 (cell culture); PXD021328 (clinical) | PXD021328 (clinical) | | |
| GQGVPINTNSSPDDQIGYYRR | PXD018804, PXD018594 and PXD018241 (cell culture); PXD019423 (clinical) | PXD019423 (clinical) | | |
| GQGVPINTNSSPDDQIGYYRRATR | PXD018804 (Cell culture) | | | |
| GQQQQGQTVTK | PXD018241 (Cell culture) | | | |
| GQQQQGQTVTKK | PXD018241 (Cell culture) | | | |
| GQQQQGQTVTKKSAAEASK | PXD018804, PXD018594 and PXD018241 (cell culture) | | | |
| GQQQQGQTVTKKSAAEASKKPR | PXD018804, PXD018594 and PXD018241 (cell culture) | | | |
| GSLPINVIVFDGK | PXD018241 (Cell culture) | | | |
| GTGPEAGLPYGANK | PXD018241 (Cell culture) | | | |
| GTLEPEYFNSVCR | PXD018241 (Cell culture) | | | |
| GTTVLLKEPCSSGTYEGNSPFHPLADNK | PXD018241 (Cell culture) | | | |
| GVAPGTAVLR | PXD018241 (Cell culture) | | | |
| GVEAVMYMGTLSYEQFK | PXD018241 (Cell culture) | PXD020493 (Clinical) | | |
| GVEAVMYMGTLSYEQFKK | PXD018241 (Cell culture) | | | |
| GVITHDVSSAINRPQIGVVR | PXD018241 (Cell culture) | | | |
| GVLPQLEQPYVFIK | PXD018241 (Cell culture) | | | |
| GVLPQLEQPYVFIKR | PXD018241 (Cell culture) | | | |
| GVQIPCTCGK | PXD018241 (Cell culture) | PXD021328 (Clinical) | | |
| GVVFLHVTYVPAQEK | PXD018241 (Cell culture) | | | |
| GVYFASTEK | PXD018241 (Cell culture) | | | |
| GVYYPDK | Not Detected | | Yes | |
| GVYYPDKVFR | PXD018804, PXD018594 and PXD018241 (cell culture) | PXD021328 (clinical) | | |
| GWIFGTTLDSK | PXD018804, PXD018594 and PXD018241 (cell culture); PXD021328 (clinical) | PXD021328 (Clinical) | Yes | Yes |
| GYHLMSFPQSAPH | PXD018241 (Cell culture) | | | |
| GYHLMSFPQSAPHGVVFLHVTYVPAQEK | PXD018241 (Cell culture) | | | |
| HADFDTWFSQR | PXD018241 (Cell culture) | | | |
| HCLHVVGPNVNK | PXD018241 (Cell culture) | | | |
| HCLHVVGPNVNKGEDIQLLK | PXD018241 (Cell culture) | | | |
| HFDEGNCDTLK | PXD018241 (Cell culture) | | | |
| HGGGVAGALNK | PXD018241 (Cell culture) | | | |

| | | | | |
|---|---|---|---|---|
| HGTFTCASEYTGNYQCGHYK | PXD018241 (Cell culture) | | | |
| HIDAYKTFPPTEPK | PXD018804, PXD018594 and PXD018241 (cell culture) | | | |
| HSLSHFVNLDNLR | PXD018241 (Cell culture) | | | |
| HTDFSSEIIGYK | PXD018804 and PXD018241 (cell culture) | | | |
| HTFSNYQHEETIYNLLK | PXD018241 (Cell culture) | | | |
| HTPINLVR | PXD018804, PXD018594 and PXD018241 (cell culture) | | Yes | Yes |
| HVICTSEDMLNPNYEDLLIR | PXD018241 (Cell culture) | | | |
| HVICTSEDMLNPNYEDLLIRK | PXD018241 (Cell culture) | | | |
| HWPQIAQF | PXD021328 (Clinical) | | | |
| HWPQIAQFAPSASAF | PXD018241 (Cell culture); PXD021328 (Clinical) | | | |
| HWPQIAQFAPSASAFF | PXD018241 (Cell culture); PXD021328 (Clinical) | | | |
| HWPQIAQFAPSASAFFGM | PXD018241 (Cell culture) | | | |
| HWPQIAQFAPSASAFFGMSR | PXD018804, PXD018594 and PXD018241 (cell culture); PXD021328 (clinical) | | | |
| HYVYIGDPAQLPAPR | PXD018241 (Cell culture) | | | |
| IADYNYK | Not Detected | | Yes | |
| IADYNYKLPDDFTGCVIAWNSNNLDSK | PXD018241 (Cell culture) | | | |
| IAEIPKEEVKPFITESKPSVEQR | PXD018241 (Cell culture) | | | |
| IAGHHLGR | PXD018804 and PXD018594 (Cell culture); PXD021328 (Clinical) | | | Yes |
| IAQFAPSASAFFGMSR | PXD018241 (Cell culture) | | | |
| IFTIGTVTLK | PXD018804, PXD018594 and PXD018241 (cell culture) | | | |
| IFVDGVPFVVSTGYHFR | PXD018241 (Cell culture) | | | |
| IGMEVTPSGTWLTY | PXD021328 (Clinical) | | | |
| IGMEVTPSGTWLTYTGAIK | PXD018804, PXD018594 and PXD018241 (cell culture); PXD021328 and PXD019423 (clinical) | PXD021328 and PXD019423(clinical) | | Yes |
| IGMEVTPSGTWLTYTGAIKLDDKDPNFK | PXD018241 (Cell culture) | | | |
| IGMEVTPSGTWLTYTGAIKLDDKDPNFKDQVILLNK | PXD018241 (Cell culture) | | | |
| IGNYKLNTDHSSSSDNIALLVQ | PXD018804, PXD018594 and PXD018241 (cell culture) | | | |
| ILGAGCFVDDIVK | PXD018241 (Cell culture) | | | |
| IMASLVLAR | PXD018241 (Cell culture) | | | |

| | | | | |
|---|---|---|---|---|
| IMTWLDMVDTSLSGFK | PXD018241 (Cell culture) | | | |
| IQDSLSSTASALGK | PXD018804, PXD018594 and PXD018241 (cell culture) | | | |
| IQEGVVDYGAR | PXD018804, PXD018594 and PXD018241 (cell culture) | | | |
| IRGGDGKMKDLSPR | PXD018804 and PXD018594 (cell culture) | | | |
| ISEMHPALR | PXD018804, PXD018594 and PXD018241 (cell culture) | | | |
| ISNCVADYSVLYNSASFSTFK | PXD018241 (Cell culture) | | | |
| ITEEVGHTDLMAAYVDNSSLTIK | PXD018241 (Cell culture) | | | |
| ITEHSWNADLYK | PXD018241 (Cell culture) | | | |
| ITFGGPSDSTGSNQNGER | PXD018804, PXD018594 and PXD018241 (cell culture); PXD021328 (clinical) | PXD021328 (clinical) | | |
| ITFGGPSDSTGSNQNGERSGAR | PXD018804, PXD018594 and PXD018241 (cell culture) | | | |
| ITGLYPTLNISDEFSSNVANYQK | PXD018241 (Cell culture) | | | |
| IVDEPEEHVQIH | PXD018241 (Cell culture) | | | |
| IVDEPEEHVQIHTI | PXD018241 (Cell culture) | | | |
| IVDEPEEHVQIHTID | PXD018241 (Cell culture) | PXD021328 (Clinical) | | |
| IVDEPEEHVQIHTIDG | PXD018241 (Cell culture) | | | |
| IVDEPEEHVQIHTIDGSSGVVNPVMEPIYDEPTTTTSVPL | PXD018241 (Cell culture) | | | |
| IVITSGDGTTSPISEHDYQIGGYTEK | PXD018241 (Cell culture) | | | |
| IVLQLPQGTTLPK | PXD018241 (Cell culture) | | | |
| IVQLSEISMDNSPNLAWPLIVTALR | PXD018241 (Cell culture) | | | |
| IVYTACSHAAVDALCEK | PXD018241 (Cell culture) | | | |
| IWVATEGALNTPK | PXD018241 (Cell culture) | | | |
| IYCPACHNSEVGPEHSLAEYHNESGLK | PXD018241 (Cell culture) | PXD019423 (Clinical) | | |
| IYSKHTPINLVR | PXD018804 (Cell culture) | | | |
| KADETQALPQ | PXD018804 (Cell culture) | | | |
| KADETQALPQR | PXD018804, PXD018594 and PXD018241 (cell culture); PXD021328 (clinical) | PXD021328 (clinical) | | |
| KADETQALPQRQ | PXD018804 and PXD018594 (cell culture) | | | |
| KADETQALPQRQK | PXD018804 and PXD018594 (cell culture) | | | |
| KADETQALPQRQKK | PXD018804 and PXD018594 (cell culture) | | | |
| KADETQALPQRQR | Not Detected | | | Yes |

| | | | | |
|---|---|---|---|---|
| KADETQALPQRQRQK | Not Detected | | | Yes |
| KDAPYIVGDVVQEGVLTAVVIPTKK | PXD018241 (Cell culture) | | | |
| KDGIIWVATEGALNTPK | PXD018241 (Cell culture) | | | |
| KDKKKKADETQALPQR | PXD018594 (Cell culture) | | | |
| KDNSYFTEQPIDLVPNQPYPNASFDNFK | PXD018241 (Cell culture) | | | |
| KFDTFNGECPNFVFPLNSIIK | PXD018241 (Cell culture) | | | |
| KKADETQALPQR | PXD018804, PXD018594 and PXD018241 (cell culture); PXD021328 (clinical) | PXD021328 (Clinical) | | Yes |
| KKADETQALPQRQK | PXD018804 and PXD018594 (cell culture) | | | |
| KKKADETQALPQR | PXD018804 and PXD018594 (cell culture) | | | |
| KLDNDALNNIINNAR | PXD018241 (Cell culture) | | | |
| KLMPVCVETK | PXD018241 (Cell culture) | | | |
| KPTETICAPLTVFFDGR | PXD018241 (Cell culture) | | | |
| KQQTVTLLPAADLDD | PXD018241 (Cell culture) | | | |
| KQQTVTLLPAADLDDFSK | PXD018804, PXD018594 and PXD018241 (cell culture); PXD021328 (clinical) | PXD021328 (clinical) | | |
| KQQTVTLLPAADLDDFSKQLQQSMSSADSTQA | PXD018241 (Cell culture) | | | |
| KSAAEASK | PXD018241 (Cell culture) | | | |
| KSAPLIELCVDEAGSK | PXD018241 (Cell culture) | | | |
| KSNHNFLVQAGNVQLR | PXD018241 (Cell culture) | | | |
| KSNLKPFER | PXD018241 (Cell culture); PXD021328 (Clinical) | PXD021328 (Clinical) | | |
| KTLNSLEDK | PXD018804 and PXD018241 (Cell culture) | | | |
| KTLNSLEDKAFQLTPIAVQMTK | PXD018241 (Cell culture) | | | |
| KVDGVVQQLPETYFTQSR | PXD018241 (Cell culture) | | | |
| KVKPTVVVNAANVYLK | PXD018241 (Cell culture) | | | |
| KVPTDNYITTYPGQGLNGYTVEEAK | PXD018241 (Cell culture) | | | |
| LATTEELPDEFVVVTVK | PXD018804, PXD018594 and PXD018241 (cell culture) | | | |
| LCEEMLDNR | PXD018241 (Cell culture) | | | |
| LDDKDPNFK | PXD018241 (Cell culture); PXD021328 (Clinical) | PXD021328 (Clinical) | | |
| LDDKDPNFKDQVILLNK | PXD018804, PXD018594 and PXD018241 (cell culture); PXD021328 (clinical) | PXD021328 (clinical) | | |
| LDDKDPNFKDQVILLNKHIDAYK | PXD018804, PXD018594 and PXD018241 (cell culture) | | | |
| LDDKDPQFKDNVILLNK | PXD018241 (Cell culture) | | | |

| | | | | |
|---|---|---|---|---|
| LDDKDPQFKDNVILLNKHIDAYK | PXD018804 (Cell culture) | | | |
| LDGVVCTEIDPK | PXD018241 (Cell culture) | | | |
| LDKVEAEVQIDR | PXD018804, PXD018594 and PXD018241 (cell culture) | | | |
| LDNDALNNIINNAR | PXD018241 (Cell culture) | | | |
| LGASQRVAGDSGFAAYSR | PXD018804 (Cell culture) | | | Yes |
| LGSPLSLNMAR | PXD018804 and PXD018241 (Cell culture) | | | |
| LGTGPEAGLPYGANK | PXD018594 and PXD018241 (Cell culture); PXD021328 (Clinical) | PXD021328 (Clinical) | | |
| LHNWNCVNCDTFCAGSTFISDEVAR | PXD018241 (Cell culture) | | | |
| LHVTYVPAQEK | PXD018241 (Cell culture) | | | |
| LIANQFNSAIGK | PXD018804, PXD018594 and PXD018241 (cell culture); PXD021328 (clinical) | PXD021328 (clinical) | | |
| LIANQFNSAIGKIQDSLSSTASALGK | PXD018241 (Cell culture) | | | |
| LKPVLDWLEEK | PXD018241 (Cell culture) | | | |
| LKTLVATAEAELAK | PXD018241 (Cell culture) | | | |
| LKVDTANPK | PXD018804 and PXD018594 (cell culture) | | | |
| LLDRLNQLESK | PXD021328 (Clinical) | | | |
| LLHKPIVW | PXD018241 (Cell culture) | | | |
| LLHKPIVWHVNNATNK | PXD018241 (Cell culture) | | | |
| LLPAADLDDFSK | PXD018241 (Cell culture) | | | |
| LMPVCVETK | PXD018804 and PXD018241 (cell culture) | | | |
| LMVVIPDYNTYK | PXD018241 (Cell culture) | | | |
| LNDLCFTNVYADSFVIR | PXD018241 (Cell culture) | | | |
| LNDLCFTNVYADSFVIRGDEVR | PXD018241 (Cell culture) | | | |
| LNQLESKMSGK | PXD018804, PXD018594 and PXD018241 (cell culture) | | | |
| LNTDHSSSSDNIALLVQ | PXD018804 and PXD018594 (cell culture); PXD021328 (Clinical) | PXD021328 and PXD019423 (Clinical) | | |
| LPDDFTGCVIAWNSNNLDSK | PXD018241 (Cell culture) | | Yes | |
| LQAGNATEVPANSTVLSFCAFAVDAAK | PXD018241 (Cell culture) | | | |
| LQDVVNQNAQALN | PXD018241 (Cell culture) | | | |
| LQDVVNQNAQALNTLVK | PXD018804, PXD018594 and PXD018241 (cell culture); PXD021328 (clinical) | | | |
| LQFTSLEIPR | PXD018241 (Cell culture) | | | |

| | | | | |
|---|---|---|---|---|
| LQNNELSPVALR | PXD018241 (Cell culture) | | | |
| LQSLENVAFNVVNK | PXD018241 (Cell culture) | | | |
| LQSLQTYVTQQLIR | PXD018804, PXD018594 and PXD018241 (cell culture); PXD021328 (clinical) | | | Yes |
| LRSDVLLPLTQY | PXD018241 (Cell culture) | | | |
| LSHQSDIEVTGDSCNNYMLTYNK | PXD018241 (Cell culture) | | | |
| LSYGIATVR | PXD018241 (Cell culture) | | | |
| LTDNVYIK | PXD018241 (Cell culture) | | | |
| LTPCGTGTSTDVVYR | PXD018241 (Cell culture) | | | |
| LTQHGKEDLK | PXD018241 (Cell culture); PXD021328 (Clinical) | PXD021328 (Clinical) | | |
| LVDPQIQLAVTR | PXD018804, PXD018594 and PXD018241 (cell culture) | | | |
| LVSSFLEMK | PXD018241 (Cell culture) | | | |
| MADQAMTQMYK | PXD018241 (Cell culture) | | | |
| MADSNGTITVEELKK | PXD018241 (Cell culture) | | | |
| MAGNGGDAALALLLLDR | PXD018804, PXD018594 and PXD018241 (cell culture); PXD021328, PXD019423 and PXD020493 (clinical) | PXD021328, PXD019423 and PXD020493 (clinical) | | |
| MAGNGGDAALALLLLDRLN | PXD018241 (Cell culture) | | | |
| MAGNGGDAALALLLLDRLNQLESK | PXD018804, PXD018594 and PXD018241 (cell culture); PXD021328 and PXD019423 (clinical) | PXD021328 and PXD019423 (clinical) | | |
| MAGNGGDAALALLLLDRLNQLESKMSGK | PXD018804, PXD018594 and PXD018241 (cell culture) | | | |
| MASGGGETALALLLLDR | PXD018241 (Cell culture) | | | |
| MASGGGETALALLLLDRLNQLESK | PXD018804 (Cell culture) | | | |
| MENAVGRDQNNVGPK | PXD018804 and PXD018241 (Cell culture) | | | |
| MESLVPGFNEK | PXD018241 (Cell culture) | | | |
| MFDAYVNTFSSTFNVPMEK | PXD018241 (Cell culture) | | | |
| MFVFLVLLPLVSSQCVNLTTR | Not Detected | | Yes | |
| MKDLSPRWYFYYLGTGPEAGLPYGANK | PXD018241 (Cell culture) | | | |
| MNYQVNGYPNMFITR | PXD018241 (Cell culture) | | | |
| MSECVLGQSK | PXD018804, PXD018594 and PXD018241 (cell culture) | | | |
| MSECVLGQSKR | PXD018241 (Cell culture) | | | |
| MSGKGQQQQGQTVTK | PXD018241 (Cell culture) | | | |
| MSGKGQQQQGQTVTKK | PXD018241 (Cell culture) | | | |

| | | | | |
|---|---|---|---|---|
| NAAIVLQLPQGTTLPK | PXD018241 (Cell culture) | | | |
| NADIVEEAK | PXD018241 (Cell culture) | | | |
| NADIVEEAKK | PXD018804 and PXD018241 (cell culture) | | | |
| NAPRITFGGPSDSTGSNQNGER | PXD018241 (Cell culture) | | | |
| NDGVYFASTEK | PXD018241 (Cell culture) | | | |
| NFNGLTGTGVLTESNKK | PXD018241 (Cell culture) | | | |
| NFTTAPAICHDGK | PXD018241 (Cell culture) | | | |
| NFTTAPAICHEGK | PXD018241 (Cell culture) | | | |
| NGSIHLYFDK | PXD018241 (Cell culture) | | | |
| NGVLITEGSVK | PXD018241 (Cell culture) | | | |
| NHTSPDVDLGDISGINASVVNIQK | PXD018241 (Cell culture) | | | |
| NIDGYFK | Not Detected | PXD021328 (Clinical) | Yes | |
| NIKPVPEVK | PXD018241 (Cell culture) | | | |
| NLNESLIDLQELGK | PXD018241 (Cell culture) | | | |
| NLNSSRVPDLLV | PXD018241 (Cell culture) | | | |
| NLYDKLVSSFLEMK | PXD018241 (Cell culture) | | | |
| NNELSPVALR | PXD018241 (Cell culture) | | | |
| NPANNAAIVLQLPQGT | PXD021328 (Clinical) | PXD021328 (Clinical) | | |
| NPANNAAIVLQLPQGTTLPK | PXD018804, PXD018594 and PXD018241 (cell culture); PXD021328 (clinical) | PXD021328 and PXD019423(clinical) | | Yes |
| NPANNAAIVLQLPQGTTLPKG | PXD018241 (Cell culture) | | | |
| NPANNAAIVLQLPQGTTLPKGFYAEGSR | PXD018241 (Cell culture) | | | |
| NPANNAAIVLQLPQGTTLPKGFYAEGSRG | PXD018241 (Cell culture) | | | |
| NPLLYDANYFL | PXD018241 (Cell culture) | PXD019423 (Clinical) | | |
| NPLLYDANYFLCWH | PXD018241 (Cell culture) | | | |
| NSIDAFKLNIK | PXD018241 (Cell culture) | | | |
| NSSPDDQIGYYR | PXD018241 (Cell culture); PXD021328 (Clinical) | PXD021328 (Clinical) | | |
| NSTPGSSR | PXD018241 (Cell culture) | | | |
| NSTPGSSRGTSPA | PXD018241 (Cell culture) | | | |
| NTASWFTALTQHGK | PXD018241 (Cell culture) | | | |
| NTASWFTALTQHGKEDLK | PXD018241 (Cell culture) | | | |
| NTNPIQLSSYSLFDMSK | PXD018241 (Cell culture) | | | |
| NTNSSPDDQIGYYR | PXD021328 (Clinical) | PXD021328 (Clinical) | | |

| | | |
|---|---|---|
| NTQEVFAQVK | PXD018241 (Cell culture) | |
| NTVCTVCGMWK | PXD018241 (Cell culture) | |
| NVATLQAENVTGLFK | PXD018241 (Cell culture) | |
| NVIPTITQMNLK | PXD018241 (Cell culture) | |
| NVSLDNVLSTFISAAR | PXD018241 (Cell culture) | |
| NVTQAFGR | PXD021328 (Clinical) | |
| NYFITDAQTGSSK | PXD018241 (Cell culture) | |
| NYVFTGYR | PXD018241 (Cell culture) | |
| PAADLDDFSK | PXD018241 (Cell culture); PXD021328 (Clinical) | PXD021328 (Clinical) |
| PANNAAIVLQLPQGTTLPK | PXD018241 (Cell culture) | |
| PDDQIGYYR | PXD018241 (Cell culture) | |
| PINTNSSPDDQIGYYRR | PXD018241 (Cell culture) | |
| PLLESELVIGAVILR | PXD018241 (Cell culture) | |
| PNFKDQVILLNK | PXD018241 (Cell culture) | |
| PNNTASWFTALTQHGK | PXD018241 (Cell culture) | |
| PPAYTNSFTR | PXD018241 (Cell culture) | |
| PQGLPNNTASWFTALTQHGK | PXD018241 (Cell culture) | |
| PQGLPNNTASWFTALTQHGKEDLK | PXD018241 (Cell culture) | |
| PQIAQFAPSASAFFGMSR | PXD018241 (Cell culture) | |
| PSASAFFGMSR | PXD018241 (Cell culture) | |
| PSFYVYSR | PXD018241 (Cell culture) | |
| PSGTWLTYTGAIK | PXD018241 (Cell culture) | |
| PSSKRFQPFQQFGR | PXD021328 (Clinical) | |
| PVETSNSFDVLK | PXD018241 (Cell culture) | |
| QASLNGVTLIGEAVK | PXD018241 (Cell culture) | |
| QEILGTVSWNLR | PXD018241 (Cell culture) | |
| QFDTYNLWNTFTR | PXD018241 (Cell culture) | |
| QGDDYVYLPYPDPSR | PXD018241 (Cell culture) | |
| QGEIKDATPSDFVR | PXD018804 and PXD018241 (cell culture) | |
| QGFVDSDVETK | PXD018241 (Cell culture) | |
| QGFVDSDVETKDVVECLK | PXD018241 (Cell culture) | |
| QGTDYKHWPQIAQFAPSASAFFGMSR | PXD018804, PXD018594 and PXD018241 (cell culture); PXD020493 (clinical) | |
| QHLKDGTCGLVEVEK | PXD018241 (Cell culture) | |

| | | | | |
|---|---|---|---|---|
| QIAPGQTGK | PXD018804 and PXD018241 (Cell culture) | | Yes | |
| QIVESCGNFK | PXD018241 (Cell culture) | | | |
| QKKQQTVTLLPAADLDDFSK | PXD018804, PXD018594 and PXD018241 (cell culture) | | | |
| QKRTATKAYNVTQAFGR | PXD018804 and PXD018594 (cell culture); PXD020493 (Clinical) | PXD020493 (Clinical) | | |
| QLPFFYYSDSPCESHGK | PXD018241 (Cell culture) | | | |
| QLPQGTTLPK | PXD021328 (Clinical) | | | |
| QLQQSMSSADSTQA | PXD018804, PXD018594 and PXD018241 (cell culture); PXD021328 (clinical) | PXD021328 (clinical) | Yes | |
| QLSSNFGAISSVLNDILSR | PXD018804, PXD018594 and PXD018241 (cell culture) | | | |
| QPTNGVGYQPYR | PXD018241 (Cell culture) | | | |
| QQQGQTVTKK | PXD018241 (Cell culture) | | | |
| QQTVTLLPAADLDDFSK | PXD018804, PXD018594 and PXD018241 (cell culture); PXD021328 (clinical) | PXD021328 (clinical) | | |
| QSYGFQPTNGVGYQPYR | PXD018241 (Cell culture) | | | |
| QVVNVVTTK | PXD018241 (Cell culture) | | | |
| QVVSDIDYVPLK | PXD018241 (Cell culture) | | | |
| QYGDCLGDIAAR | PXD018241 (Cell culture) | | | |
| QYNVTQAFGR | PXD018241 (Cell culture) | | | |
| RCPAEIVDTVSALVYDNK | PXD018241 (Cell culture) | | | |
| RFDNPVLPFNDGVYFASTEK | PXD018241 (Cell culture); PXD021328 (Clinical) | PXD021328 (Clinical) | | |
| RGPEQTQGNFGDQDLIR | PXD018241 (Cell culture) | | | |
| RGPEQTQGNFGDQELIR | PXD018804, PXD018594 and PXD018241 (cell culture); PXD021328, PXD019423 and PXD020493 (clinical) | PXD021328, PXD019423 and PXD020493 (clinical) | | Yes |
| RGPEQTQGNFGDQELIRQ | PXD018804 (Cell culture) | | | |
| RGQGVPINTNSSPDDQIGYYR | PXD018594 and PXD018241 (Cell culture) | PXD019423 (Clinical) | | |
| RITFGGPSDSTGSNQNGER | PXD018241 (Cell culture) | | | |
| RNPANNAAIVLQLPQGTTLPK | PXD018241 (Cell culture) | | | |
| RPINPTDQSSYIVDSVTVK | PXD018241 (Cell culture) | | | |
| RPQGLPNNTASW | PXD018241 (Cell culture); PXD021328 (Clinical) | | | |
| RPQGLPNNTASWF | PXD018241 (Cell culture); PXD021328 (Clinical) | | | |
| RPQGLPNNTASWFT | PXD021328 (Clinical) | | | |
| RPQGLPNNTASWFTAL | PXD018241 (Cell culture); PXD021328 (Clinical) | | | |

| | | | | |
|---|---|---|---|---|
| RPQGLPNNTASWFTALTQ H | PXD018241 (Cell culture) | | | |
| RPQGLPNNTASWFTALTQ HGK | PXD018804, PXD018594 and PXD018241 (cell culture); PXD021328 and PXD019423 (clinical) | | | |
| RPQGLPNNTASWFTALTQ HGKEDLK | PXD018241 (Cell culture); PXD021328 (Clinical) | PXD021328 (Clinical) | | |
| RPQGLPNNTASWFTALTQ HGKEDLKFPR | PXD018241 (Cell culture); PXD021328 and PXD019423 (Clinical) | PXD021328 and PXD019423 (Clinical) | | |
| RPQGLPNNTASWFTALTQ HGKEELR | PXD018241 (Cell culture) | | | |
| RSFIEDLLFNK | PXD018241 (Cell culture) | | | |
| RTATKAYNVTQAFGR | PXD018804 (Cell culture); PXD020493 (Clinical) | PXD020493 (Clinical) | | Yes |
| RWQLALSK | PXD018241 (Cell culture) | | | |
| SADAQSFLNR | PXD018241 (Cell culture) | | | |
| SAFYILPSIISNEK | PXD018241 (Cell culture) | | | |
| SAGFPFNK | PXD018241 (Cell culture) | | | |
| SALEPLVDLPIGINITR | PXD018241 (Cell culture) | | | |
| SAPLIELCVDEAGSK | PXD018241 (Cell culture) | | | |
| SAYENFNQHEVLLAPLLSAG IFGADPIHSLR | PXD018241 (Cell culture) | | | |
| SDVLLPLTQYNR | PXD018241 (Cell culture) | | | |
| SEAGVCVSTSGR | PXD018804 and PXD018241 (cell culture) | | | |
| SEDAQGMDNLACEDLKPVS EEVVENPTIQK | PXD018241 (Cell culture) | | | |
| SFIEDLLFNK | PXD018804, PXD018594 and PXD018241 (cell culture); PXD021328 (clinical) | | | |
| SFIEDLLFNKVTLADAGFIK | PXD018804, PXD018594 and PXD018241 (cell culture) | | | |
| SFNPETNILLNVPLHGTILTR PLLESELVIGAVILR | PXD021328 (Clinical) | PXD021328 (Clinical) | | |
| SFYVYANGGK | PXD018241 (Cell culture) | | | |
| SGDGTTSPISEHDYQIGGYT EK | PXD018241 (Cell culture) | | | |
| SGETLGVLVPHVGEIPVAYR | PXD018241 (Cell culture) | | | |
| SGETLGVLVPHVGEIPVAYR K | PXD018241 (Cell culture) | | | |
| SHKPPISFPLCANGQVFGLY K | PXD018241 (Cell culture) | | | |
| SHNIALIWNVK | PXD018241 (Cell culture) | | | |
| SILSPLYAFASEAAR | PXD018241 (Cell culture) | | | |
| SIVITSGDGTTSPISEHDYQI GGYTEK | PXD018241 (Cell culture) | | | |

| | | | |
|---|---|---|---|
| SLENVAFNVVNK | PXD018241 (Cell culture) | | |
| SLKVPATVSVSSPDAVTAYNGYLTSSSK | PXD018241 (Cell culture) | | |
| SLTENKYSQLDEEQPMEID | PXD018241 (Cell culture) | | |
| SMWSFNPETNILLNVPLHGTILTR | PXD018241 (Cell culture) | | |
| SNGTITVEELKK | PXD018241 (Cell culture) | | |
| SNHNFLVQAGNVQLR | PXD018241 (Cell culture) | | |
| SNLKPFER | PXD018804, PXD018594 and PXD018241 (cell culture); PXD021328 (clinical) | PXD021328 (clinical) | |
| SPDDQIGYYR | PXD018804 (Cell culture) | PXD021328 (clinical) | |
| SQDLSVVSK | PXD018241 (Cell culture) | | |
| SREETGLLMPLK | PXD018241 (Cell culture) | | |
| SREETGLLMPLKAPK | PXD018241 (Cell culture) | | |
| SSEYKGPITDVFYK | PXD018241 (Cell culture) | | |
| SSEYKGPITDVFYKENSYTTTIKPVTYK | PXD018241 (Cell culture) | | |
| SSGTYEGNSPFHPLADNK | PXD018241 (Cell culture) | | |
| SSPDDQIGYYR | PXD018804, PXD018594 and PXD018241 (cell culture); PXD021328 (clinical) | PXD021328 (clinical) | |
| SSPDDQIGYYRR | PXD018804 (Cell culture) | | |
| SSVLHSTQDLFLPF | PXD018241 (Cell culture) | | |
| STDTGVEHVTFFIYNK | PXD018241 (Cell culture) | PXD019423 (Clinical) | |
| SVLYNSASFSTFK | PXD018241 (Cell culture) | | |
| SVLYYQNNVFMSEAK | PXD018241 (Cell culture) | | |
| SVNITFELDER | PXD018241 (Cell culture) | PXD021328 (Clinical) | |
| SVTSSIVITSGDGTTSPISEHDYQIGGYTEK | PXD018241 (Cell culture) | | |
| SVYPVASPNECNQMCLSTLMK | PXD018241 (Cell culture) | | |
| SVYYTSNPTTFHLDGEVITFDNLK | PXD018241 (Cell culture) | | |
| SWFTALTQHGKEDLK | PXD018241 (Cell culture) | | |
| SWMESEFR | PXD018804 and PXD018241 (Cell culture); PXD021328 (Clinical) | PXD021328 (Clinical) | Yes |
| SYELQTPFEIK | PXD018241 (Cell culture) | | |
| SYLTPGDSSSGW | PXD018241 (Cell culture) | | |
| SYLTPGDSSSGWTAGAAAYYVGYLQPR | PXD018804 and PXD018241 (cell culture) | | |
| SYYKLGASQR | PXD018241 (Cell culture) | | |

| | | | | |
|---|---|---|---|---|
| TAGAAAYYVGYLQPR | PXD018241 (Cell culture) | PXD021328 (Clinical) | | |
| TALTQHGKEDLK | PXD018241 (Cell culture); PXD021328 and PXD019423 (Clinical) | PXD021328 (Clinical) | | |
| TALTQHGKEDLKFPR | PXD018241 (Cell culture) | PXD021328 (Clinical) | | |
| TASWFTALTQHGK | PXD018241 (Cell culture) | | | |
| TATKAYNVTQAFGR | PXD018804, PXD018594 and PXD018241 (cell culture); PXD020493 (clinical) | PXD020493 (clinical) | | Yes |
| TATKQYNVTQAFGR | PXD018241 (Cell culture) | | | |
| TCGQQQTTLK | PXD018241 (Cell culture) | | | |
| TDGTLMIER | PXD018241 (Cell culture) | | | |
| TFPPTEPK | PXD018594 and PXD018241 (Cell culture) | | | |
| TFPPTEPKK | PXD018241 (Cell culture) | | | |
| TFPPTEPKKDK | PXD018241 (Cell culture) | | | |
| TFPPTEPKKDKK | PXD018804, PXD018594 and PXD018241 (cell culture) | | | |
| TFPPTEPKKDKKK | PXD018804 (Cell culture) | | | |
| TFYVLPNDDTLR | PXD018241 (Cell culture) | | | |
| THVQLSLPVLQVR | PXD018241 (Cell culture) | | | |
| TIAFGGCVFSYVGCHNK | PXD018241 (Cell culture) | | | |
| TIGPDMFLGTCR | PXD018241 (Cell culture) | | | |
| TILGSALLEDEFTPFDVVR | PXD018804, PXD018594 and PXD018241 (cell culture) | | | |
| TLADAGFIK | PXD018241 (Cell culture) | | | |
| TLATHGLAAVNSVPWDTIANYAK | PXD018241 (Cell culture) | | | |
| TLETAQNSVR | PXD018241 (Cell culture) | | | |
| TLLPAADLDDFSK | PXD021328 (Clinical) | PXD021328 and PXD019423 (Clinical) | | |
| TLNSLEDK | PXD018241 (Cell culture) | | | |
| TLNSLEDKAFQLTPIAVQMTK | PXD018804 and PXD018241 (Cell culture) | | | |
| TLNSLEDKAFQLTPIAVQMTKLATTEELPDEFVVVTVK | PXD018241 (Cell culture) | | | |
| TLSYYKLGASQR | PXD018241 (Cell culture) | | | |
| TLSYYKLGASQRVAGDSGFAAYSR | PXD018241 (Cell culture) | PXD021328 (Clinical) | | |
| TLVATAEAELAK | PXD018594 and PXD018241 (Cell culture) | | | |
| TNSSPDDQIGYYR | PXD018804 and PXD018241 (cell culture) | | | |
| TNSSPDDQIGYYRR | PXD018804 (Cell culture) | | | |

| | | | | |
|---|---|---|---|---|
| TNVYADSFVIR | PXD018241 (Cell culture) | | | |
| TNVYLAVFDK | PXD018241 (Cell culture) | | | |
| TNVYLAVFDKNLYDK | PXD018241 (Cell culture) | | | |
| TPEEHFIETISLAGSYK | PXD018241 (Cell culture) | | | |
| TPPIKDFGGFNFSQILPDPSK PSK | PXD018241 (Cell culture) | | | |
| TQGNFGDQELIR | PXD018804 (Cell culture) | | | |
| TQLPPAYTNSFTR | PXD018804, PXD018594 and PXD018241 (cell culture); PXD021328 (clinical) | PXD021328 (clinical) | Yes | |
| TQSLLIVNNATNVVIK | PXD018241 (Cell culture) | | Yes | |
| TTEVVGDIILKPANNSLK | PXD018241 (Cell culture) | | | |
| TTLPVNVAFELWAK | PXD018241 (Cell culture) | | | |
| TTNGDFLHFLPR | PXD018804, PXD018594 and PXD018241 (cell culture) | | | |
| TVAGVSICSTMTNR | PXD018241 (Cell culture) | | | |
| TVGELGDVR | PXD018241 (Cell culture) | | | |
| TVYSDVENPHLMGWDYPK | PXD018241 (Cell culture) | | | |
| VAGDSGFAAY | Not Detected | | | Yes |
| VAGDSGFAAYSR | PXD018804, PXD018594 and PXD018241 (cell culture); PXD021328 (clinical) | PXD021328 (clinical) | | |
| VAGDSGFAAYSRYR | PXD018804 and PXD018594 (Cell culture) | PXD019423 (Clinical) | | Yes |
| VATEGALNTPK | PXD018241 (Cell culture); PXD021328 (Clinical) | | | |
| VCEFQFCNDPFLGVYYHK | PXD018241 (Cell culture) | | Yes | |
| VCGVSAAR | PXD018241 (Cell culture) | | | |
| VDGQVDLFR | PXD018241 (Cell culture) | | | |
| VDGVDVELFENK | PXD018241 (Cell culture) | | | |
| VDGVVQQLPETYFTQSR | PXD018241 (Cell culture) | PXD020493 (Clinical) | | |
| VEAEVQIDR | PXD018241 (Cell culture) | | | |
| VEAFEYYHTTDPSFLGR | PXD018804, PXD018594 and PXD018241 (cell culture) | | | |
| VECTTIVNGVR | PXD018241 (Cell culture) | | | |
| VFSAVGNICYTPSK | PXD018241 (Cell culture) | | | |
| VGGNYNYLYR | PXD018804, PXD018594 and PXD018241 (cell culture); PXD021328 (clinical) | PXD021328 (clinical) | Yes | |
| VGGSCLLSGHNLAK | PXD018241 (Cell culture) | | | |
| VGGSCVLSGHNLAK | PXD018241 (Cell culture) | | | |
| VIGHSMQNCVLK | PXD018241 (Cell culture) | | | |
| VIHFGAGSDK | PXD018241 (Cell culture) | | | |

| | | | | |
|---|---|---|---|---|
| VKPTVVVNAANVYLK | PXD018241 (Cell culture) | | | |
| VNINIVGDFK | PXD018241 (Cell culture) | | | |
| VPATVSVSSPDAVTAYNGYLTSSSK | PXD018241 (Cell culture) | | | |
| VPTDNYITTYPGQGLNGYTVEEAK | PXD018241 (Cell culture) | | | |
| VQIGEYTFEK | PXD018241 (Cell culture) | | | |
| VQPTESIVR | PXD018804, PXD018594 and PXD018241 (cell culture) | PXD021328 (clinical) | Yes | |
| VTFFPDLNGDVVAIDYK | PXD018804 and PXD018241 (cell culture) | | | |
| VTFGDDTVIEVQGYK | PXD018241 (Cell culture) | | | |
| VTLADAGFIK | PXD018804 and PXD018594 (cell culture); PXD021328 (Clinical) | PXD021328 (Clinical) | | |
| VTSAMQTMLFTMLR | PXD018241 (Cell culture) | | | |
| VVNQNAQALNTLVK | PXD018241 (Cell culture) | | | |
| VVSTTTNIVTR | PXD018241 (Cell culture) | | | |
| VVTTFDSEYCR | PXD018241 (Cell culture) | | | |
| VVVLSFELLHAPATVCGPK | PXD018241 (Cell culture) | | | |
| VYANLGER | PXD018241 (Cell culture) | | | |
| VYPIILRL | PXD018804 (Cell culture) | | | |
| VYSSANNCTFEYVSQPFLMDLEGK | PXD018241 (Cell culture) | PXD019423 (Clinical) | Yes | |
| VYSTGSNVFQTR | PXD018804, PXD018594 and PXD018241 (cell culture); PXD021328 (clinical) | PXD021328 (clinical) | Yes | |
| WKYPQVNGLTSIK | PXD018241 (Cell culture) | | | |
| WVATEGALNTPK | PXD018241 (Cell culture) | | | |
| WVLNNDYYR | PXD018241 (Cell culture) | | | |
| WYFYYLGTGPEAGLPYGAN | PXD018804 (Cell culture) | | | |
| WYFYYLGTGPEAGLPYGANK | PXD018804, PXD018594 and PXD018241 (cell culture); PXD021328 (clinical) | PXD021328 (Clinical) | | Yes |
| WYFYYLGTGPEAGLPYGANKDGIIWVATEGALNTPK | PXD018804, PXD018594 and PXD018241 (cell culture) | | | Yes |
| WYFYYLGTGPEAGLPYGANKDGIIWVATEGALNTPKDHIGTR | PXD018241 (Cell culture) | | | |
| WYFYYLGTGPEASLPYGANK | PXD018241 (Cell culture) | | | |
| YCALAPNMMVTNNTFTLK | PXD018241 (Cell culture) | | | |
| YFSGAMDTTSYR | PXD018804 and PXD018241 (cell culture) | | | |

| | | | |
|---|---|---|---|
| YLGTGPEAGLPYGANK | PXD018241 (Cell culture); PXD021328 (Clinical) | PXD021328 and PXD019423 (Clinical) | |
| YLVQQESPFVMMSAPPAQYELK | PXD018241 (Cell culture) | | |
| YMNSQGLLPPK | PXD018241 (Cell culture) | | |
| YMSALNHTK | PXD018241 (Cell culture) | | |
| YNENGTITDAVDCALDPLSETK | PXD018241 (Cell culture) | | Yes |
| YNLPTMCDIR | PXD018241 (Cell culture) | | |
| YNSASFSTFK | PXD018241 (Cell culture) | | |
| YPANSIVCR | PXD018241 (Cell culture) | | |
| YPQVNGLTSIK | PXD018241 (Cell culture) | | |
| YSQLDEEQPMEID | PXD018241 (Cell culture) | | |
| YSTLQGPPGTGK | PXD018241 (Cell culture) | | |
| YTMADLVYALR | PXD018241 (Cell culture) | | |
| YTQLCQYLNTLTLAVPYNMR | PXD018241 (Cell culture) | | |
| YVDNNFCGPDGYPLECIK | PXD018241 (Cell culture) | | |
| YVDNNFCGPDGYPLECIKDLLAR | PXD018241 (Cell culture) | | |
| YVLMDGSIIQFPNTYLEGSVR | PXD018241 (Cell culture) | | |
| YVQIPTTCANDPVGFTLK | PXD018241 (Cell culture) | | |
| YYLGTGPEAGLPYGANK | PXD021328 (Clinical) | PXD021328 (Clinical) | |
